# Tools for measuring sexual and reproductive health and rights (SRHR) indicators in humanitarian settings

**DOI:** 10.1101/2021.03.23.21254101

**Authors:** Céline M. Goulart, Amanda Giancola, Humaira Nakhuda, Anita Ampadu, Amber Purewal, Jean-Luc Kortenaar, Diego G. Bassani

**Author notes:** Corresponding Author: Céline M Goulart.

## Abstract

**Background:** Effective measurement of all health indicators and especially SRHR is difficult in humanitarian settings. Displacement and insecurity due to conflict, natural disasters, and epidemics place women and girls at higher risk of SRHR-related morbidity and mortality and reduce the coverage of essential SRHR services. This scoping review describes the measurement tools, methods, and indicators used to measure SRHR coverage and outcome indicators in humanitarian settings in the past 15 years and presents an accessible dashboard that can be used by governments, researchers and implementing organizations to identify available SRHR measurement tools.

**Methods:** Scientific articles published between January 2004 and May 2019 were identified using Embase, Medline, PsycInfo, CINAHL, Scopus, PAIS index as well as relevant non-peer-reviewed literature available through websites of humanitarian organizations. Publications including data from low- or middle-income countries (LMICs), focused on women and/or girls living in areas impacted by a humanitarian crisis, where data was collected within five years of the crisis were included. Indicators extracted from these publications were categorized according to validated SRHR indicators recommended by the World Health Organization (WHO). Measurement tools, sampling and data collection methods, gap areas (geographical, topical and contextual), and indicators were catalogued for easy access in an interactive Tableau dashboard.

**Results:** Our search yielded 42,081 peer-reviewed publications and 2,569 non-peer-reviewed reports. After initial title and abstract screening, 385 publications met the inclusion criteria. SRHR indicators were categorized into nine domains: abortion, antenatal care, family planning, gender-based violence, HIV and sexually transmitted infections, maternal health, maternal mortality, menstrual and gynecological health, and obstetric care (delivery). A total of 65 tools and questionnaires measuring SRHR were identified, of which 25 were designed specifically for humanitarian settings.

**Discussion:** Although SRHR was measured in humanitarian settings, several gaps in measurement were identified. Abortion and gynaecological health were not consistently measured across included studies or validated WHO indicators. Toolkits and indicators identified in this review may be used to inform future SRHR data collection in humanitarian settings. However, identifying and/or developing innovative data collection methodologies should be a research priority, especially in light of the recent COVID-19 pandemic.

## 1. Background

Sexual and reproductive health and rights (SRHR) are essential for the health and development of all people, as outlined by target 3.7 of the Sustainable Development Goals (SDGs)(1) and the Minimum Initial Service Package (MISP) (2). Women and girls are significantly impacted by humanitarian crises and face SRHR challenges in these contexts. We used the definition of humanitarian crisis defined as *“a serious disruption of the functioning of a community or a society causing widespread human, material, economic or environmental losses which exceed the ability of the affected community or society to cope using its own resources, necessitating a request to national or international level for external assistance. The disaster situation may either be manmade (e*.*g. armed conflict) or a natural phenomenon (e*.*g. drought)”* (3). In 2018, the United Nations Population Fund (UNFPA) estimated that 136 million people had been affected by conflict, hazards, pandemics and displacement, and were in need of humanitarian assistance. Of those, an estimated 34 million were women and girls aged 15-49 years and five million were pregnant (4).

### 1.1 Summary of existing literature

While access to healthcare is generally challenging in humanitarian settings, access to SRHR poses its own unique set of barriers, including poor sanitation, inadequate living conditions and stigma surrounding SRHR services (5, 6). Displacement can negatively impact the power and agency of girls and women to make decisions surrounding sexual activity (7). Good quality SRHR services are not consistently available across a diversity of humanitarian settings (8, 9). Low quality or inaccessible SRHR services can lead to a greater risk of unplanned pregnancy, HIV &sexually transmitted infections (STIs), and greater maternal morbidity and mortality (10).

Effective measurement of SRHR-related indicators in low- and middle-income countries (LMICs) is difficult, and even more so in humanitarian settings. According to the Interagency Field Manual on Reproductive Health in Humanitarian Settings (IAFM), access to some or all of the affected populations may be complicated due to ongoing security issues. Further, where routine data collection strategies are disrupted, and in the few instances where data can still be collected or continue to be produced, data quality is likely compromised and the severity of the quality implications are usually unknown (11). In addition to these factors, the costs of implementing programs in conflict settings are much higher than in non-conflict settings, and the ethical implications are far more complex (12). Many conflict-affected countries are politically fragile and may lack the capacity to provide research practice guidelines and monitor research ethics. This means that researchers could circumvent their own national ethical standards for the use of poorly enforced guidelines. Additionally, humanitarian organizations may face political barriers to the dissemination of sensitive research findings (13).

### 1.2 Objective

In order to streamline data collection and measurement of indicators relevant to the planning, delivery, uptake and outcomes of developmental assistance in such settings, there is a need to understand the coverage of services and SRHR-related morbidity and mortality among women and girls (11). This scoping review describes the measurement tools, methods and indicators used to measure SRHR coverage and outcome indicators in humanitarian settings and identifies gaps in measurement across the domains of SRHR in these contexts. This scoping review also presents an interactive online dashboard with our findings. However, our study does not explore the technical, logistical or financial aspects of conducting research or evaluations in humanitarian settings, which is aptly explored in the IAFM (12).

## 2. Methods

A scoping review provides a broad synthesis of the literature available on a topic of interest, and therefore was our method of choice for exploring the measurement of SRHR in humanitarian settings (14). The research question was defined by inclusion and exclusion criteria and a search of both the peer-reviewed and non-peer-reviewed literature was conducted. This was followed by title abstract screening, full text screening and extraction, each of which were conducted in duplicate, as explained in the sections below (14). This scoping review was conducted in accordance with the Preferred Reporting Items for Systematic Reviews and Meta-Analyses (PRISMA) statement (15).

### 2.1. Definitions and Inclusion Criteria

The World Health Organization (WHO)’s definition of SRHR was used in this review, which includes the domains of abortion, antenatal care (ANC), family planning (FP), gender-based violence (GBV), HIV and sexually transmitted infections (STIs), maternal health, maternal mortality, menstruation and gynecological health, and obstetric care (delivery) (16). The search strategy was based on the domains listed above and related terms relevant to SRHR coverage and morbidity/mortality. See Additional file 1 for full search strategy.

To be included in this review, the humanitarian event had to result in over 1,000 human casualties. The event had to be distinct from latent civil unrest, or long-standing epidemics, such as HIV. This definition was also used to build our second scoping review on gender equality and women’s empowerment (GEWE) (17). Together these papers form a set of scoping reviews on the status of women across humanitarian settings in LMICs.

For a publication to meet the inclusion criteria, it had to:

- Include women and/or girls that had been impacted by a humanitarian crisis in a LMIC. Data must have been collected either during the time of conflict or no more than five years post-event. This included populations living in LMICs who had fled a country with ongoing humanitarian event. Refugee populations living in high-income countries were excluded, as their access to resources and housing may be privileged. This also excluded populations living in LMICs that were not impacted by a humanitarian event.
- Measure quantitative SRHR outcomes.
- Be an interventional or observational study on an SRHR-related topic.
- Be published after January 1st, 2004 to May 2019. Publications before the year 2004 were excluded on the basis of relevance and feasibility.
- Be in the English language or have an English translation available online, as this project did not have the capacity for translation.

Qualitative studies, and those without any measurement methods were excluded. Outcomes that were based on non-empirical data, including models or simulations were excluded. Indicators that included only counts (number of individuals or events) without a clear denominator were also excluded. Indicators measuring intimate partner violence (IPV) and non-partner sexual violence (NPSV) prevalence were excluded from our standard indicator count if they did not capture the experience of violence that occurred in the 12 months preceding measurement, as defined in the WHO standard indicators (16, 18). However, they were counted towards the sexual violence and intimate partner violence indicator type count. See Additional file 2 for detailed inclusion and exclusion criteria.

### 2.2. Search

Scientific articles published between January 2004 and May 2019 were identified using Embase, Medline, PsycInfo, CINAHL, Scopus and PAIS index. The websites and databases of UNFPA, United Nations High Commission for Refugees (UNHCR), Oxfam, Care International, WHO, International Rescue Committee (IRC), International Committee of the Red Cross (ICRC), World Vision, Médecins Sans Frontières (MSF) Field Research, Active Learning Network for Accountability and Performance (ALNAP), Save the Children International (SCI), Sexual Violence Research Initiatives (SVRI), IAWG, Women’s Refugee Commission, Humanitarian Response, Relief Web and Plan International were searched for relevant non-peer-reviewed literature. The first 200 citations were screened from each source and then the full-text documents were screened for eligibility.

### 2.3 Screening Process

The results of the index literature search were uploaded to a screening software that allowed reviewers to work in duplicate. This review used Rayyan – an abstract screening interface as well as a title and abstract prediction classifier based on a machine learning algorithm (19). Details of this tool used for title and abstract screening can be seen in Additional file 3.

Following the title and abstract screening process, conducted in duplicate in Rayyan, the full-texts of publications were also screened in duplicate and assessed for eligibility; an exclusion reason was assigned for publications that did not meet the study criteria.

Relevant systematic reviews identified through the database and non-peer-reviewed searches were manually screened for relevant citations. Ten additional studies were included through this process. The GEWE review also yielded relevant papers that were not retrieved through the SRHR search, contributing 53 additional studies.

### 2.4 Data Extraction and Analysis

Information of interest from the included publications was extracted into a Microsoft Excel spreadsheet. A narrative synthesis was conducted to group indicators thematically into nine different domains of SRHR (20). Within these domains of SRHR, indicators were further sorted into types and subtypes. Extracted and sorted data were then descriptively analyzed using STATA 16, to tabulate frequencies (21). Demographics, measurement methods and tools, and indicators were counted once per study. These counts were then used to describe gaps in measurement across domains.

Indicators extracted from the selected papers were compared to standard SRHR indicators and their corresponding data collection methods from the WHO Core Indicators and the WHO Reproductive Health Indicators (16, 18). If extracted indicators met the WHO recommended definition and study population, they were classified as standard indicators. Indicators that used recommended probability-based sampling methods were noted (16). Indicators from the standard WHO Core and SRHR lists were excluded if the population of interest was men (outside of our study population) or were specific to nationwide rates (e.g. fertility rate), rather than conflict or humanitarian settings. Data collection toolkits and surveys from our included studies are collated in Additional file 4. Toolkits and surveys are mapped to WHO standard indicators in Additional file 5.

Measurement tools, methods, gaps and indicators identified in this analysis were then used to create interactive Tableau dashboards. They are now accessible online, along with the results from our gender equality and women’s empowerment review (22). A link to the online dashboards can be found in Additional file 6.

## 3. Results

The search yielded 42,081 scientific publications and 2,569 reports. After removing duplicates, title/abstract and full text screening, 385 publications met the inclusion criteria and were included in this review. Of these studies, 22% (n=86) were coverage studies, 40% (n=155) were morbidity/mortality studies and 37% (n=144) covered both topics. Additional file 7 includes the full list of studies.

**Figure 1.**
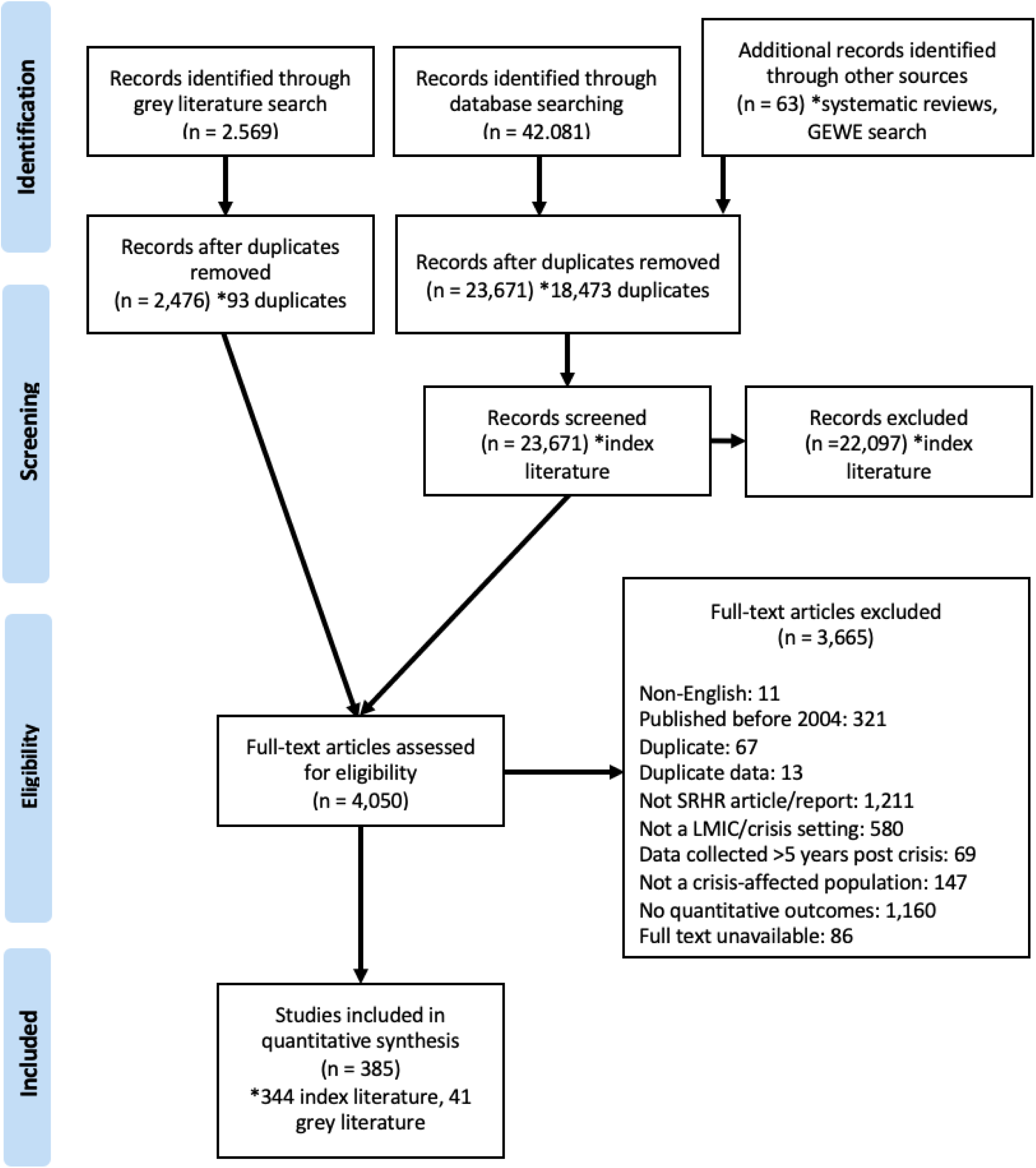
PRISMA chart.

### 3.1. Study Characteristics

Most of the studies included in this review (86%) were conducted with populations affected by conflict within the past five years. Nine percent of studies (n=36) were conducted in disaster and post-disaster settings, and 7% (n=26) in epidemic and post-epidemic settings. Women of reproductive age (WRA), pregnant and postpartum women, and adolescents/adolescent girls were the most represented study populations. Only 1% of studies (n=5) were conducted among children/girls. A total of 32% (n=123) were conducted with refugee populations, 29% (n=111) with populations of internally displaced persons (IDPs), and 37% (n=142) did not report the displacement status of their study population. Over a third of the studies 38% (n=147) collected data on populations living in refugee camps and informal settlements. Population dwelling was not reported for 41% of studies (n=158). Figure 2 includes the study characteristics, stratified by studies on coverage, morbidity/mortality and both.

**Figure 2:**
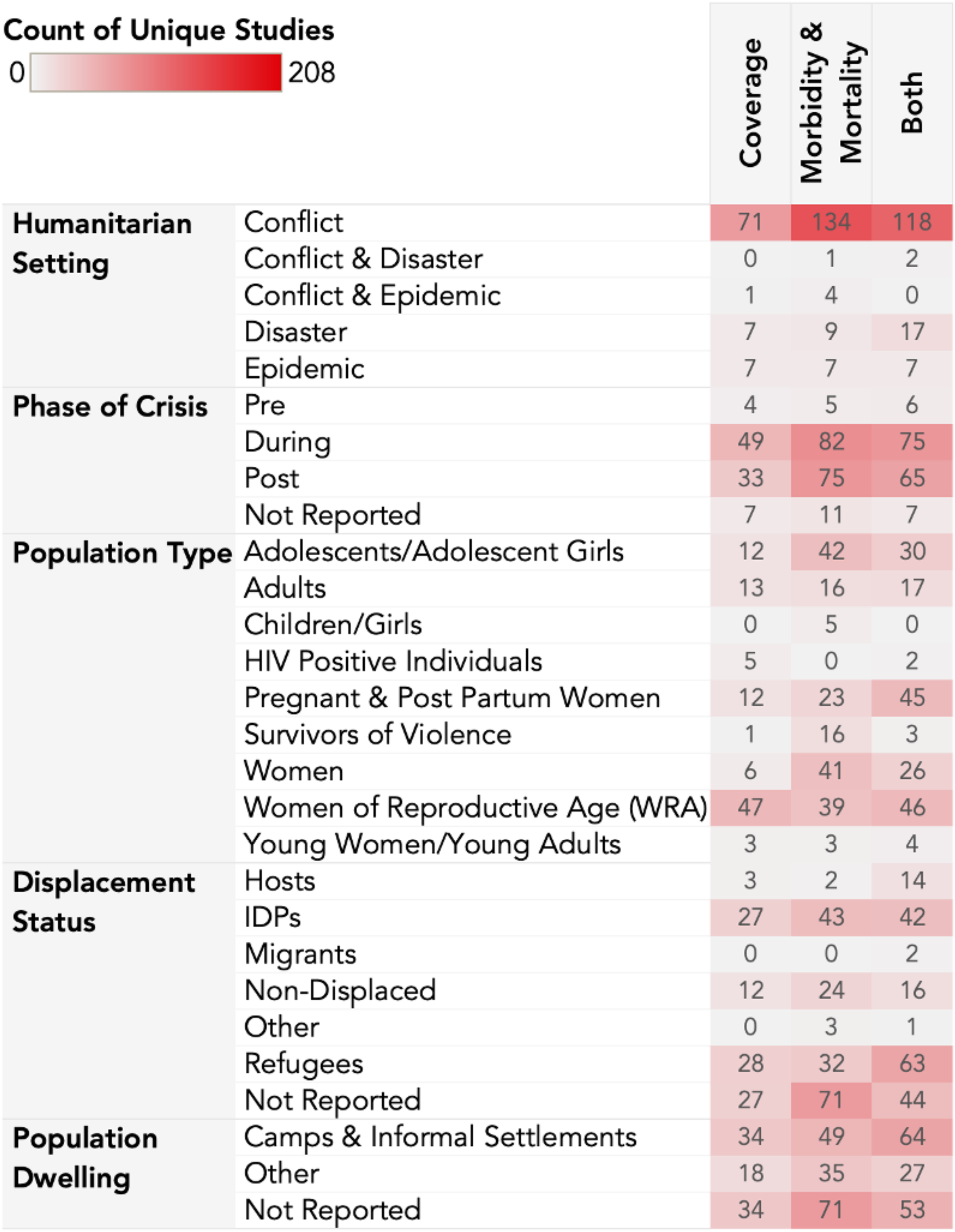
Study Characteristics.

**Figure 3:**
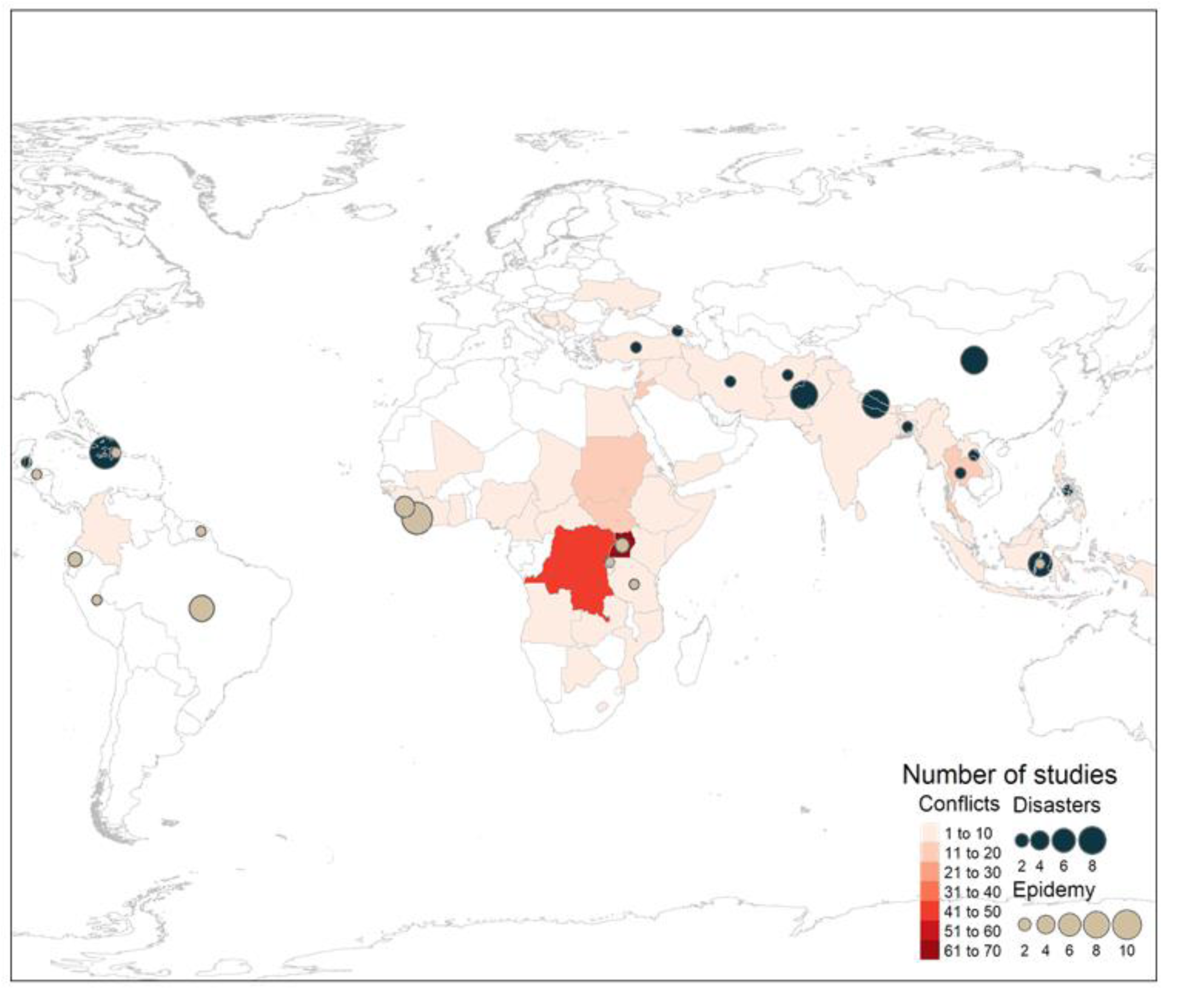
Geographical location and number of studies by type of humanitarian setting.

The studies identified covered 66 countries. Uganda (n=64), the Democratic Republic of the Congo (DRC) (n=45), Lebanon (n=19), Liberia (n=18), and Jordan (n=18) were the most common conflict-affected population sites for data collection. China (n=6), Nepal (n=6), and Pakistan(n=6) were the most frequently studied disaster settings and Sierra Leone (n=4), Liberia (n=5), and Brazil (n=6) were the most common epidemic settings. Figure 3 illustrates all included countries by type of humanitarian setting. Refugee populations originated from 29 countries, the most common being Syria, Myanmar, DRC, Palestine and Somalia. Figure 3 illustrates all the countries of origin of refugee populations in this review.

**Figure 4:**
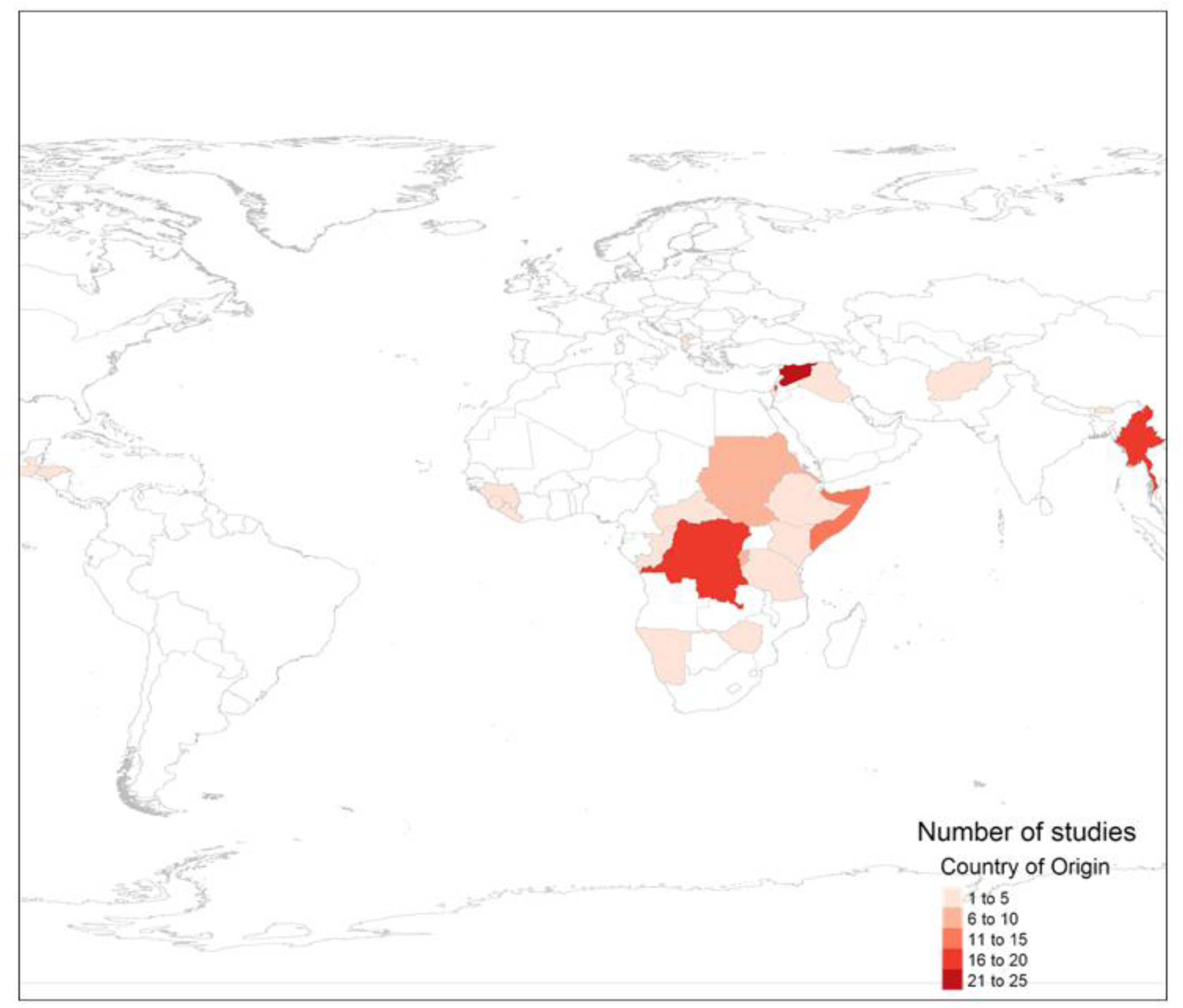
Geographical location and number of studies by country of origin of refugee populations.

### 3.2. Measurement Methods Used

Ninety-five percent (n=367) of the studies in this review were observational, and nearly three quarters of these studies (70%) were cross-sectional (n=269). One hundred and fifty-six studies (41%) were facility level studies, while 214 (56%) were conducted at the population level. Thirteen studies (3%) did not report the data level, and two used both population and facility data (0.5%). Figure 5 illustrates the distribution of the data collection methods, by data level. Only 17 interventional studies (4%), including 11 randomized controlled trials (RCTs) (3%), met the inclusion criteria for our review paper.

**Figure 5:**
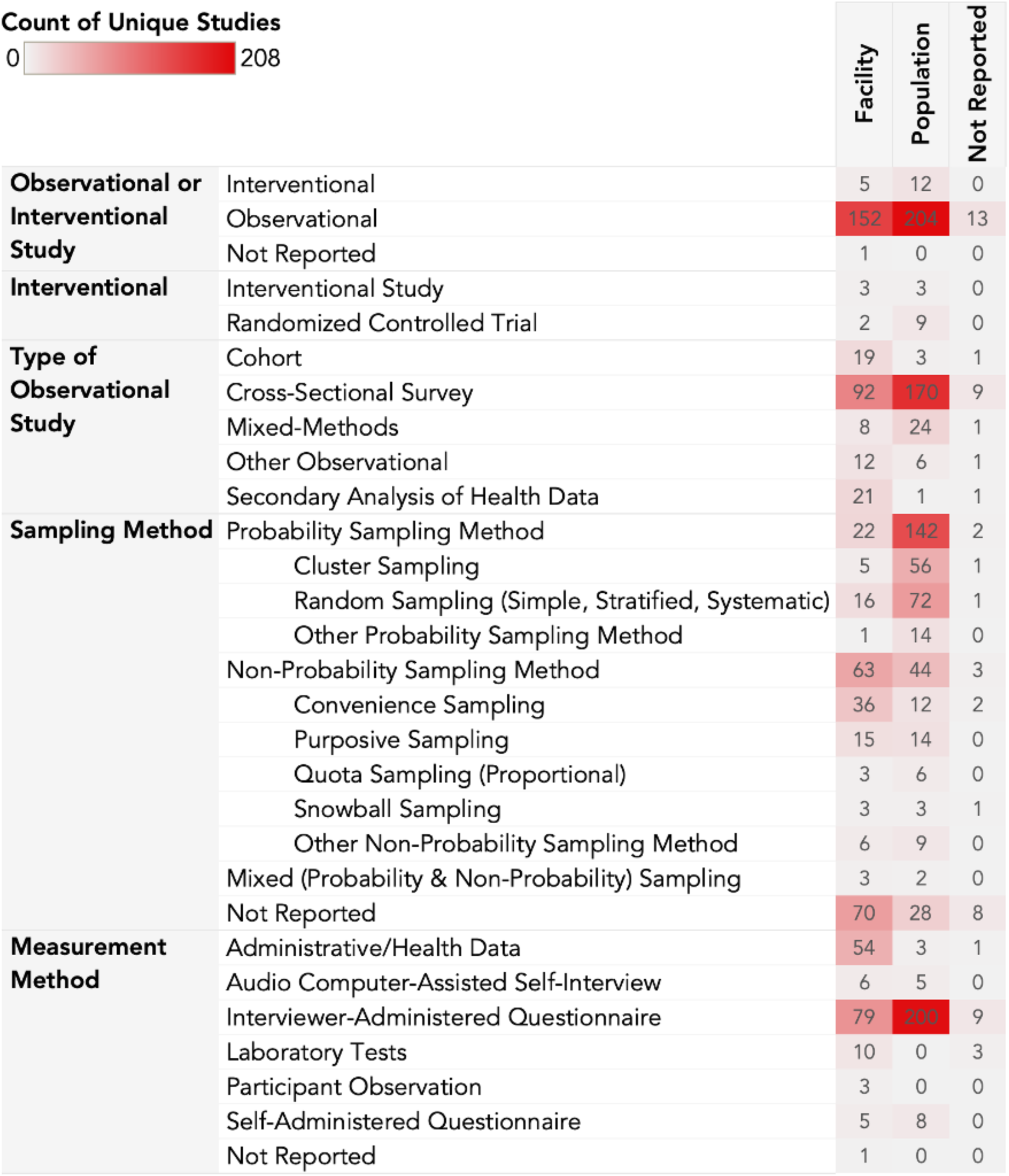
Distribution of the data collection methods.

A total of 166 studies (43%) used probability sampling methods, including cluster sampling (37%, n=62) and simple, stratified and systematic random sampling (54%, n=89). Additionally, 29% (n=110) of the studies employed non-probability sampling, of which convenience (45%, n=50), purposive (26%, n=29) and quota sampling (8%, n=9) were the most common. Sampling methods were not reported for 27% (n=105) of publications.

Approximately 75% of all publications reported used interviewer-administered questionnaires for data collection, 15 % (n=58) of all studies relied on administrative data, 3% (n=13) relied on laboratory tests and self-administered questionnaires, and 3% (n=11) used audio computer-assisted self-interviews (ACASI).

This review identified 65 toolkits and questionnaires measuring SRHR in humanitarian settings, of which 25 were designed specifically for humanitarian settings. The most frequently used toolkits in this review were: the CDC Reproductive Health Assessment Toolkit for Conflict-Affected Women (7% of all studies, n=26), the Demographic and Health Survey (DHS) (6 % of all studies, n=22), and the WHO Multi-Country Study on Women’s Health and Domestic Violence Against Women (5% of all studies, n=18). The CDC Reproductive Health Assessment Toolkit for Conflict-Affected Women used 20 standard indicators, and the Demographic and Health Survey used 21 standard indicators. See Additional file 4 for the full list of toolkits and surveys. The full list of standard indicators can be found in Figure 7.

### 3.3. Domains of SRHR

Figure 6 illustrates the representation of SRHR domains by study count. Indicators of SRHR were categorized into nine domains: abortion, ANC, FP, GBV, HIV and STIs, maternal health, maternal mortality, menstruation and gynecological health, and obstetric care (delivery). The abortion and maternal mortality domains were the least common in our review, found in only n=30 (8%) and n=19 studies (5%), respectively. Menstruation and gynecological health were represented in 44 studies (11%). GBV had the highest representation of studies, with 178 papers (48%) including at least one GBV indicator. Obstetric care (delivery), HIV and STIs, and FP were found in a similar number of studies in our review (n=99, 26%; n=97, 25%; and n=93, 24% respectively).

**Figure 6:**
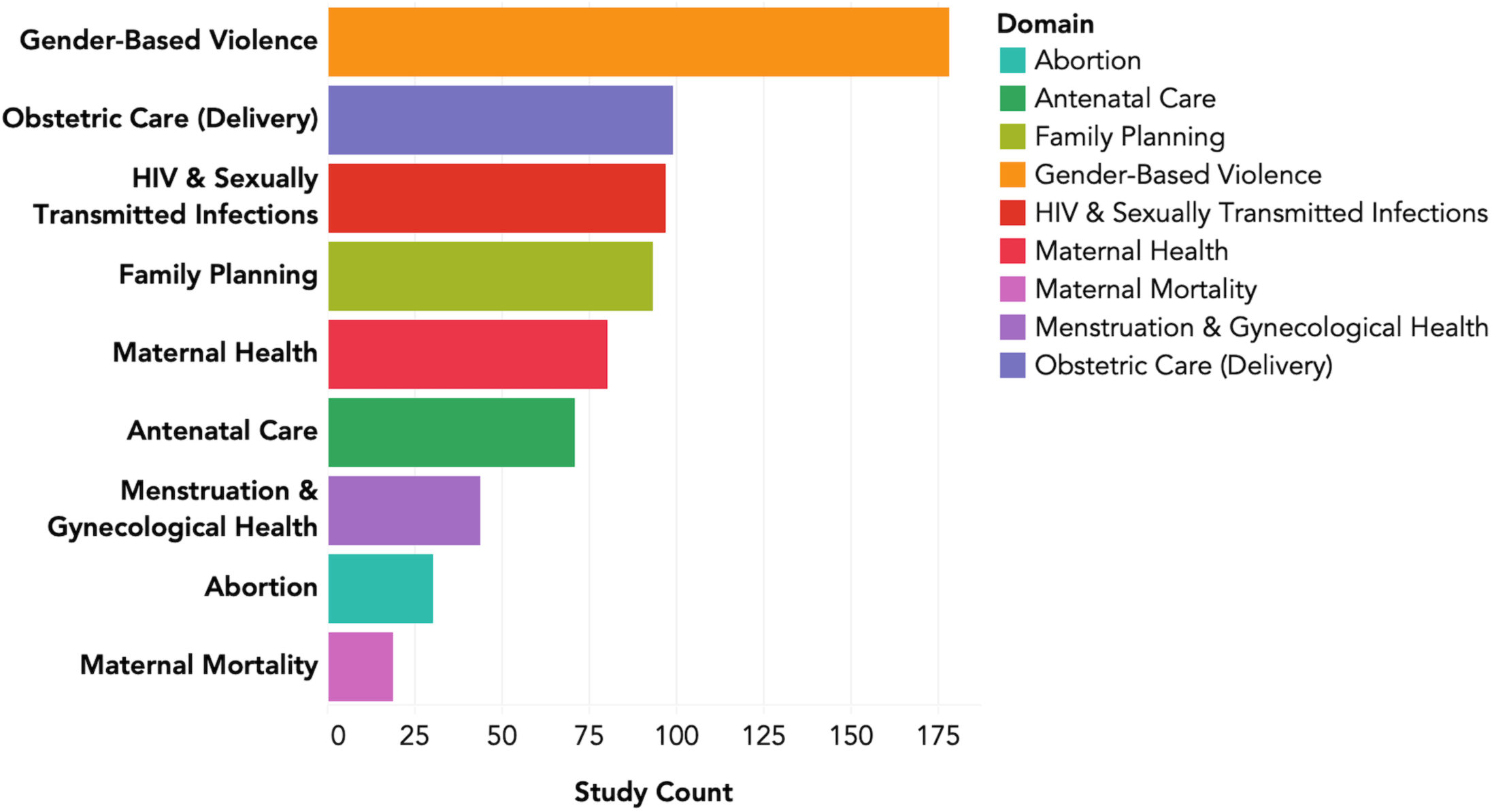
Number of studies by SRHR domain.

### 3.4. Gaps in Measurement

Figure 7 includes all the WHO standard indicators found in this study, by study count, probability methods, toolkit count, and country count. Figure 8 illustrates the various types of gaps within our scoping review. Domains of SRHR were organized into indicator types and then subtypes, which can be found in additional file 8. Indicator types/subtypes that have few, or no check marks represent gaps. Indicator types/subtypes that were present in 15 or more of the 385 papers included in this scoping review are considered common across studies. Standardization exists for indicator types/subtypes that are validated by a WHO standard indicator. Indicators types/subtypes used in five or more countries are considered common across countries. Indicator types/subtypes are considered common in the toolkits if they were present in three or more toolkits retrieved from this review. Finally, an indicator type/subtype is measured in detail when it is measured by the same indicator in three or more studies, or three or more different indicators within one study.

**Figure 7:**
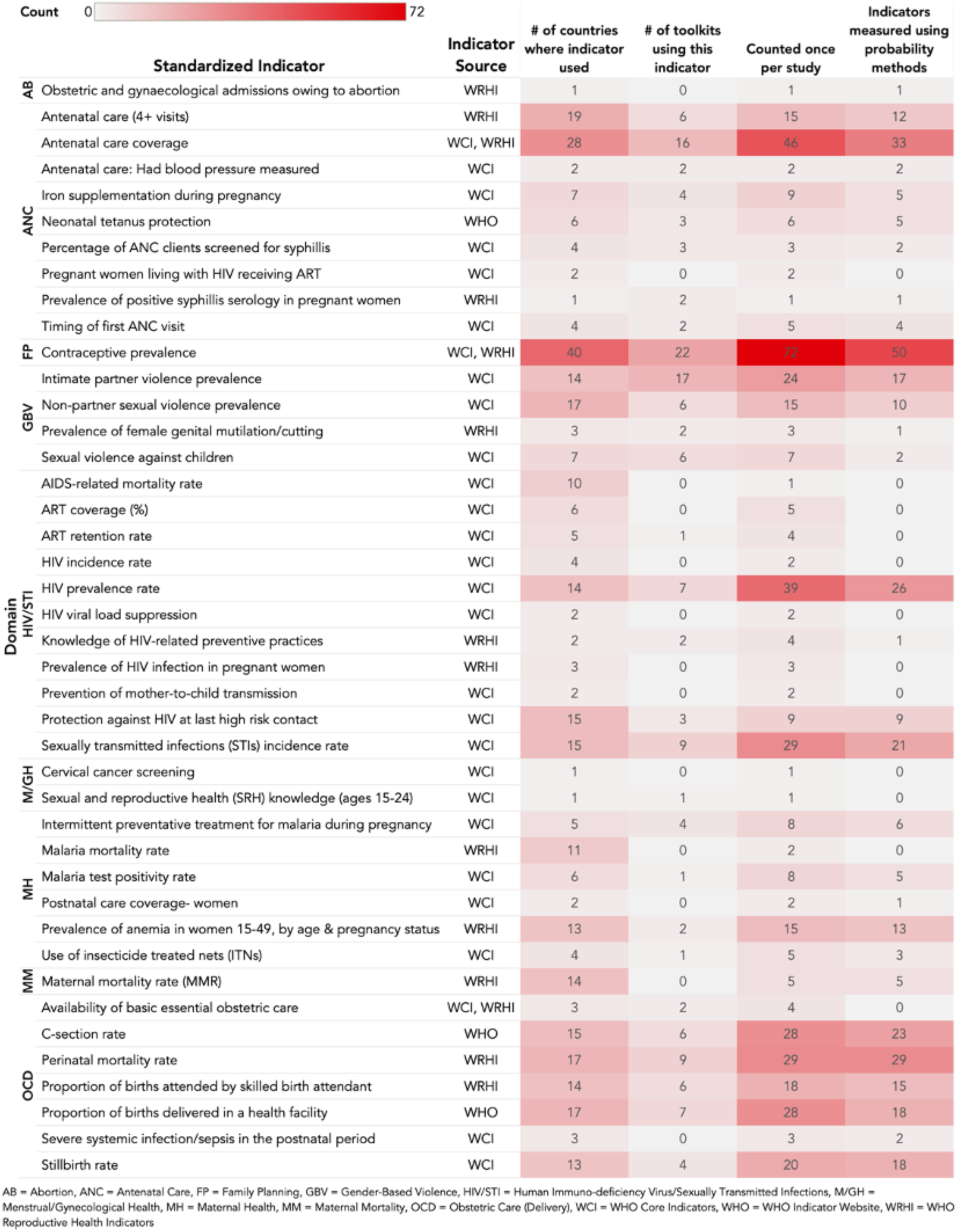
Frequency of studies including standardized indicators by domain.

**Figure 8:**
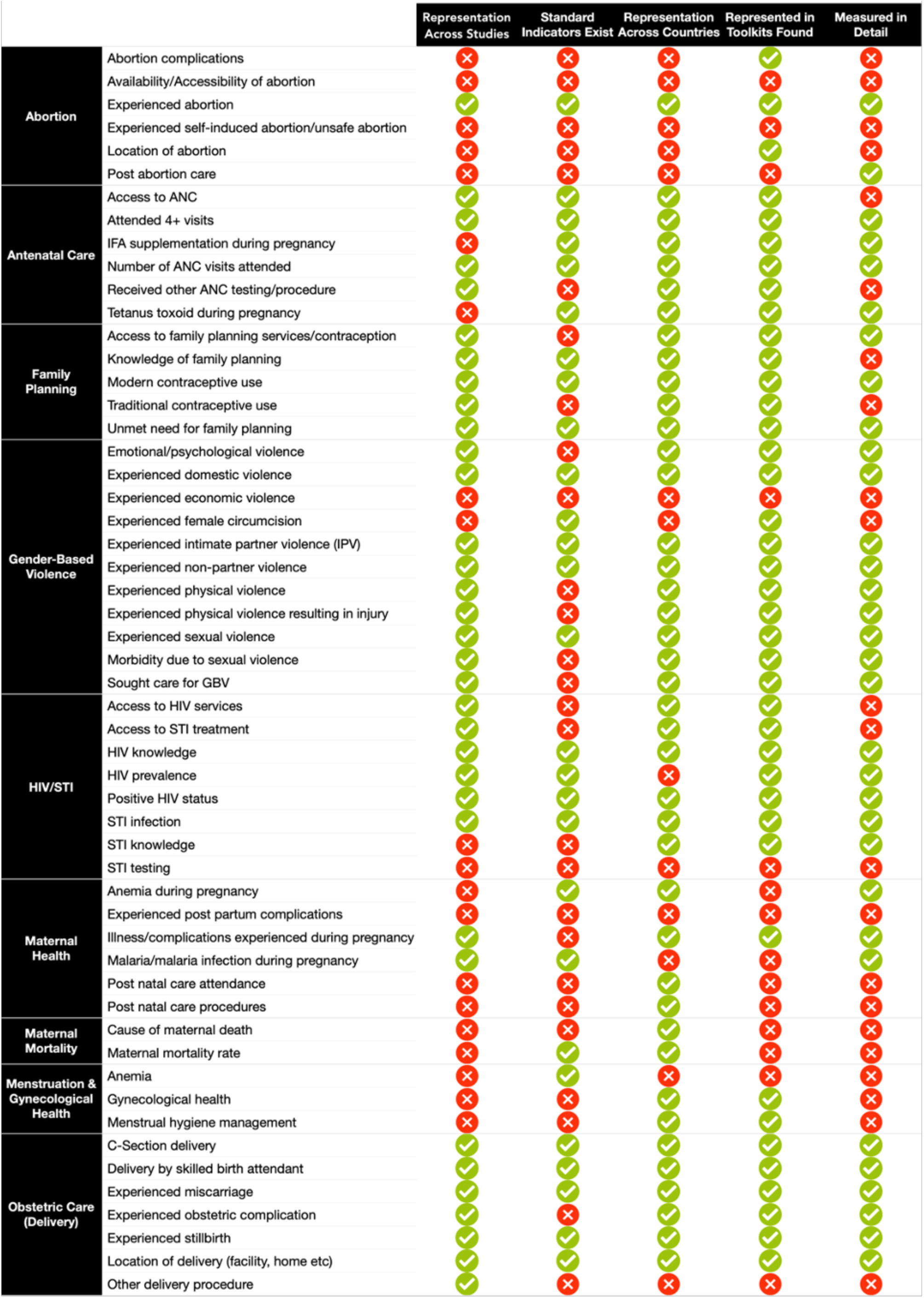
Measurement gap areas.

From our gap mapping exercise, it became clear that some types and subtypes of SRHR were more adequately and frequently measured than others.

#### a. The Pregnancy Continuum

Indicators and tools related to stages of the pregnancy continuum were represented across studies, across countries and across toolkits. ANC and obstetric care were well measured, including the standard indicators ANC coverage (n=46), ANC (four or more visits) (n=15), proportion of births delivered by a skilled birth attendant (n=18), proportion of births delivered in a health facility (n=28), perinatal mortality rate (n=29), stillbirth rate (n=20) and C-section rate (n=28). Contraceptive prevalence was measured in 72 studies, 69% of which used probability methods, and in a total of 40 countries. It also was represented in 22 of the toolkits found in this review, making contraceptive prevalence the most frequent standard indicator measured. However, two indicators related to syphilis (positive status during pregnancy and screening and positive status) were only measured once and thrice respectively. Additionally, the standard indicator postnatal care (PNC) for women was only found in two studies, only one of which used a probability sampling method. Maternal mortality rate (MMR) was only measured in 5 studies, in 14 different countries. Cause of maternal death was measured in 8 studies.

#### b. Gender-Based Violence

In the GBV domain, nine different tools to measure IPV and five tools to measure NPSV prevalence were found in 24 and 15 studies, respectively. The indicator types sexual violence and intimate partner violence were found in 110 and 61 studies respectively. However, indicators measuring female genital mutilation/cutting (FGM/C) were poorly represented. Prevalence of female genital cutting was only measured in three instances in three different countries.

#### c. Abortion

Abortion as a domain was not common in the literature, indicating that there is a lack of measurement of both safe and unsafe abortion in humanitarian settings. Post abortion care (n=5, 1%), self-induced abortion (n=1, 0.3%), and location of abortion (n=4, 1%) were reported in a few studies throughout this review. Abortion complications, unsafe abortion, availability/accessibility of safe abortion were not explicitly measured.

#### d. Menstruation &Sexual Health

Menstruation as an SRHR domain was also not common in this review. Menstrual hygiene management (MHM) and menstrual irregularities were measured in ten studies. There were no WHO standard indicators related to menstruation.

HIV prevalence rate was the most commonly measured standard indicator in the HIV/STI domain, having been represented in 39 studies. Cervical cancer screening was extremely underrepresented in our study (n=1), yet it is classified as a WHO Core Indicator. Cervical cancer screening and sexual and reproductive health (SRH) knowledge were solely measured in one study and one country; they were not well represented in the toolkits.

## 4.0. Discussion

### 4.1. Discussion of Measurement Methods Used

The lack of interventional research in this review may be in part due to the scope of our study, however it could also underscore the difficulty of conducting interventional research among displaced populations; it is challenging to distinguish a control group in conflict settings, and in the absence of this group, the study results are less comparable (23). While the majority of studies in this review employed a cross-sectional study design, a variety of sampling methods were used. Random sampling is the optimal sampling approach for cross-sectional surveys as it reduces the possibility of selection bias and allows results to be more generalizable. It is recommended (along with clustered sampling) to be employed when measuring the WHO Reproductive Health Indicators (16). However, there may be constraints to employing this method in humanitarian settings due to the displacement of study populations (24). Sampling can be challenging in insecure and violent circumstances (25). Lack of resources and infrastructure can limit access to populations and can limit the ability to conduct research, thus compromising the quality and scientific validity of the results (13).

Although a cross-sectional design limits the ability to make causal statements about findings, it was the most common design used in unstable environments (26-28). Non-probability methods are more feasible while conducting research with some marginalized populations, especially in situations with limited resources and research capacity. However, non-probability sampling results in findings that are less representative of the population and less generalizable across settings (29, 30). Additionally, some of the non-random samples within our review were very small, further reducing their generalizability (31). Non-probability sampling methods are also prone to selection bias. For example, employing purposive or convenience sampling in a facility-based study will limit the study population to women who seek care, excluding women for whom care is unavailable or inaccessible (24, 25, 32).

Questionnaires providing self-reported data were the most commonly used measurement instrument. However, these instruments have a number of limitations, in humanitarian settings and in other contexts. Underreporting can occur, especially when the subject matter is sensitive in nature (33, 34). Participants may omit information due to a lack of privacy, especially when data collection takes place in a crowded environment, or near other household members (35). Social desirability bias may occur, as participants may under- or over-report experiences based on the perceived expectations of the researcher (31, 36, 37). Self-reported and interview-administered questionnaires are also prone to recall bias due to the retrospective nature of the questionnaires (25, 31, 38-40). There are limitations to using retrospective administrative data as well, as patient data may not be verifiable (41).

### 4.2. The Pregnancy Continuum

ANC (four or more visits), ANC coverage, proportion of births delivered in a health facility, proportion of births attended by a skilled birth attendant, C-section rate, perinatal mortality rate, and stillbirth rate are well recognized indicators of SRHR. These are well established and validated indicators found throughout the literature, using probability sampling methods (16, 18). From our review, it is evident that antenatal care, family planning, and obstetric care (delivery) are priority areas for non-governmental organizations (NGOs) and governments when responding to humanitarian crises. It is important to note that though the WHO now recommends eight ANC visits, four visits or more was chosen as the indicator as it was used in most ANC studies included in this review (42).

However, post-natal care (PNC) for mothers is insufficiently captured by the data on SRHR in LMICs in crisis, especially compared to other time periods along the pregnancy continuum. The available data on PNC included in this scoping review focuses predominantly on attendance and procedures, and to a lesser extent postpartum depression and other postpartum complications. Much of PNC focuses on newborn rather than maternal health (43), which explains the dearth of data on PNC in our scoping review, since our population of interest is women of reproductive age. In spite of this, *PNC coverage for women* is a standard WHO indicator, demonstrating its significance as a measure of SRHR (16).

The domain of maternal mortality as a whole and mortality rates, including AIDS-related mortality rate and MMR, were not commonly found in the literature. Maternal mortality is difficult to estimate in humanitarian settings. A review on the measurement of maternal mortality in humanitarian settings found that there was selection bias in how maternal deaths were measured (40). Few studies confirmed deaths with verbal autopsy or medical records. Studies conducted at the household level relied on recall from family members. More representative samples are needed, and deaths need to be reconfirmed to increase the reliability of the data (44). However, it is also possible that our study criteria excluded some maternal mortality studies that relied on statistical modelling to estimate MMR at the national level.

### 4.3. Gender-Based Violence (GBV)

While GBV was the most frequently measured domain in this review, FGM/C was poorly represented. FGM/C is defined as the partial or total removal of external female genitalia for non-medical reasons. There are no health benefits to FGM/C for women and girls, and FGM/C is known to lead to a number of health problems, including severe bleeding, problems urinating, cysts, infections, complications with childbirth and in some cases, death (45). FGM/C is contextual, and its frequency depends on the cultural norms of the population. In some emergency settings, there are too few individuals in the community who practice FGM to make tracking a priority for resource allocation (46). FGM/C can be challenging to define based on severity, practice, cultural beliefs and stigma, rendering it difficult to measure quantitatively at the population level (46, 47). Family separation, social disruption, health issues, and poverty caused by crises make access to FGM/C prevention interventions and programs difficult; FGM/C responses tend to be secondary in emergency settings where basic provisions of food, water, and healthcare are a priority (46). One study excluded FGM/C from their GBV screening tool because it was expected that much of the adult female population would have experienced this practice, and the consequent number of referrals would overwhelm the health system (48, 49).

In addition to FGM/C, economic violence was not commonly found in the GBV domain. Economic violence can negatively impact the sexual and reproductive health of women in crisis. However, because economic violence is not a measurement of SRH, it is better-suited (much like other GBV subtypes) to the GEWE scoping review; economic violence is a direct indication of inequality and/or disempowerment (17).

### 4.4. Abortion

Despite increased funding and capacity to deliver SRH interventions by NGOs, comprehensive abortion care remains as a significant gap in adolescent sexual and reproductive health services in humanitarian settings (12). Abortion is complicated to measure globally, as it is stigmatized and therefore ca be underreported (50).

Barriers to accessing abortion in humanitarian settings result in challenges to measuring safe abortion. In order for abortion to be considered safe, it must be conducted by a trained health professional using a recommended method. If neither or only one of these criteria is met, the abortion is considered unsafe ((50, 51). At the participant level, barriers to accessing abortion services include stigma, confidentiality (52), availability, shortage in trained workers and equipment, and religious barriers (51).

There are also barriers to abortion and abortion measurement at the organization/facility level. Abortion services may be perceived as too complicated to provide in the humanitarian space, despite being safe and well-established medical procedures (53). Further, many researchers have shown that post-abortion care services can be offered in politically unstable and culturally conservative settings where abortion is stigmatized (54, 55). Another potential barrier to the provision and measurement of abortion is that some donors do not fund abortion services (53). Additionally, abortion is illegal in some settings, although 190 out of 196 countries globally do permit abortion, some under limited circumstances (53); for example, in the DRC, abortion is legal if two physicians agree that it is necessary to save the life of the mother (56).

Though abortion is supported by international treaties and laws, there is still a need for decriminalization, particularly in conflict-settings where there are high levels of pregnancy as a result of sexual violence (57, 58). The legalization of abortion will also promote a higher standard for these services and reduce instances where untrained providers offer these procedures illegally (52).

Abortion is represented by just one standard indicator in this review: obstetric and gynecological admissions owing to abortion (16). This indicator does not sufficiently capture the true prevalence of abortions in humanitarian settings or if unsafe abortions are prevalent within the community. Measuring the proportion of admissions owing to abortion will underestimate the actual abortion rate in the region (53). In 2019, 92% of women of reproductive age in Sub-Saharan Africa lived in countries with restrictive laws surrounding abortion. From 2010-2014, 77% of abortions were unsafe, compared to 45% globally (50). In settings where abortion services are illegal or inaccessible, the magnitude of unsafe abortions and associated maternal mortality should be measured. Further, there should be additional standard indicators to capture the stigma and laws that surround abortion. Not only does abortion need to be funded and accessible, it also needs to be measured using validated methods that are comprehensive.

### 4.5. Menstruation &Gynecological Health

Very few papers contained within our scoping review focused on MHM and other topics external to the pregnancy continuum, such as unspecified gynecological illnesses and related complications. These gaps could be attributed to a variety of factors: the inherent nature of humanitarian responses (prioritizing life-saving measures for the most vulnerable at the outset), the preference for qualitative methods to assess certain topics, or in some cases, the lack of standard indicators. Even where standardization exists, indicators may be overlooked; this may be because they pertain to non-communicable diseases, they are not immediately life-threatening or they do not affect a large number of people. This was the case for cervical cancer screening measurement in humanitarian settings.

Within the domain of menstruation and gynecological health, indicators were extracted for MHM including knowledge, satisfaction and type of menstrual product used, menstrual irregularities, and SRH symptoms, such as anemia, and reproductive/gynecological complaints and infections. Noticeably absent from our findings was an association between any of the extracted SRH symptoms and menstruation. For example, anemia was not connected to menstruation, despite its association with menarche. Despite the guidance given by the IAFM (12), and the MISP Distance Learning Module on Menstrual-related Responses (2), menstrual-related indicators in this scoping review do not reflect or measure the complexity/granularity that is outlined by these responses.

The Sphere Handbook recommends that adolescent girls and women are consulted about their needs and preferences with regards to MHM, yet it does not contain standards for these consultations (59). There are also no standard indicators for menstruation or MHM in either of the WHO Core Indicators or the WHO Reproductive Health Indicators (16, 18), which reflects the lack of consensus on the optimal way to monitor and evaluate these interventions (60). There is, however, a minimum standard for responding to the MHM needs of adolescent girls, which includes the supply of sanitary materials, and in more comprehensive responses, the delivery of puberty education to 10-14 year olds (61). Nevertheless, in the absence of standards, both for the recommended consultations with adolescent girls and women and for the indicators used to monitor and evaluate menstruation and MHM, data may be poorly collected, or missing entirely. Notable organizations and resources in this arena clearly define what should be implemented in these situations with regards to MHM, but not how they should be implemented or maintained, or whether any novel menstrual products should be introduced in these settings (60).

### 4.6 Sexual Health

Cervical cancer ranks as the leading cause of cancer death in 42 countries, predominantly in Sub-Saharan Africa and Southeast Asia. However, many of these countries were not represented within our scoping review, as they were not humanitarian settings (62). While the WHO recommends that unvaccinated women (between the ages of 30-49) are screened for cervical cancer, this may not be prioritized in countries where the prevalence is low, resources are constrained (62), or where there may be high vaccination rates in childhood.

STI testing also represented a notable gap in our scoping review. Overall, HIV-related indicators are measured more often than other STI-related indicators. HIV may be measured more commonly than other STIs, in conflict-settings because there is a body of literature that shows how conflict and the associated impacts of conflict, such as weak social and physical infrastructure, poor health, and untreated STIs, increase the risk of HIV transmission (63). This heightened risk results from increased vulnerability (due to sexual violence, poverty, and breakdown of public health education) and increased exposure opportunity (through casual or transactional sex and interactions with individuals who partake in high-risk behaviors) (63).

Overall, our scoping review yielded more tools focusing on reproductive and maternal health than sexual health-related topics (such as STIs). More emphasis must be placed on the diverse needs between groups (including, but not limited to, young girls, lesbian, gay, bisexual, transgender, queer, intersex and asexual youth, and those with disabilities) and on understanding the perceptions of and information provided to girls and women through professional and traditional services (12).

### 4.7. Limitations

This scoping review has a number of limitations. First, studies were only included if they were published or available in English, which reduced the representation of research from non-English researchers and journals. Second, studies were only extracted if they contained indicators with measured outcomes. This prevented some toolkits and checklists from being included in our review if they were found directly in the grey literature. Studies that exclusively used qualitative research methods were also excluded, which may have impacted the level of detail, particularly on lived experiences, contained within the results of our review (32). Third, indicator results reported as crude numbers were not extracted, which excluded a number of facility-based studies. Fourth, this review did not include nationwide data. For example, national maternal mortality and fertility rates were poorly represented or not included, limiting the ability to compare standard indicators across countries. Fifth, we identified ‘gaps’ using a standardized list of indicators that are used to measure SRHR in non-conflict/non-humanitarian settings, which were not specific to humanitarian settings. As mentioned above, data collection in humanitarian settings is challenging. We wanted to see if it was feasible to measure globally accepted SRHR indicators in this context. Finally, the studies contained within this scoping review were heteronormative and gender binary; GBV indicators contain the latent assumption that the (female) participant was married/in a partnership with a male individual, and all indicators were based on biological sex rather than gender-identity.

## 5. Conclusions

There was great variation in the quality and range of tools, methods and indicators used to measure SRHR in humanitarian settings. While a wide variety of sampling methods were used in these studies, cross-sectional surveys made up the majority of study designs. GBV and the pregnancy continuum dominated the domains of study. The WHO standard indicators provided a benchmark for measuring SRHR in these settings, however, not all domains of SRHR were well-represented in these standards; measurements of abortion and menstruation were lacking in humanitarian settings and should be captured by standard SRHR indicators.

The Tableau dashboard created from this scoping review provides an interactive and centralized resource for SRHR measurement, which can be updated as research in humanitarian settings evolves. It is our hope that toolkits and indicators identified in this review will inform future data collection in humanitarian settings. Climate change and the unprecedented COVID-19 pandemic could increase conflict, displacement and access to health services, further jeopardizing the measurement of SRHR in all settings, making the identification and/or development of innovative data collection methodologies a research priority.

## Data Availability

The datasets generated during and/or analyzed during the current study are available from the corresponding author on reasonable request.

https://public.tableau.com/profile/humairanakhuda#!/vizhome/SRHRGEWEScopingReviewStory_Final_Nov16/SRHRGEWEStory

## Supplementary information

Additional file 1. Search strategy. This file includes the search strategy for all 6 databases. (PDF)

Additional file 2: Inclusion and exclusion criteria. This files includes the inclusion and exclusion criteria for the review. (PDF)

Additional file 3: Rayyan screening tool. This file describes the machine learning software used in the title/abstract stage of the review. (PDF)

Additional file 4: Data collection toolkits and surveys. This is a list of all tools utilized by studies in this review. (PDF)

Additional file 5: Toolkits, Surveys and Standard Indicators. This file looks at how many standard indicators are included in each tool found. (PDF)

Additional file 6. Measurement of Sexual and Reproductive Health and Rights, Gender Equality, and Women’s Empowerment in Humanitarian Settings. This link leads to the interactive Tableau dashboard. https://canwach.ca/initiatives/canadian-collaborative-for-global-health/improving-measurement-sexual-and-reproductive-health-and-rights-womens-empowerment-and-gender/

Additional file 7: Study List. Includes list of all included index and grey literature. (PDF)

Additional file 8: SRHR Domains, Indicator Types, and Indicator Subtypes. This file includes the calculated frequency of each SRHR indicator type and subtype. (PDF)

### Abbreviations

ACASI: audio computer-assisted self-interviews;
ALNAP: Active Learning Network for Accountability and Performance;
ANC: Antenatal Care;
CanWaCH: Canadian Partnership for Women and Children’s Health;
DHS: Demographic and Health Survey;
FGM/C: Female Genital Mutilation/Cutting;
FP: Family Planning;
GBV: Gender-Based Violence;
GEWE: Gender Equality and Women’s Empowerment;
HIV: Human Immunodeficiency Virus;
IAFM: Interagency Field Manual;
IAWG: Inter-Agency Working Group;
ICRC: International Committee of the Red Cross;
IDP: Internally Displaced Persons;
IRC: International Rescue Committee;
LMICs: Low- and Middle-Income Countries;
MHM: Menstrual Hygiene Management;
MISP: Minimum Initial Service Package;
MMR: Maternal Mortality Rate;
MSF: Médecins Sans Frontières;
NGOs: Non-Governmental Organizations;
PNC: Post-Natal Care;
PRISMA: Preferred Reporting Items for Systematic Reviews;
SCI: Save the Children International;
SDGs: Sustainable Development Goals;
SRHR: Sexual and Reproductive Health and Rights;
STIs: Sexually Transmitted Infections;
SVRI: Sexual Violence Research Initiatives;
UNFPA: United Nations Population Fund;
UNHCR: United Nations High Commission for Refugees;
WHO: World Health Organization;
WRA: Women of Reproductive Age

## Declarations

Ethics approval and consent to participate: N/A

### Consent for publication

N/A

## Authors’ contributions

CG coordinated the study, analyzed the data, planned, wrote and edited the manuscript, directed the team. She also participated in screening, data extraction and cleaning of the study. AG, AA and AP screened, extracted and cleaned the data. They also contributed to manuscript writing and editing. HN created all the Tableau visuals and is responsible for the development of the online Interactive Tableau platform. She also edited and contributed to the writing process. JLK assisted with the machine learning aspect of the project, and wrote the corresponding appendix. DB is the principal investigator and was responsible for the creation of this project in partnership with CanWaCH.

## Acknowledgements

Canadian Partnership for Women and Children’s Health (CanWaCH), Canadian Red Cross, Luay Basil, Katie McLaughlin, Mariella Munyuzangabo, Neil Yang &Malasha D’souza.

