## Additional file 1 for "Tools for measuring sexual and reproductive health and rights (SRHR) indicators in humanitarian settings"

### Additional file 1: Search Strategy

#### Database(s): CINAHL Modified Search

#### Context Concept

1. MH (disasters or emergencies or natural disasters or mass casualty incidents)
2. (MH "Crime Victims") OR (MH "Victims")
3. AB ((disaster or emergenc*) N2 victim*) OR TI ((disaster or emergenc*) N2 victim*)
4. AB ((disaster or disasters or catastrophe or catastrophes) N5 (environ* or human* or manmade or "man made" or nature or natural or weather)) OR TI ((disaster or disasters or catastrophe or catastrophes) N5 (environ* or human* or manmade or "man made" or nature or natural or weather))
5. AB ("mass casualty" or "mass casualties" or "mass fatalities" or "mass fatality") OR TI ("mass casualty" or "mass casualties" or "mass fatalities" or "mass fatality")
6. AB ((crisis or crises) N5 (environ* or human* or manmade or "man made" or nature or natural or weather or setting*)) OR TI ((crisis or crises) N5 (environ* or human* or manmade or "man made" or nature or natural or weather or setting*))
7. AB ((crisis or crises or conflict) N3 (affected)) OR TI ((crisis or crises or conflict) N3 (affected))
8. (MM "War+") OR (MM "Holocaust") OR (MM "War Crimes+")
9. (MH "Violence") OR (MH "Exposure to Violence")
10. AB ("warfare and armed conflict*" or warfare or "armed conflict*" or "war crime*" or "ethnic cleansing*" or "gas poisoning" or genocide or holocaust or "war exposure") OR TI ("warfare and armed conflict*" or warfare or "armed conflict*" or "war crime*" or "ethnic cleansing*" or "gas poisoning" or genocide or holocaust or "war exposure")
11. AB ("afghan campaign" or "gulf war" or "iraq war" or "war time" or "wartime" or "war torn" or "war affected" or "insurgency" or "intra conflict") OR TI ("afghan campaign" or "gulf war" or "iraq war" or "war time" or "wartime" or "war torn" or "war affected" or "insurgency" or "intra conflict")
12. AB ((armed or zone or political or civil or setting*) N3 (conflict or conflicts or attack or attacks or war or wars or "no fly")) OR TI ((armed or zone or political or civil or setting*) N3 (conflict or conflicts or attack or attacks or war or wars or "no fly"))
13. AB (fragile N2 (state* or countr* or nation* or situation* or setting*)) OR TI (fragile N2 (state* or countr* or nation* or situation* or setting*))
14. AB ("Post conflict" or "postconflict" or "post war" or peacebuilding or peacekeeping) OR TI ("Post conflict" or "postconflict" or "post war" or or peacebuilding or peacekeeping)
15. AB (war N3 related) OR TI (war N3 related)
16. AB ("militant group#" or "militant organi?ation#" or militia or combatant or rebel*) OR TI ("militant group#" or "militant organi?ation#" or militia or combatant or rebel*)
17. MH (disease outbreaks)
18. AB ("disaster medicine" or "disaster outbreak*" or epidemic* or "disease outbreak*") OR TI ("disaster medicine" or "disaster outbreak*" or epidemic* or "disease outbreak*")
19. MH (Emergency Medical Services)
20. AB ((emergency or emergencies) N5 (environ* or human* or manmade or "man made" or nature or natural or weather or complex)) OR TI ((emergency or emergencies) N5 (environ* or human* or manmade or "man made" or nature or natural or weather or complex))

21. MH (starvation)
22. AB (famine or famines or starvation or starvations) OR TI (famine or famines or starvation or starvations)
23. AB (avalanche# or cyclone# or drought# or earthquake# or flood* or hurricane# or landslide# or "land slide#" or mudslide# or "mud slide#" or storm# or tornado* or tsunami# or typhoon# or volcanic or rubble) OR TI (avalanche# or cyclone# or drought# or earthquake# or flood* or hurricane# or landslide# or "land slide#" or mudslide# or "mud slide#" or storm# or tornado* or tsunami# or typhoon# or volcanic or rubble)
24. MH (refugees)
25. AB (evacuee or evacuees or refugee or refugees or squatter or squatters or transients or "asylum seeker") OR TI (evacuee or evacuees or refugee or refugees or squatter or squatters or transients or "asylum seeker")
26. (MH "Humanitarian Aid") OR (MH "Rescue Work")
27. AB ((rescue or relief or aid) N3 (plan or plans or activity or activities or agency or agencies)) OR TI ((rescue or relief or aid) N3 (plan or plans or activity or activities or agency or agencies))
28. AB ("aid plan" or "aid work" or "relief plan" or "relief work" or "rescue plan" or "rescue work") OR TI ("aid plan" or "aid work" or "relief plan" or "relief work" or "rescue plan" or "rescue work")
29. AB ((staff* oor worker#) N3 (relief or aid or rescue)) OR TI ((staff*or worker#) N3 (relief or aid or rescue))
30. AB (humanitarian assistance*) OR TI (humanitarian assistance)
31. AB (humanitarian N5 (aid or response or relief or crisis or crises or emergency or emergencies or disaster or disasters)) OR TI (humanitarian N3 (aid or response or relief or crisis or crises or emergency or emergencies or disaster or disasters))
32. MH (altruism)
33. AB (humanitarianism or altruism) OR TI (humanitarianism or altruism)
34. AB ("displaced children" or "displaced child" or "displaced families" or "displaced family" or "displaced individuals" or "displaced internally" or "displaced men" or "displaced people" or "displaced peoples" or "displaced person" or "displaced persons" or "displaced population" or "displaced populations" or "displaced women" or "displaced adolescent" or "displaced adolescents" or "forced displacement" or "forced displacements" or "forcibl* displace*" or "internal displaced" or "internal displacement" or "internally displaced" or "population displaced" or "population displacement" or "forced migration" or "migrant*") OR TI ("displaced children" or "displaced child" or "displaced families" or "displaced family" or "displaced individuals" or "displaced internally" or "displaced men" or "displaced people" or "displaced peoples" or "displaced person" or "displaced persons" or "displaced population" or "displaced populations" or "displaced women" or "displaced adolescent" or "displaced adolescents" or "forced displacement" or "forced displacements" or "forcibl* displace*" or "internal displaced" or "internal displacement" or "internally displaced" or "population displaced" or "population displacement" or "forced migration" or "migrant*")
35. (MH "refugee camps")
36. AB ((camp or camps) N3 (refugee or transit or displace* or temporary or informal)) OR TI ((camp or camps) N3 (refugee or transit or displace* or temporary or informal))
37. AB (settlement# N3 (temporary or informal)) OR TI (settlement# N3 (temporary or informal))

**Population Concept**

1. AB ((displace* or refugee#) N3 (child* or famil* or men or wom?n or individual* or adolescent* or people* or population* or person* or girl* or boy* or youth)) OR TI ((displace* or refugee#) N3 (child* or famil* or men or wom?n or individual* or adolescent* or people* or population* or person* or girl* or boy* or youth))
2. (MH "Young Adult") OR (MH "Adolescence")
3. AB (adolescen* or teen* or youth# or "young adult#" or girl* or boy* or "young wom?n" or "young girl*" or "young boy*") OR TI (adolescen*or teen* or youth# or "young adult#" or girl* or boy* or "young wom?n" or "young girl*" or "young boy*")
4. (MH "Expectant Mothers") OR (MH "Expectant Parents") OR (MH "Mothers") OR (MH "Adolescent Mothers") or (MH “Women”)
5. AB ("mother#") OR TI ("mother#”)
6. MH (female)
7. AB (parturients) OR TI (parturients)
8. AB ("sexually active") OR TI ("sexually active")
9. MH (refugees)
10. AB (refugee or refugees) OR TI (refugee or refugees)
11. AB (women N2 reproductive N2 age) OR TI (women N2 reproductive N2 age)

**Low- and Middle-Income Countries**

12. MH (Developing Countries) OR AB (Developing Countries) OR TI (Developing Countries)
13. (MH "Low and Middle Income Countries")
14. MH (Africa or Asia or or West Indies or South America or Latin America or Central America) OR AB (Africa or Asia or West Indies or South America or Latin America or Central America) OR TI (Africa or Asia or West Indies or South America or Latin America or Central America)
15. MH (Afghanistan or Albania or Algeria or American Samoa or Angola or Armenia or Armenian or Azerbaijan or Bangladesh or Benin or Byelarus or Byelorussian or Belarus or Belorussian or Belorussia or Belize or Bhutan or Bolivia or Bosnia or Herzegovina or Hercegovina or Botswana or Brasil or Brazil or Bulgaria or Burkina Faso or Burkina Fasso or Upper Volta or Burundi or Urundi or Cambodia or Khmer Republic or Kampuchea or Cameroon or Cameroons or Cameron or Camerons or Cameroun or Cape Verde or Cabo Verde or Central African Republic or Chad or China or Colombia or Comoros or Comoro Islands or Comores or Mayotte or Congo or Zaire or Costa Rica or Cote d'Ivoire or Ivory Coast or Cuba or Djibouti or French Somaliland or Dominica or Dominican Republic or East Timor or East Timur or Timor Leste or Ecuador or Egypt or United Arab Republic or El Salvador or Equatorial Guinea or Eritrea or Ethiopia or Fiji or Gabon or Gabonese Republic or Gambia or Gaza or Georgia or Georgia Republic or Georgian Republic or Ghana or Gold Coast or Grenada or Guatemala or Guinea or Bissau or Guiana or Guyana or Haiti or Honduras or India or Maldives or Indonesia or Iran or Iraq or Jamaica or Jordan or Kazakhstan or Kazakh or Kenya or Kiribati or Korea or Democratic People's Republic of Korea or Kosovo or Kyrgyzstan or Kirghizia or Kyrgyz Republic or Kirghiz or Kirgizstan or Lao PDR or Laos or Lebanon or Lesotho or Basutoland or Liberia or Libya or Macedonia or Madagascar or Malagasy Republic or Malaysia or Malaya or Malay or Sabah or Sarawak or Malawi or Nyasaland or Mali or Marshall Islands or Mauritania or Mauritius or Agalega Islands or Mexico or Micronesia or Middle East or Moldova or

Moldovia or Moldovian or Mongolia or Montenegro or Morocco or Ifni or Mozambique
or Myanmar or Myanma or Burma or Namibia or Nauru or Nepal or Nicaragua or Niger or
Nigeria or Pakistan or Papua New Guinea or Palestine or Paraguay or Peru or Philippines
or Philipines or Phillipines or Phillippines or Romania or Rumania or Roumania or Russia
or Russian or Rwanda or Ruanda or Saint Lucia or St Lucia or Saint Vincent or St Vincent
or Grenadines or Samoa or Samoan Islands or Navigator Island or Navigator Islands or
Sao Tome or Senegal or Serbia or Montenegro or Sierra Leone or Sri Lanka or Ceylon or
Solomon Islands or Somalia or South Africa or Sudan or South Sudan or Suriname or
Surinam or Swaziland or Eswatini or Syria or Syrian Arab Republic or Tajikistan or
Tadzhikistan or Tadjikistan or Tadzhik or Tanzania or Thailand or Togo or Togolese
Republic or Tonga or Tunisia or Turkey or Turkmenistan or Turkmen or Tuvalu or Uganda
or Ukraine or USSR or Soviet Union or Union of Soviet Socialist Republics or Uzbekistan or
Uzbek or Vanuatu or New Hebrides or Venezuela or Vietnam or Viet Nam or West Bank
or Yemen or Yugoslavia or Zambia or Zimbabwe or Rhodesia) OR AB (Afghanistan or
Albania or Algeria or American Samoa or Angola or Armenia or Armenian or Azerbaijan or
Bangladesh or Benin or Byelarus or Byelorussian or Belarus or Belorussian or Belorussia
or Belize or Bhutan or Bolivia or Bosnia or Herzegovina or Hercegovina or Botswana
or Brasil or Brazil or Bulgaria or Burkina Faso or Burkina Fasso or Upper Volta or Burundi
or Urundi or Cambodia or Khmer Republic or Kampuchea or Cameroon or Cameroons or
Cameron or Camerons or Cameroun or Cape Verde or Cabo Verde or Central African
Republic or Chad or China or Colombia or Comoros or Comoro Islands or Comores or
Mayotte or Congo or Zaire or Costa Rica or Cote d'Ivoire or Ivory Coast or Cuba or
Djibouti or French Somaliland or Dominica or Dominican Republic or East Timor or
East Timur or Timor Leste or Ecuador or Egypt or United Arab Republic or El Salvador or
Equatorial Guinea or Eritrea or Ethiopia or Fiji or Gabon or Gabonese Republic or Gambia
or Gaza or Georgia or Georgia Republic or Georgian Republic or Ghana or Gold Coast or
Grenada or Guatemala or Guinea or Bissau or Guiana or Guyana or Haiti or Honduras or
India or Maldives or Indonesia or Iran or Iraq or Jamaica or Jordan or Kazakhstan or
Kazakh or Kenya or Kiribati or Korea or Democratic People's Republic of Korea or Kosovo
or Kyrgyzstan or Kirghizia or Kyrgyz Republic or Kirghiz or Kirgizstan or Lao PDR or Laos or
Lebanon or Lesotho or Basutoland or Liberia or Libya or Macedonia or Madagascar or
Malagasy Republic or Malaysia or Malaya or Malay or Sabah or Sarawak or Malawi or
Nyasaland or Mali or Marshall Islands or Mauritania or Mauritius or Agalega Islands or
Mexico or Micronesia or Middle East or Moldova or Moldovia or Moldovian or Mongolia
or Montenegro or Morocco or Ifni or Mozambique or Myanmar or Myanma or Burma or
Namibia or Nauru or Nepal or Nicaragua or Niger or Nigeria or Pakistan or Papua New
Guinea or Palestine or Paraguay or Peru or Philippines
or Philipines or Phillipines or Phillippines or Romania or Rumania or Roumania or Russia
or Russian or Rwanda or Ruanda or Saint Lucia or St Lucia or Saint Vincent or St Vincent
or Grenadines or Samoa or Samoan Islands or Navigator Island or Navigator Islands or
Sao Tome or Senegal or Serbia or Montenegro or Sierra Leone or Sri Lanka or Ceylon or
Solomon Islands or Somalia or South Africa or Sudan or South Sudan or Suriname or
Surinam or Swaziland or Eswatini or Syria or Syrian Arab Republic or Tajikistan or
Tadzhikistan or Tadjikistan or Tadzhik or Tanzania or Thailand or Togo or Togolese
Republic or Tonga or Tunisia or Turkey or Turkmenistan or Turkmen or Tuvalu or Uganda
or Ukraine or USSR or Soviet Union or Union of Soviet Socialist Republics or Uzbekistan or
Uzbek or Vanuatu or New Hebrides or Venezuela or Vietnam or Viet Nam or West Bank
or Yemen or Yugoslavia or Zambia or Zimbabwe or Rhodesia) OR TI (Afghanistan or

Albania or Algeria or American Samoa or Angola or Armenia or Armenian or Azerbaijan or Bangladesh or Benin or Byelarus or Byelorussian or Belarus or Belorussian or Belorussia or Belize or Bhutan or Bolivia or Bosnia or Herzegovina or Hercegovina or Botswana or Brasil or Brazil or Bulgaria or Burkina Faso or Burkina Fasso or Upper Volta or Burundi or Urundi or Cambodia or Khmer Republic or Kampuchea or Cameroon or Cameroons or Cameron or Camerons or Cameroun or Cape Verde or Cabo Verde or Central African Republic or Chad or China or Colombia or Comoros or Comoro Islands or Comores or Mayotte or Congo or Zaire or Costa Rica or Cote d'Ivoire or Ivory Coast or Cuba or Djibouti or French Somaliland or Dominica or Dominican Republic or East Timor or East Timur or Timor Leste or Ecuador or Egypt or United Arab Republic or El Salvador or Equatorial Guinea or Eritrea or Ethiopia or Fiji or Gabon or Gabonese Republic or Gambia or Gaza or Georgia or Georgia Republic or Georgian Republic or Ghana or Gold Coast or Grenada or Guatemala or Guinea or Bissau or Guiana or Guyana or Haiti or Honduras or India or Maldives or Indonesia or Iran or Iraq or Jamaica or Jordan or Kazakhstan or Kazakh or Kenya or Kiribati or Korea or Democratic People's Republic of Korea or Kosovo or Kyrgyzstan or Kirghizia or Kyrgyz Republic or Kirghiz or Kirgizstan or Lao PDR or Laos or Lebanon or Lesotho or Basutoland or Liberia or Libya or Macedonia or Madagascar or Malagasy Republic or Malaysia or Malaya or Malay or Sabah or Sarawak or Malawi or Nyasaland or Mali or Marshall Islands or Mauritania or Mauritius or Agalega Islands or Mexico or Micronesia or Middle East or Moldova or Moldovia or Moldovian or Mongolia or Montenegro or Morocco or Ifni or Mozambique or Myanmar or Myanma or Burma or Namibia or Nauru or Nepal or Nicaragua or Niger or Nigeria or Pakistan or Papua New Guinea or Palestine or Paraguay or Peru or Philippines or Philipines or Phillipines or Phillippines or Romania or Rumania or Roumania or Russia or Russian or Rwanda or Ruanda or Saint Lucia or St Lucia or Saint Vincent or St Vincent or Grenadines or Samoa or Samoan Islands or Navigator Island or Navigator Islands or Sao Tome or Senegal or Serbia or Montenegro or Sierra Leone or Sri Lanka or Ceylon or Solomon Islands or Somalia or South Africa or Sudan or South Sudan or Suriname or Surinam or Swaziland or Eswatini or Syria or Syrian Arab Republic or Tajikistan or Tadzhikistan or Tadjikistan or Tadzhik or Tanzania or Thailand or Togo or Togolese Republic or Tonga or Tunisia or Turkey or Turkmenistan or Turkmen or Tuvalu or Uganda or Ukraine or USSR or Soviet Union or Union of Soviet Socialist Republics or Uzbekistan or Uzbek or Vanuatu or New Hebrides or Venezuela or Vietnam or Viet Nam or West Bank or Yemen or Yugoslavia or Zambia or Zimbabwe or Rhodesia)

16. AB ((developing or less* developed or under developed or underdeveloped or middle income or low* income or underserved or under served or deprived or poor*) N2 (countr* or nation? or population? or world)) OR TI ((developing or less* developed or under developed or underdeveloped or middle income or low* income or underserved or under served or deprived or poor*) N2 (countr* or nation? or population? or world))
17. AB ((developing or less* developed or under developed or underdeveloped or middle income or low* income) N2 (economy or economies)) OR TI ((developing or less* developed or under developed or underdeveloped or middle income or low* income) N2 (economy or economies))
18. AB (low* N2 (gdp or gnp or gross domestic or gross national)) OR TI (low* N2 (gdp or gnp or gross domestic or gross national))
19. AB (low N3 middle N3 (countr* or nation*)) OR TI (low N3 middle N3 (countr* or nation*))

20. AB (lmic or lmics or third world or lami countr*) OR TI (lmic or lmics or third world or lami countr*)
21. AB (transitional countr*) OR TI (transitional countr*)

**Measurement Concept**

22. MH (Health status indicators)
23. AB (indicator* or "health indicator" or evaluation or tool* or assessment or methodolog* or standards or "unmet need" or coverage) OR TI (indicator* or "health indicator" or evaluation or tool* or assessment or methodolog* or standards or "unmet need" or coverage)
24. AB (monitor* or surveillance or screening) OR TI (monitor* or surveillance or screening)
25. AB ("rapid counting" or "aerial surveillance" or "flow monitoring" or "enumeration" or "reproductive health assessment toolkit") OR TI ("rapid counting" or "aerial surveillance" or "flow monitoring" or "enumeration" or "reproductive health assessment toolkit")
26. (MH "Needs Assessment") OR (MH "Clinical Assessment Tools+") OR (MH "Clinical Indicators") OR (MH "Outcome Assessment") OR (MH "Process Assessment (Health Care)")
27. AB ((rapid or needs) N2 (assessment or evaluation)) OR TI ((rapid or needs) N2 (assessment or evaluation))
28. (MH "Data Collection+") OR (MH "Data Collection Methods+") OR (MH "Study Design+")
29. (MH "Questionnaires+") OR (MH "Research Instruments+") OR (MH "Instrument by Type+") OR (MH "Research Measurement+") or (MH “surveys”)
30. AB (scale* or survey* or questionnaire*) OR TI (scale* or survey* or questionnaire*)
31. AB (index or indices) OR TI (index or indices)
32. AB ("minimum initial service package" or MISP) OR TI ("minimum initial service package" or MISP)
33. AB (measure* or metric* or method*) OR TI (measure* or metric* or method*)
34. (MH "Health Status")
35. (MH "Health Knowledge") OR (MH "Attitude to Health")
    (MH "Health Care Delivery") OR (MH "Health Services Accessibility")
    (MH "Health Services Needs and Demand") OR (MH "Health and Welfare Planning")
    AB ((health N3 access*)) OR TI ((health N3 access*))
    (MH "Quality-Adjusted Life Years") OR (MH "Quality of Life") OR (MH "Disability-Adjusted Life Years")
    AB (outcome# N3 (health or measur* or assess* or (score or scoring) or index or indices or scale# or monitor#)) OR TI ((outcome# N3 (health or measur* or assess* or (score or scoring) or index or indices or scale# or monitor#))
    (MH "Health Services Research")
36. (MH "Morbidity") OR (MH "Mortality") OR (MH "Maternal Mortality") OR (MH "Prevalence") OR (MH "Incidence") OR (MH "Infant Mortality")
    AB (morbidity or mortality or incidence or prevalence) OR TI (morbidity or mortality or incidence or prevalence)

**SRHR Indicator Concept**

1. MH (Gynecology)

2. AB (gynaecology or gynecology) OR TI (gynaecology or gynecology)
3. MH (Reproductive Health) OR (MH "Sexual Health") OR (MH "Women's Health") OR (MH "Maternal-Child Health")
4. AB ("reproductive health" or "sexual health" or “reproductive services”) OR TI ("reproductive health" or "sexual health" or “reproductive services”)
5. MH (Sex Education) OR (MH “HIV Education”) OR (MH ”Childbirth Education”)
6. AB ("family planning education" or "family planning counselling" or "family planning instructor#" " or "family planning training" or "sex* education" or "sex* instruction" or (reproductive health N3 knowledge) or (HIV N3 knowledge) or (STI* N3 knowledge) or (sex* knowledge)) OR TI ("family planning education" or "family planning counselling" or "family planning instructor" or "family planning instructors" or "family planning training" or "sex education" or "sex instruction" or "sexual health education" or "sexual education" or "reproductive health knowledge" or "HIV knowledge" or "STI* knowledge" or "sexuality education")
7. AB ("minimum initial service package" or MISP) OR TI ("minimum initial service package" or MISP)
8. MH (Maternal health services)
9. MH (Maternal Welfare) OR (MH “Maternal-Child Welfare”)
10. AB ("maternal health" or "maternal welfare" or "maternal child health" or "maternal child welfare" or "maternal health services") or TI ("maternal health" or "maternal welfare" or "maternal child health" or "maternal child welfare" or "maternal health services")
11. MH(Obstetrics)
12. AB (obstetric*) or TI(obstetric*)
13. MH (Delivery, Obstetric) OR (MH “Obstetric Care”)
14. (MH "Obstetric Emergencies") OR (MH "Pregnancy Complications+")
15. AB ((labor or labour) N5 (birth* or breech or childbirth or childbirths or complicat* or difficult or early or easy or induce* or induction or late or obstetric* or onset or pregnan* or present*)) OR TI ((labor or labour) N5 (birth* or breech or childbirth or childbirths or complicat* or difficult or early or easy or induce* or induction or late or obstetric* or onset or pregnan* or present*))
16. MH (Pregnancy) OR (MH “Labor”) OR (MH “Childbirth”)
17. AB (pregnanc* or "child bearing" or childbearing) OR TI(pregnanc* or "child bearing" or childbearing)
18. MH ( Prenatal Care)
19. AB("prenatal" or "pre natal" or "antenatal" or "ante natal") OR TI("prenatal" or "pre natal" or "antenatal" or "ante natal")
20. MH (Perinatal Care) or (MH “Intrapartum Care”)
21. AB("perinatal care" or "peri natal care" or "perinatal care" or "peri natal care") OR TI ("perinatal care" or "peri natal care" or "perinatal care" or "peri natal care")
22. AB ("peripartum* period*" or "perinatal* period*" or "peri natal* period*")OR TI ("peripartum* period*" or "perinatal* period*" or "peri natal* period*")
23. AB (birth or childbirth* or parturition* or "safe delivery" or "safely delivered") OR TI (birth or childbirth* or parturition* or "safe delivery" or "safely delivered")
24. AB (antepartum or "ante partum" or intrapartum or "intra partum")OR TI (antepartum or "ante partum" or intrapartum or "intra partum")
25. (MH “Perinatal Death”)

26. AB (stillbirth or stillbirths or stillborn or stillborns or "still birth" or "still births" or "still born" or "still borns")OR TI (stillbirth or stillbirths or stillborn or stillborns or "still birth" or "still births" or "still born" or "still borns")
27. AB ("emergency obstetric care" OR "basic emergency obstetric care" OR "comprehensive emergency obstetric care" OR "basic emoc" OR "comprehensive emoc") OR TI ("emergency obstetric care" OR "basic emergency obstetric care" OR "comprehensive emergency obstetric care" OR "basic emoc" OR "comprehensive emoc")
28. AB (emoc or emonc or cemoc or bemoc) OR TI (emoc or emonc or cemoc or bemoc)
29. MH (Midwifery+) OR (MH "Midwives+")
30. AB ("birth attendant" or "birth attendants" or midwife or midwives or midwifery or "traditional birth attendant*" or "skilled birth attendant*") OR TI ("birth attendant" or "birth attendants" or midwife or midwives or midwifery or "traditional birth attendant*" or "skilled birth attendant*")
31. MH (Postnatal Care) OR (MH “Postnatal Period”)
32. AB (postnatal or "post natal" or postpartum or "post partum" or puerperium or puerperal) OR TI (postnatal or "post natal" or postpartum or "post partum" or puerperium or puerperal)
33. (MH "Abortion, Incomplete") OR (MH "Abortion, Spontaneous") OR (MH "Abortion, Habitual") OR (MH "Abortion, Criminal") OR (MH "Abortion, Induced")
34. AB (miscarriage*) OR TI (miscarriage*)
35. AB (abortion or abortions or aborted or aborting or abortus) OR TI (abortion or abortions or aborted or aborting or abortus)
36. AB ((unwanted or unintended) N3(pregnanc* )) OR TI((unwanted or unintended) N3 (pregnanc*))
37. AB (Abortion N3 (safe or unsafe or post or care)) OR TI (Abortion N3 (safe or unsafe or post or care))
38. MH (Misoprostol)
39. (MH "Abortifacient Agents") OR (MH "Mifepristone") AB (Misoprostol or mifepristone or cytotec or mifeprex or mifegymiso or "abortion pill") OR TI (Misoprostol or mifepristone or cytotec or mifeprex or mifegymiso or "abortion pill")
40. MH (Birth Intervals)
41. MH (family planning) OR (MH "Family Planning, Natural")
42. AB ("birth interval" or "birth intervals" or "birth spacing" or "birth spacings" or "child spacing" or "child spacings" or "family building" or "family planning" or "pregnancy interval" or "pregnancy intervals") OR TI ("birth interval" or "birth intervals" or "birth spacing" or "birth spacings" or "child spacing" or "child spacings" or "family building" or "family planning" or "pregnancy interval" or "pregnancy intervals")
43. AB ("safe motherhood") OR TI("safe motherhood")
44. (MH "Contraception") OR (MH "Contraceptive Agents, Male") OR (MH "Contraceptives, Postcoital") OR (MH "Contraceptive Agents") OR (MH "Contraceptives, Oral Combined") OR (MH "Contraceptive Devices")
45. AB ("birth regulation" or "birth control*" or "coitus interruptus" or "fertility control" or contracept* or "conception control" or antifertility or anticonception or "fertility control" or "fertili?ation inhibition" or "inhibition of fertili?ation" or "fertili?ation inhibition" or "inhibition of fertili?ation" or "pregnancy prevent*") OR TI ("birth regulation" or "birth control*" or "coitus interruptus" or "fertility control" or contracept* or "conception control" or antifertility or

anticonception or "fertility control" or "fertili?ation inhibition" or "inhibition of fertili?ation" or "fertili?ation inhibition" or "inhibition of fertili?ation" or "pregnancy prevent*")

46. MH (Contraceptive Devices) OR (MH "Intrauterine Devices") OR (MH "Diaphragms, Contraceptive") OR (MH "Condoms") OR (MH "Cervical Caps")
47. AB ("cervical cap" or "cervical caps" or "i.u.d." or "intra uterine device*" or "intracervical device" or "intrauterine device*" or "iucd" or "iud" or "iuds" or "progestin implant" or "vaginal diaphragm" or "vaginal diaphragms" or "vaginal ring" or "vaginal shield" or "vaginal rings" or "vaginal shields" or "vaginal sponge" or "vaginal sponges" or "coiled spring*" or condom or condoms or spermicide or "rhythm method" or "calendar method" or "pull out method" or "tubal ligation" or vasectomy or "depo provera") OR TI ("cervical cap" or "cervical caps" or "i.u.d." or "intra uterine device*" or "intracervical device" or "intrauterine device*" or "iucd" or "iud" or "iuds" or "progestin implant" or "vaginal diaphragm" or "vaginal diaphragms" or "vaginal ring" or "vaginal shield" or "vaginal rings" or "vaginal shields" or "vaginal sponge" or "vaginal sponges" or "coiled spring*" or condom or condoms or spermicide or "rhythm method" or "calendar method" or "pull out method" or "tubal ligation" or vasectomy or "depo provera")
48. AB (("birth control" or contracept*) N3 (pill or patch or implant or shot or injection or withdrawal)) OR TI (("birth control" or contracept*) N3 (pill or patch or implant or shot or injection or withdrawal))
49. MH (Contraceptives, Postcoital)
50. AB ("morning after pill" or "postcoital antifertility agent" or "postcoital pill" or "plan B" or “emergency contraception”) or TI ("morning after pill" or "postcoital antifertility agent" or "postcoital pill" or "plan B" or “emergency contraception”)
51. MH (Sexual Abstinence)
52. AB (celibacy or "postpartum abstinence" or "sexual abstinence" or virginity) OR TI (celibacy or "postpartum abstinence" or "sexual abstinence" or virginity)
53. MH (hiv/ or hiv-1/ or hiv-2/)
54. MH (Human immunodeficiency virus)
55. AB ("acquired immune deficiency syndrome virus" or "acquired immunodeficiency syndrome virus" or "aids associated lentivirus" or "aids associated retrovirus" or "aids associated virus" or "aids related virus" or "aids virus" or "aids viruses" or "hiv" or "human immuno deficiency virus" or "human immunodeficiency virus" or "human immunodeficiency viruses" or "aids related illness*") OR TI("acquired immune deficiency syndrome virus" or "acquired immunodeficiency syndrome virus" or "aids associated lentivirus" or "aids associated retrovirus" or "aids associated virus" or "aids related virus" or "aids virus" or "aids viruses" or "hiv" or "human immuno deficiency virus" or "human immunodeficiency virus" or "human immunodeficiency viruses" or "aids related illness*")
56. MH (HIV Infections+)
57. AB ("hiv infection*" or "hiv seropositivit*" or "hiv coinfection*") OR TI ("hiv infection*" or "hiv seropositivit*" or "hiv coinfection*")
58. MH (Acquired Immunodeficiency Syndrome)
59. AB ("aids" or "acquired immune deficiency syndrome*" or "acquired immuno deficiency syndrome*" or "acquired immunodeficiency syndrome*" or "aids related opportunistic infections”) OR TI ("aids" or "acquired immune deficiency syndrome*" or "acquired immuno

deficiency syndrome*" or "acquired immunodeficiency syndrome*" or "aids related opportunistic infections”)

60. (MH "Disease Transmission, Vertical")  (MH "Postexposure Follow-Up")
61. AB ("post exposure prophylaxis" or PEP) OR TI ("post exposure prophylaxis" or PEP)
62. MH (Antiretroviral Therapy, Highly Active)
63. MH (Anti-HIV agents)
64. AB ("Antiretroviral therapy" or "antiretroviral treatment" or ART or ARV or antiretroviral* or HAART or "anti hiv agent*") OR TI ("Antiretroviral therapy" or "antiretroviral treatment" or ART or ARV or antiretroviral* or HAART or "anti hiv agent*")
65. AB (pmtct or "prevention of HIV mother to child transmission" or "prevention of mother to child transmission" or "eliminate mother to child transmission" or "prevention of mother to child HIV transmission") OR TI (pmtct or "prevention of HIV mother to child transmission" or "prevention of mother to child transmission" or "eliminate mother to child transmission" or "prevention of mother to child HIV transmission")
66. MH (Sexually Transmitted Diseases)
67. AB ("sexually transmitted disease*" or "sexually transmitted infection*" or "venereal disease*" or "venereal infection" or "venereal infections" or STIs or STDs) OR TI ("sexually transmitted disease*" or "sexually transmitted infection*" or "venereal disease*" or "venereal infection" or "venereal infections" or STIs or STDs)
68. MH (sexually transmitted diseases, bacterial/ or chancroid/ or chlamydia infections/ or chlamydia trachomatis/ or gonorrhea/ or syphilis/ or Trichomonas Vaginitis/
(MH "Sexually Transmitted Diseases, Bacterial+") OR (MH "Chlamydia Infections") OR (MH "Sexually Transmitted Diseases, Fungal") OR (MH "Sexually Transmitted Diseases, Protozoal+")
69. AB ("chancroid" or "chancroids" or chlamidiosis or "chlamydia infection*" or "chlamydia trachomatis" or "gonorrhea" or "granuloma inguinale" or "granuloma venereum" or "Klebsiella granulomatis infection" or "Neisseria gonorrhoeae infection" or "syphilis" or "syphilitic disorder or trichomoniasis or "trichomonas vaginitis") OR TI ("chancroid" or "chancroids" or chlamidiosis or "chlamydia infection*" or "chlamydia trachomatis" or "gonorrhea" or "granuloma inguinale" or "granuloma venereum" or "Klebsiella granulomatis infection" or "Neisseria gonorrhoeae infection" or "syphilis" or "syphilitic disorder or trichomoniasis or "trichomonas vaginitis")
70. (MH "Sexually Transmitted Diseases, Viral") OR (MH "Warts, Venereal") OR (MH "Herpes Genitalis")  (MH "Hepatitis B+")
71. MH (Papillomavirus Infections)
72. AB ("anal wart" or "anal warts" or "anogenital wart" or "anogenital warts" or "condyla acuminatum" or "condylatum acuminatum" or "condyloma accuminatum" or "condyloma acuminata" or "condylomata acuminata" or "genital herpes" or "genital wart" or "genital warts" or "herpes genitalis" or "herpes progenitalis" or "herpes simplex genitalis" or "herpes simplex virus genital infection" or "penile wart" or "penile warts" or "perianal wart" or "perianal warts" or "venereal wart" or "venereal warts"or "human papillomavirus*" or HPV or "papillomavirus infection*" or "hepatitis B") OR TI ("anal wart" or "anal warts" or "anogenital wart" or "anogenital warts" or "condyla acuminatum" or "condylatum acuminatum" or "condyloma accuminatum" or "condyloma acuminata" or "condylomata acuminata" or "genital herpes" or "genital wart" or "genital warts" or "herpes genitalis" or "herpes progenitalis" or "herpes simplex genitalis" or "herpes simplex virus genital infection" or "penile wart" or "penile warts"

or "perianal wart" or "perianal warts" or "venereal wart" or "venereal warts"or "human papillomavirus*" or HPV or "papillomavirus infection*" or "hepatitis B")

73. MH (Scabies/)
74. AB ("sarcoptic mange" or scabies) OR TI ("sarcoptic mange" or scabies)
75. (MH "Lice Infestations")
76. AB ("crab lice" or "crab lices" or "crab louse" or "crab louses" or "Pediculus pubis" or "phthirus" or "Phtirus pubis" or "Pthirus pubis" or "pubic lice" or "pubic louse") OR TI ("crab lice" or "crab lices" or "crab louse" or "crab louses" or "Pediculus pubis" or "phthirus" or "Phtirus pubis" or "Pthirus pubis" or "pubic lice" or "pubic louse")
77. MH (intimate partner violence) OR (MH “Domestic Violence”) OR (MH “Gender-Based Violence”)
78. AB ("partner abuse" or "partner violence" or "wife abuse" or "spouse abuse" or "spousal abuse" or "domestic violence" or "domestic abuse" or "gender based violence" or "gender-based violence" or "GBV" or "IPV" or "sex* based violence") OR TI ("partner abuse" or "partner violence" or "wife abuse" or "spouse abuse" or "spousal abuse" or "domestic violence" or "domestic abuse" or "gender based violence" or "gender-based violence" or "GBV" or "IPV" or "sex* based violence")
79. AB ((abuse* or assault* or violence) N2 (woman or women)) OR TI ((abuse* or assault* or violence) N2 (woman or women))
80. (MH "Sexual Abuse+") OR (MH "Rape") OR (MH "Child Abuse, Sexual") OR (MH "Human Trafficking") OR (MH “Circumcision, Female”)
81. AB ("coerced intercourse" or "forced intercourse" or "forced prostitution" or "forced sex" or "human trafficking" or "human traffickings" or rape or "sex trafficking" or "sex traffickings" or "sex* abuse*" or "sex* assault*" or "sex* crime*" or "sex* offense" or "sex* offenses" or "sex* slave*" or "sexual aggression" or "sexual bullying" or "sexual coercion" or "sexual exploitation*" or "sexual harassment" or "sexual trauma" or "sexual violence" or "unwanted sex" or "unlawful sex" or "honor killings" or “transactional sex”)OR TI ("coerced intercourse" or "forced intercourse" or "forced prostitution" or "forced sex" or "human trafficking" or "human traffickings" or rape or "sex trafficking" or "sex traffickings" or "sex* abuse*" or "sex* assault*" or "sex* crime*" or "sex* offense" or "sex* offenses" or "sex* slave*" or "sexual aggression" or "sexual bullying" or "sexual coercion" or "sexual exploitation*" or "sexual harassment" or "sexual trauma" or "sexual violence" or "unwanted sex" or "unlawful sex" or "honor killings" or “transactional sex”)
82. AB ("physical* abuse*" or "physical* assault*" or "physical violence" or "psychological violence" or "emotional violence" or "economic violence" or "female circumcision" or "female genital mutilation" or "female genital cutting" or FGM) OR TI ("physical* abuse*" or "physical* assault*" or "physical violence" or "psychological violence" or "emotional violence" or "economic violence" or "female circumcision" or "female genital mutilation" or "female genital cutting" or FGM)
83. MH (fistula or vaginal fistula)
84. AB (fistula or fistulas or "genital trauma" or "genital injury" or "vaginal trauma" or "vaginal injury") OR TI (fistula or fistulas or "genital trauma" or "genital injury" or "vaginal trauma" or "vaginal injury")

85. MH (pregnancy complications/ or abortion, spontaneous/ or diabetes, gestational/ or hypertension, pregnancy-induced/ or obstetric labor complications/
86. (MH "Pre-Eclampsia") OR (MH "Pregnancy-Induced Hypertension") OR (MH "Eclampsia") AB ("toxaemia" or " toxemia" or "hypertension edema proteinuria gestosis" or "pre eclampsia" or "pre eclamptic toxaemia" or "pre eclamptic toxemia" or "preeclamptic toxaemia" or "preeclamptic toxemia" or "pregnancy toxaemia*" or "pregnancy toxemia*" or "toxemia of pregnanc*" or "toxemic pregnancy" or eclampsia or preeclampsia) OR TI("toxaemia" or " toxemia" or "hypertension edema proteinuria gestosis" or "pre eclampsia" or "pre eclamptic toxaemia" or "pre eclamptic toxemia" or "preeclamptic toxaemia" or "preeclamptic toxemia" or "pregnancy toxaemia*" or "pregnancy toxemia*" or "toxemia of pregnanc*" or "toxemic pregnancy" or eclampsia or preeclampsia)
87. MH (dystocia)
88. AB ("abnormal labor" or "abnormal labour" or "delayed labor" or "delayed labour" or "inertia uteri" or "labor obstruction" or "labour obstruction" or "obstructed labor" or "obstructed labour" or "uterus inertia" or dystocia or dystocias) OR TI ("abnormal labor" or "abnormal labour" or "delayed labor" or "delayed labour" or "inertia uteri" or "labor obstruction" or "labour obstruction" or "obstructed labor" or "obstructed labour" or "uterus inertia" or dystocia or dystocias)
89. MH (Breech Presentation)
90. AB (breech N2 (present* or position*)) OR TI (breech N2 (present* or position*))
91. MH (Uterine Hemorrhage)
92. MH (Postpartum hemorrhage)
93. AB ("vagina* haemorrhage" or "vagina* hemorrhage" or "vaginal bleeding") OR TI ("vagina* haemorrhage" or "vagina* hemorrhage" or "vaginal bleeding")
94. MH (Menstruation) OR (MH “Menstrual Cycle”)
95. AB ((Menstruation or menarche or menstrual or menses) N2 (hygiene or bleeding or cycle)) OR TI ((Menstruation or menarche or menstrual or menses) N2 (hygiene or bleeding or cycle))
96. MH (women’s rights)
97. AB ((sexual or reproductive or women’s) N3 (rights)) OR TI ((sexual or reproductive or women’s) N3 (rights))
98. AB ("Sexual initiation" or "early sexual debut") OR TI ("Sexual initiation" or "early sexual debut")

**<u>Database(s): Embase</u>**

**Search Strategy:**

1. Gynecology/

2. (gynaecology or gynecology).tw,kw.

3. Reproductive Health/

Sexual health/

4. ("reproductive health" or "sexual health" or "reproductive services").tw,kw.

5. Sexual Education/

6. ("family planning education" or "family planning counselling" or "family planning instructor*" or "family planning training" or "sex* education" or "sex* instruction" or (reproductive health adj3 knowledge) or (HIV adj3 knowledge) or (STI adj3 knowledge) or (sex* adj3 knowledge)).tw,kw.

7. ("minimum initial service package" or MISP).tw,kw.

8. Maternal Health/

9. Maternal health service/

10. Maternal Welfare/ or maternal care/

11. ("maternal health" or "maternal welfare" or "maternal child health" or "maternal child welfare" or "maternal health services" or "maternal care").tw,kw.

12. Obstetrics/

13. obstetric*.tw,kw.

14. exp Delivery, Obstetric/ Labor,obstetric/

15. ((labor or labour) adj5 (birth* or breech or childbirth or childbirths or complicat* or difficult or early or easy or induce* or induction or late or obstetric* or onset or pregnan* or present*)).tw,kw.

16. pregnancy/ or adolescent pregnancy/ or unplanned pregnancy/ or unwanted pregnancy/

17. (pregnanc* or "child bearing" or childbearing).tw,kw.

18. Prenatal Care/

19. ("prenatal" or "pre natal" or "antenatal" or "ante natal").tw,kw.

20. exp Perinatal Care/

21. ("perinatal care" or "peri natal care" or "perinatal care" or "peri natal care").tw,kw.

22. Perinatal period/

23. ("peripartum* period*" or "perinatal* period*" or "peri natal* period*").tw,kw.

24. exp Birth/

25. (birth or childbirth* or parturition* or "safe delivery" or "safely delivered").tw,kw.

26. (antepartum or "ante partum" or intrapartum or "intra partum").tw,kw.

27. exp Stillbirth/

28. (stillbirth or stillbirths or stillborn or stillborns or "still birth" or "still births" or "still born" or "still borns").tw,kw.

29. ("emergency obstetric care" or "basic emergency obstetric care" or "comprehensive emergency obstetric care" or "basic emoc" or "comprehensive emoc").tw,kw.

30. (emoc or emonc or cemoc or bemoc).tw,kw.

31. Midwife/

32. ("birth attendant" or "birth attendants" or midwife or midwives or midwifery or "traditional birth attendant*" or "skilled birth attendant*").tw,kw.

33. Postnatal Care/

Puerperium/

34. Postpartum Period/

35. (postnatal or "post natal" or postpartum or "post partum" or puerperium or puerperal).tw,kw.

36. Abortion/  or septic abortion/ or spontaneous abortion/

37. miscarriage*.tw,kw.

38. abortion, induced/ or abortion, eugenic/ or abortion, legal/ or abortion, therapeutic/

39. exp induced abortion/ or exp illegal abortion/ or exp incomplete abortion/ or exp legal abortion/ or exp selective abortion/ or exp therapeutic abortion/ or exp medical abortion/

40. (abortion or abortions or aborted or aborting or abortus).tw,kw.

41. (unwanted or unintended pregnanc*).tw,kw.

42. (Abortion adj3 (safe or unsafe or post)).tw,kw.

43. Misoprostol/

44. exp mifepristone plus misoprostol/ or exp mifepristone/ or exp induced abortion/ or exp misoprostol/

45. (Misoprostol or mifepristone or cytotec or mifeprex or mifegymiso or "abortion pill").tw,kw.

47. family planning/

48. ("birth interval" or "birth intervals" or "birth spacing" or "birth spacings" or "child spacing" or "child spacings" or "family building" or "family planning" or "pregnancy interval" or "pregnancy intervals").tw,kw.

49. "safe motherhood".tw,kw.

50. Birth control/ exp contraception/ or barrier contraception/ or emergency contraception/ or hormonal contraception/ or long-lasting reversible contraception/ or oral contraception/ or ovulation inhibition/ or reproductive sterilization/ or vaginal contraception/ coitus interruptus/

51. ("birth regulation" or "birth control*" or "coitus interruptus" or "fertility control" or contracept* or "conception control" or antifertility or anticonception or "fertility control" or "fertilization inhibition" or "inhibition of fertilization" or "fertilisation inhibition" or "inhibition of fertili?ation" or "pregnancy prevent*").tw,kw.

52. exp Contraceptive Device/ or female contraceptive device/ or birth control implant/ or contraceptive patch/ or contraceptive spong/ or female condom/ or intrauterine contraceptive device/ or tubal occlusion device/ or uterine cervix cap/ or vagina pessary/ or vagina ring/ male contraceptive device/ or condom/

53. ("cervical cap" or "cervical caps" or "i.u.d." or "intra uterine device*" or "intracervical device" or "intrauterine device*" or "iucd" or "iud" or "iuds" or "progestin implant" or "vaginal diaphragm" or "vaginal diaphragms" or "vaginal ring" or "vaginal shield" or "vaginal rings" or "vaginal shields" or "vaginal sponge" or "vaginal sponges" or "coiled spring*" or condom or condoms or spermicide or "rhythm method" or "calendar method" or "pull out method" or "tubal ligation" or vasectomy or "depo provera").tw,kw.

oral contraceptive agent/

54. (("birth control" or contracept*) adj3 (pill or patch or implant or shot or injection or withdrawal)).tw,kw.

55. Contraceptives, Postcoital/ or postcoitus contraceptive agent/

56. ("morning after pill" or "postcoital antifertility agent" or "postcoital pill" or "plan B" or "emergency contraception" or "emergency contraceptive pill" or "postcoital contraceptives").tw,kw.

57. Sexual Abstinence/

58. (celibacy or "postpartum abstinence" or "sexual abstinence" or virginity).tw,kw.

60. human immunodeficiency virus/ or human immunodeficiency virus 1/ or human immunodeficiency virus 2/

61. ("acquired immune deficiency syndrome virus" or "acquired immunodeficiency syndrome virus" or "aids associated lentivirus" or "aids associated retrovirus" or "aids associated virus" or "aids related virus" or "aids virus" or "aids viruses" or "hiv" or "human immuno deficiency virus" or "human immunodeficiency virus" or "human immunodeficiency viruses" or "aids related illness*").tw,kw.

62. Human immunodeficiency virus infection/

63. ("hiv infection*" or "hiv seropositivit*" or "hiv coinfection*").tw,kw.

64. Acquired Immunodeficiency Syndrome/

Opportunistic infection/

65. ("aids" or "acquired immune deficiency syndrome*" or "acquired immuno deficiency syndrome*" or "acquired immunodeficiency syndrome*" or "aids related opportunistic infection*").tw,kw.

66. vertical transmission/

69. Post exposure prophylaxis/

70. ("post exposure prophylaxis" or PEP).tw,kw.

71. exp antiretroviral therapy/ or highly active antiretroviral therapy/

72. anti human immunodeficiency virus agent/

73. ("Antiretroviral therapy" or "antiretroviral treatment" or ART or ARV or antiretroviral* or HAART or "anti hiv agent*").tw,kw.

74. (pmtct or "prevention of HIV mother to child transmission" or "prevention of mother to child transmission" or "eliminate mother to child transmission" or "prevention of mother to child HIV transmission").tw,kw.

- (mother adj2 child transmission).tw,kw.

75. Sexually Transmitted Disease/

76. ("sexually transmitted disease*" or "sexually transmitted infection*" or "venereal disease*" or "venereal infection" or "venereal infections" or STIs or STDs).tw,kw.

77. sexually transmitted diseases/ or condyloma acuminatum/ or genital herpes/ or gonorrhea/ or granuloma inguinale/ or lymphogranuloma venereum/ or secondary syphilis/ or syphilis/ or tables dorsalis/ or ulcus molle/ chlamydia trachomatis/ or chlamydia/ or chlamydiasis/ or vaginal trichomoniasis/

78. ("chancroid" or "chancroids" or chlamidiosis or "chlamydia infection*" or "chlamydia trachomatis" or "gonorrhea" or "granuloma inguinale" or "granuloma venereum" or "Klebsiella granulomatis infection" or "Neisseria gonorrhoeae infection" or "syphilis" or "syphilitic disorder or trichomoniasis or trichomonas vaginitis").tw,kw.

80. Hepatitis B/

Wart virus/ or papillomavirus infection/

81. ("anal wart" or "anal warts" or "anogenital wart" or "anogenital warts" or "condyla acuminatum" or "condylatum acuminatum" or "condyloma accuminatum" or "condyloma acuminata" or "condylomata acuminata" or "genital herpes" or "genital wart" or "genital warts" or

"herpes genitalis" or "herpes progenitalis" or "herpes simplex genitalis" or "herpes simplex virus genital infection" or "penile wart" or "penile warts" or "perianal wart" or "perianal warts" or "venereal wart" or "venereal warts" or "human papillomavirus*" or HPV or "papillomavirus infection*" or "hepatitis B").tw,kw.

82. Scabies/

83. ("sarcoptic mange" or scabies).tw,kw.

84. Phthirus/

85. ("crab lice" or "crab lices" or "crab louse" or "crab louses" or "Pediculus pubis" or "phthirus" or "Phtirus pubis" or "Pthirus pubis" or "pubic lice" or "pubic louse").tw,kw.

86. partner violence/ or domestic violence/ or marital rape/ or gender based violence/

88. ("partner abuse" or "partner violence" or "wife abuse" or "spouse abuse" or "spousal abuse" or "domestic violence" or "domestic abuse" or "gender based violence" or "gender-based violence" or "GBV" or "IPV" or "sex* based violence").tw,kw.

89. ((abuse* or assault* or violence) adj2 (woman or women)).tw,kw.

Sexual crime/ or sexual assault/ or rape/ or sexual abuse/ human trafficking/ or sex trafficking/ female genital mutilation/ or female genital mutilation type i/ or female genital mutilation type ii/ or female genital mutilation type iii/ or female genital mutilation type iv/ or reinfibulation/

91. ("coerced intercourse" or "forced intercourse" or "forced prostitution" or "forced sex" or "human trafficking" or "human traffickings" or rape or "sex trafficking" or "sex traffickings" or "sex* abuse*" or "sex* assault*" or "sex* crime*" or "sex* offense" or "sex* offenses" or "sex* slave*" or "sexual aggression" or "sexual bullying" or "sexual coercion" or "sexual exploitation*" or "sexual harassment" or "sexual trauma" or "sexual violence" or "unwanted sex" or "unlawful sex" or "honor killings").tw,kw.

violence/ or physical violence/

92. ("physical* abuse*" or "physical* assault*" or "physical violence" or "psychological violence" or "emotional violence" or "economic violence" or "female circumcision" or "female genital mutilation" or "female genital cutting" or FGM).tw,kw.
fistula/ or urinary tract fistula/

93. rectovaginal fistula/ or vagina disease/ or cystovaginal fistula/

94. (fistula or fistulas or "genital trauma" or "genital injur*" or "vaginal trauma" or "vaginal injur*").tw,kw.

95. utor hypertension, pregnancy-induced/ or obstetric labor complications/ pregnancy complication/ or intrauterine infection/ or maternal hypertension/ or pregnancy diabetes mellitus/

exp labor complication/ or exp cephalopelvic disproportion/ or exp dystocia/ or exp "immature and premature labor"/ or exp malpresentation/ or exp obstetric hemorrhage/ or exp placenta accreta/

or exp placenta previa/ or exp solutio placentae/ or exp uterine complication/ or exp vasa previa/ or breech presentation/

96. “eclampsia and preeclampsia"/ or pregnancy toxemia/ or eclampsia/ or preeclampsia/

97. ("toxaemia" or "toxemia" or "hypertension edema proteinuria gestosis" or "pre eclampsia" or "pre eclamptic toxaemia" or "pre eclamptic toxemia" or "preeclamptic toxaemia" or "preeclamptic toxemia" or "pregnancy toxaemia*" or "pregnancy toxemia*" or toxemia of pregnanc* or toxemic pregnancy or eclampsia or preeclampsia).tw,kw.

99. ("abnormal labor" or "abnormal labour" or "delayed labor" or "delayed labour" or "inertia uteri" or "labor obstruction" or "labour obstruction" or "obstructed labor" or "obstructed labour" or "uterus inertia" or dystocia or dystocias).tw,kw.

102. Postpartum hemorrhage/

103. ("vagina* haemorrhage" or "vagina* hemorrhage" or "vaginal bleeding" or "postpartum hemorrhage" or "post partum hemorrhage" ).tw,kw.

104. Menstruation/ or menstrual cycle/

105. ((Menstruation or menarche or menstrual or menses) adj2 (hygiene or bleeding or cycle)).tw,kw.

106. Human rights/ or Reproductive rights/ or women’s rights/

108. ("Sexual initiation" or "early sexual debut").tw,kw.

**Humanitarian Concept**

109. disaster/ or mass disaster/ or natural disaster/ or emergency/

Disaster planning/

110. exp disaster victim/

111. ((disaster or emergenc*) adj2 victim*).tw,kw.

112. ((disaster or disasters or catastrophe or catastrophes) adj5 (environ* or human or manmade or "man made" or nature or natural or weather)).tw,kw.

113. ("mass casualty" or "mass casualties" or "mass fatalities" or "mass fatality").tw,kw.

114. ((crisis or crises) adj5 (environ* or human or manmade or "man made" or nature or natural or weather or setting*)).tw,kw.

115. ((crisis or crises or conflict) adj3 affected).tw,kw.

116. "warfare and armed conflicts"/ or armed conflict*/ or warfare/ or warfare/ or ethnic cleansing/ or genocide/ or holocaust/ or war exposure/

Exp warfare/ or military phenomena/

war/ or war crime/ or war exposure/

violence/ or ethnic cleansing/ or ethnic conflict/ or genocide/

117. ("warfare and armed conflict*" or warfare or "war crime*" or "ethnic cleansing*" or "gas poisoning" or genocide or holocaust or "war exposure").tw,kw.

119. ("afghan campaign" or "armed conflict" or "armed conflicts" or "gulf war" or "iraq war" or "war time" or "wartime" or "war torn" or "war affected" or "insurgency" or "intra conflict").tw,kw.

120. ((armed or zone or political or civil or setting*) adj3 (conflict or conflicts or attack or attacks or war or wars or "no fly")).tw,kw.

121. (Fragile adj2 (state* or countr* or nation* or situation* or setting*)).tw,kw.

122. ("Post conflict" or "postconflict" or "post war" or "post conflict setting" or "peacebuilding" or peacekeeping).tw,kw.

(war adj2 related).tw,kw.

124. ("militant group?" or "militant organi?ation*" or militia or combatant or rebel*).tw,kw.

125. disaster medicine/ or emergency medicine/

127. Epidemic/

128. ("disaster medicine" or "disaster outbreak*" or "epidemic*" or "disease outbreak?" ).tw,kw.

129. emergency health service/

130. ((emergenc*) adj5 (environ* or human or manmade or "man made" or nature or natural or weather or complex)).tw,kw.

131. Starvation/

132. (famine or famines or starvation or starvations).tw,kw.

133 drought/ or flooding/ or tsunami/ or hurricane/ or tornado/ avalanche/ or earthquake/ or landslide/ or tsunami/ or volcanic ash/ or volcano/

134. (avalanche or avalanches or cyclone or cyclones or drought or droughts or earthquake or earthquakes or flood or flooded or flooding or floods or hurricane or hurricanes or landslide or landslides or "land slide" or "land slides" or mudslide or mudslides or "mud slide" or "mud slides" or storm or storms or tornado or tornadoes or tsunami or tsunamis or typhoon or typhoons or "volcanic ash" or "volcanic eruption" or "volcanic eruptions" or "volcanic gases" or rubble).tw,kw.

135. refugees/

136. (evacuee or evacuees or refugee or refugees or squatter or squatters or transients or "asylum seeker").tw,kw.

137. relief work/ or rescue work/

138. ((rescue or relief or aid) adj3 (plan or plans or activity or activities or agency or agencies)).tw,kw.

139. ("aid plan" or "aid work" or "relief plan" or "relief work" or "rescue plan" or "rescue work").tw,kw.

140. ((staff or staffs or worker or workers) adj3 (relief or aid)).tw,kw.

141. (humanitarian assistance or humanitarian assistances or relief work or relief works).tw,kw.

142. (humanitarian adj3 (aid or response or relief or crisis or crises or emergency or emergencies or disaster or disasters)).tw,kw.

143. Altruism/

144. (humanitarianism or altruism).tw,kw.

145. ("displaced children" or "displaced child" or "displaced families" or "displaced family" or "displaced individuals" or "displaced internally" or "displaced men" or "displaced people" or "displaced peoples" or "displaced person" or "displaced persons" or "displaced population" or "displaced populations" or "displaced women" or "displaced adolescent" or "displaced adolescents" or "forced displacement" or "forced displacements" or "forcibl* displace*" or "internal displaced" or "internal displacement" or "internally displaced" or "population displaced" or "population displacement" or "forced migration" or "migrant*").tw,kw.

146. (protected village).tw,kw.

147. ((camp or camps) adj3 (refugee or transit or displace* or temporary or informal)).tw,kw.

148. (Settlement adj3 (temporary or informal)).tw,kw.

**Population Concept**

150. ((displace* or refugee*) adj3 (child* or famil* or men or wom* or individual* or adolescent* or people* or population* or person* or girl* or boy* or youth)).tw,kw.

151. adolescent/ or young adult/

152. (adolescen* or teen* or youth or youths or "young adult" or "young adults" or girl* or boy* or "young wom*" or "young girl*" or "young boy*").tw,kw.

153. pregnant woman/

Exp mother/

154. (mother?).tw,kw.

155. female/

156. parturients.tw,kw.

157. "sexually active".tw,kw.

158. refugees/

159. (refugee or refugees).tw,kw.

160. (women adj2 reproductive adj2 age).tw,kw.

**Low and middle-income countries concept**

161. Developing Country.sh.

162. (Africa or Asia or Caribbean or West Indies or South America or Latin America or Central America).hw,kw,ti,ab,cp.

163. (Afghanistan or Albania or Algeria or American Samoa or Angola or Armenia or Armenian or Azerbaijan or Bangladesh or Benin or Byelarus or Byelorussian or Belarus or Belorussian or Belorussia or Belize or Bhutan or Bolivia or Bosnia or Herzegovina or Hercegovina or Botswana or Brasil or Brazil or Bulgaria or Burkina Faso or Burkina Fasso or Upper Volta or Burundi or Urundi or Cambodia or Khmer Republic or Kampuchea or Cameroon or Cameroons or Cameron or Camerons or Cameroun or Cape Verde or Cabo Verde or Central African Republic or Chad or China or Colombia or Comoros or Comoro Islands or Comores or Mayotte or Congo or Zaire or Costa Rica or Cote d'Ivoire or Ivory Coast or Cuba or Djibouti or French Somaliland or Dominica or Dominican Republic or East Timor or East Timur or Timor Leste or Ecuador or Egypt or United Arab Republic or El Salvador or Equatorial Guinea or Eritrea or Ethiopia or Fiji or Gabon or Gabonese Republic or Gambia or Gaza or Georgia or Georgia Republic or Georgian Republic or Ghana or Gold Coast or Grenada or Guatemala or Guinea or Bissau or Guiana or Guyana or Haiti or Honduras or India or Maldives or Indonesia or Iran or Iraq or Jamaica or Jordan or Kazakhstan or Kazakh or Kenya or Kiribati or Korea or Democratic People's Republic of Korea or Kosovo or Kyrgyzstan or Kirghizia or Kyrgyz Republic or Kirghiz or Kirgizstan or Lao PDR or Laos or Lebanon or Lesotho or Basutoland or Liberia or Libya or Macedonia or Madagascar or Malagasy Republic or Malaysia or Malaya or Malay or Sabah or Sarawak or Malawi or Nyasaland or Mali or Marshall Islands or Mauritania or Mauritius or Agalega Islands or Mexico or Micronesia or Middle East or Moldova or Moldovia or Moldovian or Mongolia or Montenegro or Morocco or Ifni or Mozambique or Myanmar or Myanma or Burma or Namibia or Nauru or Nepal or Nicaragua or Niger or Nigeria or Pakistan or Papua New Guinea or Palestine or Paraguay or Peru or Philippines or Philipines or Phillipines or Phillippines or Romania or Rumania or Roumania or Russia or Russian or Rwanda or Ruanda or Saint Lucia or St Lucia or Saint Vincent or St Vincent or Grenadines or Samoa or Samoan Islands or Navigator Island or Navigator Islands or Sao Tome or Senegal or Serbia or Montenegro or Sierra Leone or Sri Lanka or Ceylon or Solomon Islands or Somalia or South Africa or Sudan or South Sudan or Suriname or Surinam or Swaziland or Eswatini or Syria or Syrian Arab Republic or Tajikistan or Tadzhikistan or Tadjikistan or Tadzhik or Tanzania or Thailand or Togo or Togolese Republic or Tonga or Tunisia or Turkey or Turkmenistan or Turkmen or Tuvalu or Uganda or Ukraine or USSR or Soviet Union or Union of Soviet Socialist Republics or Uzbekistan or Uzbek or Vanuatu or New Hebrides or Venezuela or Vietnam or Viet Nam or West Bank or Yemen or Yugoslavia or Zambia or Zimbabwe or Rhodesia).hw,kw,ti,ab,cp.

164. ((developing or less* developed or under developed or underdeveloped or middle income or low* income or underserved or under served or deprived or poor*) adj2 (countr* or nation? or population? or world)).tw,kw.

165. ((developing or less* developed or under developed or underdeveloped or middle income or low* income) adj2 (economy or economies)).tw,kw.

166. (low* adj2 (gdp or gnp or gross domestic or gross national)).tw,kw.

167. (low adj3 middle adj3 (countr* or nation*)).tw,kw.

168. (lmic or lmics or third world or lami countr*).tw,kw.

169. transitional countr*.tw,kw.
low income country/ or middle income country/

**Measurement Concept**

170. Health status indicator/

171. (indicator* or "health indicator" or evaluation or tool* or assessment or methodolog* or standards or "unmet need" or coverage).tw,kw.

172. (monitor* or surveillance or screening).tw,kw.

173. ("rapid counting" or "aerial surveillance" or "flow monitoring" or "enumeration" or "reproductive health assessment toolkit").tw,kw.

Needs assessment/

174. ((rapid or needs) adj2 (assessment or evaluation)).tw,kw.

Exp questionnaire/ or data collection/ or “surveys and questionnaires”/ or “health survey”/ or contraceptive prevalence survey/ or health care survey/

175. ("data collection tool*" or "data source*" or "data collection").tw,kw.

176. (scale* or survey* or questionnaire*).tw,kw.

178. (index or indices).tw,kw.

179. ("minimum initial service package" or MISP).tw,kw.

180. (measure* or metric* or method*).tw,kw.

Health care delivery/ or health equity/ or health care quality/ or health care access/

181. "health access".tw,kw.

Attitude to health/

Health services research/

Health status/

Morbidity/ or maternal morbidity/ or perinatal morbidity/

Mortality/ or maternal mortality/ or mortality rate/ or perinatal mortality/ or prenatal mortality/ or standardized mortality ratio/

Prevalence/ or human immunodeficiency virus prevalence/

(morbidity or mortality or incidence or prevalence).tw,kw

disease burden/ or disability-adjusted life year/ or quality adjusted life year/

(outcome* adj3 (health or measur* or assess* or (score or scoring) or index or indices or scale* or monitor*)).tw,kw.

**<u>Database(s): Medline</u>**

**Search Strategy:**

**Search strategy for measurement of sexual and reproductive health rights in conflict and humanitarian settings in LMICs**

| | **Concept 1: Humanitarian crises and conflict settings** |
|---|---|
| 1 | disasters/ or emergencies/ or mass casualty incidents/ or natural disasters/ |
| 2 | disaster victims/ |
| 3 | ((disaster or emergenc*) adj2 victim*).tw,kf. |
| 4 | ((disaster or disasters or catastrophe or catastrophes) adj5 (environ* or human or manmade or "man made" or nature or natural or weather)).tw,kf. |
| 5 | ("mass casualty" or "mass casualties" or "mass fatalities" or "mass fatality").tw,kf. |
| 6 | ((crisis or crises) adj5 (environ* or human or manmade or "man made" or nature or natural or weather or setting*)).tw,kf. |
| 7 | ((crisis or crises or conflict) adj3 affected).tw,kf. |
| 8 | "warfare and armed conflicts"/ or armed conflict/ or warfare/ or war crimes/ or ethnic cleansing/ or genocide/ or holocaust/ or war exposure/ |
| 9 | ("warfare and armed conflict*" or warfare or "armed conflict" or "war crime*" or "ethnic cleansing*" or "gas poisoning" or genocide or holocaust or (war adj3 exposure)).tw,kf. |
| 10 | afghan campaign 2001-/ or gulf war/ or iraq war, 2003-2011/ |
| 11 | ("afghan campaign" or "gulf war" or "iraq war" or "war time" or "wartime" or "war torn" or "war affected" or "insurgency" or "intra conflict").tw,kf. |
| 12 | ((armed or zone or political or civil or setting*) adj3 (conflict or conflicts or attack or attacks or war or wars or "no fly")).tw,kf. |
| 13 | (Fragile adj2 (state* or countr* or nation* or situation* or setting*)).tw,kf. |
| 14 | ("Post conflict" or "postconflict" or "post war" or or "peacebuilding" or peacekeeping).tw,kf. |
| 15 | (war adj3 related).tw,kf. |

| | |
|---|---|
| 16 | ("militant group?" or "militant organization" or "militant organizations" or "militant organisation" or "militant organisations" or militia or combatant or rebel*).tw,kf. |
| 17 | Disaster Medicine/ |
| 18 | disease outbreaks/ |
| 19 | Epidemics/ |
| 20 | ("disaster medicine" or "disaster outbreak*" or “disease outbreak*“ or "epidemic*").tw,kf. |
| 21 | Emergency Medical Services/ |
| 22 | ((emergenc*) adj5 (environ* or human* or manmade or "man made" or nature or natural or weather or complex)).tw,kf. |
| 23 | Starvation/ |
| 24 | (famine or famines or starvation or starvations).tw,kf. |
| 25 | cyclonic storms/ or droughts/ or floods/ or tornadoes/ or tidal waves/ |
| 26 | avalanches/ or earthquakes/ or landslides/ or tidal waves/ or tsunamis/ or volcanic eruptions/ |
| 27 | (avalanche or avalanches or cyclone or cyclones or drought or droughts or earthquake or earthquakes or flood or flooded or flooding or floods or hurricane or hurricanes or landslide or landslides or "land slide" or "land slides" or mudslide or mudslides or "mud slide" or "mud slides" or storm or storms or tornado or tornadoes or tsunami or tsunamis or typhoon or typhoons or "volcanic ash" or "volcanic eruption" or "volcanic eruptions" or "volcanic gases" or rubble).tw,kf. |
| 28 | refugees/ |
| 29 | (evacuee or evacuees or refugee or refugees or squatter or squatters or transients or "asylum seeker?").tw,kf. |
| 30 | relief work/ or rescue work/ |
| 31 | ((rescue or relief or aid) adj3 (plan or plans or activity or activities or agency or agencies)).tw,kf. |
| 32 | ("aid plan" or "aid work" or "relief plan" or "relief work" or "rescue plan" or "rescue work").tw,kf. |

| | |
|---|---|
| 33 | ((staff or staffs or worker or workers) adj3 (relief or aid or rescue)).tw,kf. |
| 34 | (humanitarian assist*).tw,kf. |
| 35 | (humanitarian adj3 (aid or response or relief or crisis or crises or emergency or emergencies or disaster or disasters)).tw,kf. |
| 36 | Altruism/ |
| 37 | (humanitarianism or altruism).tw,kf. |
| 38 | ("displaced children" or "displaced child" or "displaced families" or "displaced family" or "displaced individuals" or "displaced internally" or "displaced men" or "displaced people" or "displaced peoples" or "displaced person" or "displaced persons" or "displaced population" or "displaced populations" or "displaced women" or "displaced adolescent" or "displaced adolescents" or "forced displacement" or "forced displacements" or "forcibl* displace*" or "internal displaced" or "internal displacement" or "internally displaced" or "population displaced" or "population displacement" or "forced migration" or "migrant*").tw,kf. |
| 39 | Exp refugee camps/ |
| 40 | (protected adj3 village*).tw,kf. |
| 41 | ((camp or camps) adj3 (refugee or transit or displace* or temporary or informal)).tw,kf. |
| 42 | (Settlement? adj3 (temporary or informal)).tw,kf. |
| 43 | 1 or 2 or 3 or 4 or 5 or 6 or 7 or 8 or 9 or 10 or 11 or 12 or 13 or 14 or 15 or 16 or 17 or 18 or 19 or 20 or 21 or 22 or 23 or 24 or 25 or 26 or 27 or 28 or 29 or 30 or 31 or 32 or 33 or 34 or 35 or 36 or 37 or 38 or 39 or 40 or 41 or 42 |
| | **Low- and middle-income countries** |
| 44 | Developing Countries.sh,kf. |
| 45 | (Africa or Asia or Caribbean or West Indies or South America or Latin America or Central America).hw,kf,ti,ab,cp. |
| 46 | (Afghanistan or Albania or Algeria or American Samoa or Angola or Armenia or Armenian or Azerbaijan or Bangladesh or Benin or Byelarus or Byelorussian or Belarus or Belorussian or Belorussia or Belize or Bhutan or Bolivia or Bosnia or Herzegovina or Hercegovina or Botswana or Brasil or Brazil or Bulgaria or Burkina Faso or Burkina Fasso or Upper Volta or Burundi or Urundi or Cambodia or Khmer Republic or Kampuchea or Cameroon or Cameroons or Cameron |

| | |
|---|---|
| | or Camerons or Cameroun or Cape Verde or Cabo Verde or Central African Republic or Chad or China or Colombia or Comoros or Comoro Islands or Comores or Mayotte or Congo or Zaire or Costa Rica or Cote d'Ivoire or Ivory Coast or Cuba or Djibouti or French Somaliland or Dominica or Dominican Republic or East Timor or East Timur or Timor Leste or Ecuador or Egypt or United Arab Republic or El Salvador or Equatorial Guinea or Eritrea or Ethiopia or Fiji or Gabon or Gabonese Republic or Gambia or Gaza or Georgia or Georgia Republic or Georgian Republic or Ghana or Gold Coast or Grenada or Guatemala or Guinea or Bissau or Guiana or Guyana or Haiti or Honduras or India or Maldives or Indonesia or Iran or Iraq or Jamaica or Jordan or Kazakhstan or Kazakh or Kenya or Kiribati or Korea or Democratic People's Republic of Korea or Kosovo or Kyrgyzstan or Kirghizia or Kyrgyz Republic or Kirghiz or Kirgizstan or Lao PDR or Laos or Lebanon or Lesotho or Basutoland or Liberia or Libya or Macedonia or Madagascar or Malagasy Republic or Malaysia or Malaya or Malay or Sabah or Sarawak or Malawi or Nyasaland or Mali or Marshall Islands or Mauritania or Mauritius or Agalega Islands or Mexico or Micronesia or Middle East or Moldova or Moldovia or Moldovian or Mongolia or Montenegro or Morocco or Ifni or Mozambique or Myanmar or Myanma or Burma or Namibia or Nauru or Nepal or Nicaragua or Niger or Nigeria or Pakistan or Papua New Guinea or Palestine or Paraguay or Peru or Philippines or Philipines or Phillipines or Phillippines or Romania or Rumania or Roumania or Russia or Russian or Rwanda or Ruanda or Saint Lucia or St Lucia or Saint Vincent or St Vincent or Grenadines or Samoa or Samoan Islands or Navigator Island or Navigator Islands or Sao Tome or Senegal or Serbia or Montenegro or Sierra Leone or Sri Lanka or Ceylon or Solomon Islands or Somalia or South Africa or Sudan or South Sudan or Suriname or Surinam or Swaziland or Eswatini or Syria or Syrian Arab Republic or Tajikistan or Tadzhikistan or Tadjikistan or Tadzhik or Tanzania or Thailand or Togo or Togolese Republic or Tonga or Tunisia or Turkey or Turkmenistan or Turkmen or Tuvalu or Uganda or Ukraine or USSR or Soviet Union or Union of Soviet Socialist Republics or Uzbekistan or Uzbek or Vanuatu or New Hebrides or Venezuela or Vietnam or Viet Nam or West Bank or Yemen or Yugoslavia or Zambia or Zimbabwe or Rhodesia).hw,kf,ti,ab,cp. |
| 47 | ((developing or less* developed or under developed or underdeveloped or middle income or low* income or underserved or under served or deprived or poor*) adj2 (countr* or nation? or population? or world)).tw,kf. |
| 48 | ((developing or less* developed or under developed or underdeveloped or middle income or low* income) adj2 (economy or economies)).tw,kf. |
| 49 | (low* adj2 (gdp or gnp or gross domestic or gross national)).tw,kf. |

| | |
|---|---|
| 50 | (low adj3 middle adj3 (countr* or nation*)).tw,kf. |
| 51 | transitional countr*.tw,kf. |
| 52 | 44 or 45 or 46 or 47 or 48 or 49 or 50 or 51 |
| 53 | 43 or 52 |
| 54 | 43 and 52 |
| | **Concept 2: Population of interest** |
| 55 | ((displace* or refugee?) adj3 (child* or famil* or men or wom* or individual* or adolescent* or people* or population* or person? or girl* or boy* or youth)).tw,kf. |
| 56 | adolescent/ or young adult/ |
| 57 | (adolescen* or teen* or youth or youths or "young adult" or "young adults" or girl* or boy* or "young wom*" or "young girl*" or "young boy*").tw,kf. |
| 58 | exp Pregnant Women/ or exp Women/ |
| | Mothers/ |
| 59 | (mother?).tw,kf. |
| 60 | female/ |
| 61 | parturients.tw,kf. |
| 62 | "sexually active".tw,kf. |
| 63 | refugees/ |
| 64 | (refugee or refugees).tw,kf. |
| 65 | (women adj2 reproductive adj2 age).tw,kf. |
| 66 | 55 or 56 or 57 or 58 or 59 or 60 or 61 or 62 or 63 or 64 or 65 |
| | **Concept 3: Measurement tools and indicators** |
| 67 | Health status indicators/ |
| 68 | (indicator* or "health indicator" or evaluation or tool* or assessment or methodolog* or standards or "unmet need" or coverage).tw,kf. |

| | |
|---|---|
| 69 | (monitor* or surveillance or screening).tw,kf. |
| 70 | ("rapid counting" or "aerial surveillance" or "flow monitoring" or "enumeration" or (reproductive health adj3 assessment toolkit)).tw,kf. |
| 71 | ((rapid or needs) adj (assessment or evaluation)).tw,kf. |
| 72 | ("data collection tool*" or "data source*" or "data collection").tw,kf. |
| 73 | data collection/ or "surveys and questionnaires"/ or contraceptive prevalence surveys/ or health care surveys/ or health surveys/ |
| 74 | (scale* or survey* or questionnaire*).tw,kf. |
| 75 | (index or indices).tw,kf. |
| 76 | ("minimum initial service package" or MISP).tw,kf. |
| 77 | (measure* or metric* or method*).tw,kf. |
| 78 | "delivery of health care"/ or health services accessibility/ or health equity/ |
| 79 | "health access".tw,kf. |
| 80 | Health Knowledge, Attitudes, Practice/ |
| 81 | health services research/ or needs assessment/ |
| | Health status/ |
| | morbidity/ or incidence/ or prevalence/ |
| | Mortality/ or maternal mortality/ or perinatal mortality/ |
| | (morbidity or mortality or incidence or prevalence).tw,kf. |
| | quality-adjusted life years/ |
| | (outcome* adj3 (health or measu* or assess* or (score or scoring) or index or indices or scale* or monitor*)).tw,kf. |
| 82 | 67 or 68 or 69 or 70 or 71 or 72 or 73 or 74 or 75 or 76 or 77 or 78 or 79 or 80 or 81 |
| | **Concept 4: SRHR** |
| 83 | Gynecology/ |

| | |
|---|---|
| 84 | (gynaecology or gynecology).tw,kf. |
| 85 | Reproductive Health/ |
| 86 | ("reproductive health" or "sexual health" or "reproductive services").tw,kf. |
| 87 | Sex Education/ |
| 88 | ("family planning education" or "family planning counselling" or "family planning instructor?" or "family planning training" or "sex* education" or "sex* instruction" or (reproductive health adj3 knowledge) or (HIV adj3 knowledge) or ("STI? adj3 knowledge) or (sex* adj5 knowledge)).tw,kf. |
| 89 | ("minimum initial service package" or MISP).tw,kf. |
| 90 | Maternal Health/ |
| 91 | Maternal health services/ |
| 92 | Maternal Welfare/ |
| 93 | ("maternal health" or "maternal welfare" or "maternal child health" or "maternal child welfare" or "maternal health services").tw,kf. |
| 94 | reproductive health services/ or family planning services/ or maternal health services/ |
| 95 | Obstetrics/ |
| 96 | exp Delivery, Obstetric/ |
| 97 | ((labor or labour) adj5 (birth* or breech or childbirth or childbirths or complicat* or difficult or early or easy or induce* or induction or late or obstetric* or onset or pregnan* or present*)).tw,kf. |
| 98 | Pregnancy/ |
| 99 | (pregnanc* or "child bearing" or childbearing).tw,kf. |
| 100 | Prenatal Care/ |
| 101 | ("prenatal" or "pre natal" or "antenatal" or "ante natal").tw,kf. |
| 102 | Perinatal Care/ |
| 103 | ("perinatal care" or "peri natal care" or "perinatal care" or "peri natal care").tw,kf. |

| | |
|---|---|
| 104 | Peripartum Period/ |
| 105 | ("peripartum* period*" or "perinatal* period*" or "peri natal* period*").tw,kf. |
| 106 | Parturition/ |
| 107 | (birth or childbirth* or parturition* or "safe delivery" or "safely delivered").tw,kf. |
| 108 | (antepartum or "ante partum" or intrapartum or "intra partum").tw,kf. |
| 109 | Stillbirth/ |
| 110 | (stillbirth or stillbirths or stillborn or stillborns or "still birth" or "still births" or "still born" or "still borns").tw,kf. |
| 111 | ("emergency obstetric care" or "basic emergency obstetric care" or "comprehensive emergency obstetric care" or "basic emoc" or "comprehensive emoc").tw,kf. |
| 112 | (emoc or emonc or cemoc or bemoc).tw,kf. |
| 113 | Midwifery/ |
| 114 | ("birth attendant" or "birth attendants" or midwife or midwives or midwifery or "traditional birth attendant*" or "skilled birth attendant*").tw,kf. |
| 115 | Postnatal Care/ |
| 116 | Postpartum Period/ |
| 117 | (postnatal or "post natal" or postpartum or "post partum" or puerperium or puerperal).tw,kf. |
| 118 | Abortion, Spontaneous/ |
| 119 | miscarriage*.tw,kf. |
| 120 | abortion, induced/ or abortion, eugenic/ or abortion, legal/ or abortion, therapeutic/ |
| 121 | Abortion, Septic/ |
| 122 | (abortion or abortions or aborted or aborting or abortus).tw,kf. |
| 123 | ((unwanted or unintended) adj3 pregnanc*).tw,kf. |
| 124 | (Abortion adj3 (safe or unsafe or post or care)).tw,kf. |
| 125 | Misoprostol/ |

| | |
|---|---|
| 126 | Mifepristone/ |
| 127 | (Misoprostol or mifepristone or cytotec or mifeprex or mifegymiso or "abortion pill").tw,kf. |
| 128 | Birth Intervals/ |
| 129 | ("birth interval" or "birth intervals" or "birth spacing" or "birth spacings" or "child spacing" or "child spacings" or "family building" or "family planning" or "pregnancy interval" or "pregnancy intervals").tw,kf. |
| 130 | "safe motherhood".tw,kf. |
| 131 | contraception/ or coitus interruptus/ or contraception, barrier/ or contraception, postcoital/ or natural family planning methods/ or contraceptive effectiveness/ or long-acting reversible contraception/ or sterilization, reproductive/ |
| 132 | ("birth regulation" or "birth control*" or "coitus interruptus" or "fertility control" or contracept* or "conception control" or antifertility or anticonception or "fertility control" or "fertili?ation inhibition" or "inhibition of fertili?ation" or "fertili?ation inhibition" or "inhibition of fertili?ation" or "pregnancy prevent*").tw,kf. |
| 133 | exp Contraceptive Devices/ |
| 134 | ("cervical cap" or "cervical caps" or "i.u.d." or "intra uterine device*" or "intracervical device" or "intrauterine device*" or "iucd" or "iud" or "iuds" or "progestin implant" or "vaginal diaphragm" or "vaginal diaphragms" or "vaginal ring" or "vaginal shield" or "vaginal rings" or "vaginal shields" or "vaginal sponge" or "vaginal sponges" or "coiled spring*" or condom or condoms or spermicide or "rhythm method" or "calendar method" or "pull out method" or "tubal ligation" or vasectomy or "depo provera").tw,kf. |
| 135 | (("birth control" or contracept*) adj3 (pill or patch or implant or shot or injection or withdrawal)).tw,kf. |
| 136 | Contraceptives, Postcoital/ |
| 137 | ("morning after pill" or "postcoital antifertility agent" or "postcoital pill" or "plan B" or "emergency contraception").tw,kf. |
| 138 | Sexual Abstinence/ |
| 139 | (celibacy or "postpartum abstinence" or "sexual abstinence" or virginity).tw,kf. |

| | |
|---|---|
| 140 | hiv/ or hiv-1/ or hiv-2/ |
| 141 | Human immunodeficiency virus/ |
| 142 | ("acquired immune deficiency syndrome virus" or "acquired immunodeficiency syndrome virus" or "aids associated lentivirus" or "aids associated retrovirus" or "aids associated virus" or "aids related virus" or "aids virus" or "aids viruses" or "hiv" or "human immuno deficiency virus" or "human immunodeficiency virus" or "human immunodeficiency viruses" or "aids related illness*").tw,kf. |
| 143 | HIV Infections/ |
| 144 | ("hiv infection*" or "hiv seropositivit*" or "hiv coinfection*").tw,kf. |
| 145 | Acquired Immunodeficiency Syndrome/ |
| 146 | ("aids" or "acquired immune deficiency syndrome*" or "acquired immuno deficiency syndrome*" or "acquired immunodeficiency syndrome*" or "aids related opportunistic infections").tw,kf. |
| 147 | Infectious Disease Transmission, Vertical/ |
| 148 | Post-exposure prophylaxis/ |
| 149 | ("post exposure prophylaxis" or PEP).tw,kf. |
| 150 | Antiretroviral Therapy, Highly Active/ |
| 151 | Anti-HIV agents/ |
| 152 | ("Antiretroviral therapy" or "antiretroviral treatment" or ART or ARV or antiretroviral* or HAART or "anti hiv agent*").tw,kf. |
| 153 | (pmtct or "prevention of HIV mother to child transmission" or "prevention of mother to child transmission" or "eliminate mother to child transmission" or "prevention of mother to child HIV transmission").tw,kf. |
| 154 | Sexually Transmitted Diseases/ |
| 155 | ("sexually transmitted disease*" or "sexually transmitted infection*" or "venereal disease*" or "venereal infection" or "venereal infections" or STIs or STDs).tw,kf. |

| | |
|---|---|
| 156 | sexually transmitted diseases, bacterial/ or chancroid/ or chlamydia infections/ or chlamydia trachomatis/ or gonorrhea/ or syphilis/ or Trichomonas Vaginitis/ |
| 157 | sexually transmitted diseases, viral/ or herpes genitalis/ or Condylomata Acuminata/ or Hepatitis B/ |
| 158 | ("chancroid" or "chancroids" or chlamidiosis or "chlamydia infection*" or "chlamydia trachomatis" or "gonorrhea" or "granuloma inguinale" or "granuloma venereum" or "Klebsiella granulomatis infection" or "Neisseria gonorrhoeae infection" or "syphilis" or "syphilitic disorder" or trichomoniasis or "trichomonas vaginitis").tw,kf. |
| 159 | Papillomavirus Infections/ |
| 160 | ("anal wart" or "anal warts" or "anogenital wart" or "anogenital warts" or "condyla acuminatum" or "condylatum acuminatum" or "condyloma accuminatum" or "condyloma acuminata" or "condylomata acuminata" or "genital herpes" or "genital wart" or "genital warts" or "herpes genitalis" or "herpes progenitalis" or "herpes simplex genitalis" or "herpes simplex virus genital infection" or "penile wart" or "penile warts" or "perianal wart" or "perianal warts" or "venereal wart" or "venereal warts" or "human papillomavirus*" or HPV or "papillomavirus infection*" or "hepatitis B").tw,kf. |
| 161 | Scabies/ |
| 162 | ("sarcoptic mange" or scabies).tw,kf. |
| 163 | Phthirus/ |
| 164 | ("crab lice" or "crab lices" or "crab louse" or "crab louses" or "Pediculus pubis" or "phthirus" or "Phtirus pubis" or "Pthirus pubis" or "pubic lice" or "pubic louse").tw,kf. |
| 165 | intimate partner violence/ |
| 166 | Intimate Partner Abuse/ or Spouse abuse/ or Domestic Violence/ |
| 167 | ("partner abuse" or "partner violence" or "wife abuse" or "spouse abuse" or "spousal abuse" or "domestic violence" or "domestic abuse" or "gender based violence" or "gender-based violence" or "GBV" or "IPV" or "sex* based violence").tw,kf. |
| 168 | ((abuse* or assault* or violence) adj2 (woman or women)).tw,kf. |
| 169 | Sex offenses/ or Human Trafficking/ or Rape/ or Circumcision, Female/ |

| | |
|---|---|
| 170 | ("coerced intercourse" or "forced intercourse" or "forced prostitution" or "forced sex" or "human trafficking" or "human traffickings" or rape or "sex trafficking" or "sex traffickings" or "sex* abuse*" or "sex* assault*" or "sex* crime*" or "sex* offense" or "sex* offenses" or "sex* slave*" or "sexual aggression" or "sexual bullying" or "sexual coercion" or "sexual exploitation*" or "sexual harassment" or "sexual trauma" or "sexual violence" or "unwanted sex" or "unlawful sex" or "honor killings" or "transactional sex").tw,kf. |
| 171 | ("physical* abuse*" or "physical* assault*" or "physical violence" or "psychological violence" or "emotional violence" or "economic violence" or "female circumcision" or "female genital mutilation" or "female genital cutting" or FGM).tw,kf. |
| 172 | fistula/ or vaginal fistula/ or rectovaginal fistula/ |
| 173 | (fistula or fistulas or "genital trauma" or "genital injury" or "vaginal trauma" or "vaginal injury").tw,kf. |
| 175 | pregnancy complications/ or abortion, spontaneous/ or diabetes, gestational/ or hypertension, pregnancy-induced/ or obstetric labor complications/ |
| 176 | Eclampsia/ or Pre-eclampsia/ |
| 177 | ("toxaemia" or "toxemia" or "hypertension edema proteinuria gestosis" or "pre eclampsia" or "pre eclamptic toxaemia" or "pre eclamptic toxemia" or "preeclamptic toxaemia" or "preeclamptic toxemia" or "pregnancy toxaemia*" or "pregnancy toxemia*" or "toxemia of pregnanc*" or "toxemic pregnancy" or eclampsia or preeclampsia).tw,kf. |
| 178 | dystocia/ |
| 179 | exp Obstetric Labor Complications/ |
| 180 | ("abnormal labor" or "abnormal labour" or "delayed labor" or "delayed labour" or "inertia uteri" or "labor obstruction" or "labour obstruction" or "obstructed labor" or "obstructed labour" or "uterus inertia" or dystocia or dystocias).tw,kf. |
| 181 | Breech Presentation/ |
| 182 | (breech adj2 (present* or position*)).tw,kf. |
| 183 | Uterine Hemorrhage/ |
| 184 | Postpartum hemorrhage/ |

| | |
|---|---|
| 185 | ("vagina* haemorrhage" or "vagina* hemorrhage" or "vaginal bleeding").tw,kf. |
| 186 | Menstruation/ |
| 187 | ((Menstruation or menarche or menstrual or menses) adj2 (hygiene or bleeding or cycle)).tw,kf. |
| 188 | "Reproductive rights"/ or "women’s rights"/ |
| 189 | ((sexual or reproductive or "women’s") adj3 rights).tw,kf. |
| 190 | ("Sexual initiation" or "early sexual debut").tw,kf. |
| 191 | Or /83-190 |

**Database(s): PAIS Index – 8th May 2019**

**Humanitarian Concept:**

AB,IF,SU,TI,MESH("disaster*" OR "emergency" OR "emergencies" OR "mass casualty incident*" OR "conflict* " OR "refugee* " OR "migrant* " OR " forced displace* " OR "natural disaster*" OR "war related" OR "disaster victim" OR "starvation" OR "outbreak*" OR "epidemic*" OR "famine" OR "drought" OR "flood*" OR "complex emergenc*" OR "post conflict" OR "post war" OR "armed conflict" OR "conflict related" OR "relief work" OR "rescue work" OR "displace*" OR "asylum seeker" OR "forced migration" OR "internal displace*" OR "warfare" OR "war" OR "cyclon*" OR "hurricane" OR "earthquake" OR "typhoon" OR "crisis" OR "humanitarian" OR "humanitarian aid")

**LMIC Concept:**

AB,IF,SU,TI,MESH("developing countries" OR "Africa" OR "Asia" OR "Caribbean" OR "West Indies" OR "South America" OR "Latin America" OR "Central America" OR "low-income countr*" OR "middle-income countr*" OR (Low N/2 middle income) OR “Afghanistan” or “Albania” or “Algeria” or “American Samoa” or “Angola” or “Armenia” or “Armenian” or Azerbaijan or Bangladesh or Benin or Byelarus or Byelorussian or Belarus or Belorussian or Belorussia or Belize or Bhutan or Bolivia or Bosnia or Herzegovina or Hercegovina or Botswana or Brasil or Brazil or Bulgaria or “Burkina Faso” or “Burkina Fasso” or “Upper Volta” or Burundi or Urundi or Cambodia or “Khmer Republic” or Kampuchea or Cameroon or Cameroons or Cameron or Camerons or Cameroun or “Cape Verde” or “Cabo Verde” or “Central African Republic” or Chad or China or Colombia or Comoros or Comoro Islands or Comores or Mayotte or Congo or Zaire or “Costa Rica” or “Cote d'Ivoire” or “Ivory Coast” or Cuba or Djibouti or “French Somaliland” or Dominica or “Dominican Republic” or “East Timor” or “East Timur” or “Timor Leste” or Ecuador or Egypt or “United Arab Republic” or “El Salvador” or “Equatorial Guinea” or Eritrea or Ethiopia or Fiji or Gabon or “Gabonese Republic” or Gambia or Gaza or Georgia or “Georgia Republic” or “Georgian Republic”or Ghana or “Gold Coast” or Grenada or Guatemala or Guinea or Bissau or Guiana or Guyana or Haiti or Honduras or India or Maldives or Indonesia or Iran or Iraq or Jamaica or Jordan or Kazakhstan or Kazakh or Kenya or Kiribati or Korea or Kosovo or Kyrgyzstan or Kirghizia or “Kyrgyz Republic” or Kirghiz or Kirgizstan or “Lao PDR” or Laos or Lebanon or Lesotho or Basutoland or Liberia or Libya or Macedonia or Madagascar or “Malagasy Republic” or Malaysia or Malaya or Malay or Sabah or Sarawak or Malawi or Nyasaland or Mali or “Marshall Islands” or Mauritania or Mauritius or “Agalega Islands” or Mexico or Micronesia or “Middle East” or Moldova or Moldovia or Moldovian or Mongolia or Montenegro or Morocco or Ifni or Mozambique or Myanmar or Myanma or Burma or Namibia or Nauru or Nepal or Nicaragua or Niger or Nigeria or Pakistan or “Papua New Guinea” or Palestine or Paraguay or Peru or Philippines or Philipines or Phillipines or Phillippines or Romania or Rumania or Roumania or Russia or Russian or Rwanda or Ruanda or “Saint Lucia” or “St Lucia” or “Saint Vincent” or “St Vincent” or Grenadines or Samoa or “Samoan Islands” or “Navigator Island” or “Navigator Islands” or “Sao Tome” or Senegal or Serbia or Montenegro or “Sierra Leone” or “Sri Lanka” or Ceylon or “Solomon Islands” or Somalia or “South Africa” or Sudan or “South Sudan” or Suriname or Surinam or Swaziland or Eswatini or Syria or “Syrian Arab Republic” or Tajikistan or Tadzhikistan or Tadjikistan or Tadzhik or Tanzania or Thailand or Togo or “Togolese Republic” or Tonga or Tunisia or Turkey or Turkmenistan or Turkmen or Tuvalu or Uganda or Ukraine or USSR or “Soviet Union” or Uzbekistan or Uzbek or Vanuatu or “New Hebrides” or Venezuela or Vietnam or “Viet Nam” or “West Bank” or Yemen or Yugoslavia or Zambia or Zimbabwe or Rhodesia or "transitional countr*")

**Measurement Concept:**

AB,IF,SU,TI,MESH(“Health status indicators” OR “indicators" OR "health indicator" OR "morbidity" OR "mortality" OR "prevalence" OR "maternal morbidity" OR "maternal mortality" OR "perinatal mortality" OR “evaluation” OR "measure*" or “tool” or “assessment” or “methodolog*” or "method*" or “standard*” or "unmet need" or “coverage” OR “data collection” or "surveys and questionnaires" or "health care surveys" OR “contraceptive prevalence surveys” or ”health surveys” OR “scale*” or “survey*” or “questionnaire*” OR "psychometric*" OR "monitoring" OR "intervention" OR "needs assessment" OR "index" OR "indices" OR "metric*" OR "health access" OR "attitude*" OR "research" OR "outcome assessment" OR "outcome measure*" OR "disease burden" OR "quality adjusted life year" OR "disability adjusted life year")

**SRHR Concept:**

AB,IF,TI,SU,MESH("reproductive health" OR "sexual health" OR "reproductive services" OR "prenatal care " OR "abortion" OR "gender based violence " OR "sexual violence" OR "menstruation" OR "human immunodeficiency virus" OR "HIV" OR "AIDS" OR "Acquired immune deficiency syndrome" OR "sexually transmitted diseases" OR "STIs" OR "STDs" OR "sexual harassment" OR "gynecology" OR "family planning" OR "reproductive health knowledge" OR "sex* education" OR "pregnancy" OR "birth control" OR " contracepti*" OR "condom" OR "intimate partner violence" OR "reproductive rights" OR "birth attendant" OR "midwi*" OR "maternal health" OR "childbirth" OR "obstetrics" OR "labor complications" OR "eclampsia" OR "fistula" OR "stillbirth" OR "womens health" OR "postpartum period" OR "perinatal care" OR "postnatal care" OR "antenatal care")

AB,IF,TI,SU,MESH(adolescent OR girl OR wom*n OR "young adult")

**Database(s): PsycINFO**

**Search Strategy:**

| # | Searches | Results |
|---|---|---|
| 1 | natural disasters/ or disasters/ or emergency management/ or emergency preparedness/ or emergency services/ | 16680 |
| 2 | ((disaster or emergenc*) adj2 victim*).tw,id. | 415 |
| 3 | ((disaster? or catastrophe?) adj5 (environ* or human or manmade or "man made" or nature or natural or weather)).tw,id. | 4133 |
| 4 | ("mass casualty" or "mass casualties" or "mass fatalities" or "mass fatality").tw,id. | 173 |
| 5 | ((crisis or crises) adj5 (environ* or human or manmade or "man made" or nature or natural or weather or setting*)).tw,id. | 1385 |
| 6 | ((crisis or crises or conflict) adj3 affected).tw,id. | 576 |
| 7 | war/ or conflict/ or violence/ | 60046 |
| 8 | genocide/ or holocaust/ or mass murder/ | 2317 |
| 9 | genocide/ or mass murder/ | 1145 |
| 10 | ("warfare and armed conflict*" or warfare or "armed conflict*" or "war crime*" or "ethnic cleansing*" or "gas poisoning" or genocide or holocaust or "war exposure").tw,id. | 7685 |
| 11 | ("afghan campaign" or "gulf war" or "iraq war" or "war time" or "wartime" or "war torn" or "war affected" or "insurgency" or "intra conflict").tw,id. | 3940 |
| 12 | ((armed or zone or political or civil or setting*) adj3 (conflict or conflicts or attack or attacks or war or wars or "no fly")).tw,id. | 5297 |
| 13 | (Fragile adj2 (state* or countr* or nation* or situation* or setting*)).tw,id. | 137 |
| 14 | ("Post conflict" or "postconflict" or "post war" or "post conflict setting" or "peacebuilding" or peacekeeping).tw,id. | 3198 |
| 15 | "war related".tw,id. | 789 |

| | | |
|---|---|---|
| 16 | ("militant group?" or "militant organization" or "militant organizations" or "militant organisation" or "militant organisations" or militia or combatant or rebel*).tw,id. | 3292 |
| 17 | epidemics/ or pandemics/ | 3190 |
| 18 | ("disaster medicine" or "disaster outbreak*" or "disease outbreak?" or "epidemic*").tw,id. | 12183 |
| 19 | ((emergency or emergencies) adj5 (environ* or human or manmade or "man made" or nature or natural or weather or complex)).tw,id. | 585 |
| 20 | starvation/ | 377 |
| 21 | (famine or famines or starvation or starvations).tw,id. | 2154 |
| 22 | (avalanche or avalanches or cyclone or cyclones or drought or droughts or earthquake or earthquakes or flood or flooded or flooding or floods or hurricane or hurricanes or landslide or landslides or "land slide" or "land slides" or mudslide or mudslides or "mud slide" or "mud slides" or storm or storms or tornado or tornadoes or tsunami or tsunamis or typhoon or typhoons or "volcanic ash" or "volcanic eruption" or "volcanic eruptions" or "volcanic gases" or rubble).tw,id. | 10346 |
| 23 | refugees/ or asylum seeking/ | 5577 |
| 24 | (evacuee or evacuees or refugee or refugees or squatter or squatters or transients or "asylum seeker").tw,id. | 9857 |
| 25 | rescue workers/ | 239 |
| 26 | ((rescue or relief or aid) adj3 (plan or plans or activity or activities or agency or agencies)).tw,id. | 648 |
| 27 | ("aid plan" or "aid work" or "relief plan" or "relief work" or "rescue plan" or "rescue work").tw,id. | 169 |
| 28 | ((staff or staffs or worker or workers) adj3 (relief or aid)).tw,id. | 475 |
| 29 | prosocial behavior/ or altruism/ | 10594 |
| 30 | (humanitarian assistance or humanitarian assistances or relief work or relief works).tw,id. | 203 |

| | | |
|---|---|---|
| 31 | ("aid plan*" or "aid work*" or "relief plan*" or "relief work*" or "rescue plan*" or "rescue work*").tw,id. | 608 |
| 32 | (humanitarian adj3 (aid or response or relief or crisis or crises or emergency or emergencies or disaster or disasters)).tw,id. | 628 |
| 33 | (humanitarianism or altruism).tw,id. | 6095 |
| 34 | ("displaced children" or "displaced child" or "displaced families" or "displaced family" or "displaced individuals" or "displaced internally" or "displaced men" or "displaced people" or "displaced peoples" or "displaced person" or "displaced persons" or "displaced population" or "displaced populations" or "displaced women" or "displaced adolescent" or "displaced adolescents" or "forced displacement" or "forced displacements" or "forcibl* displace*" or "internal displaced" or "internal displacement" or "internally displaced" or "population displaced" or "population displacement" or "forced migration" or "migrant*").tw,id. | 10404 |
| 35 | (camp? adj3 (refugee or transit or displace* or temporary or informal)).tw,id. | 620 |
| 36 | ((camp or camps) adj3 (refugee or transit or displace* or temporary or informal)).tw,id. | 620 |
| 37 | protected village?.tw,id. | 1 |
| 38 | (Settlement? adj3 (temporary or informal)).tw,id. | 167 |
| 39 | crises/ | 5267 |
| 40 | 1 or 2 or 3 or 4 or 5 or 6 or 7 or 8 or 9 or 10 or 11 or 12 or 13 or 14 or 15 or 16 or 17 or 18 or 19 or 20 or 21 or 22 or 23 or 24 or 25 or 26 or 27 or 28 or 29 or 30 or 31 or 32 or 33 or 34 or 35 or 36 or 37 or 38 or 39 | 146302 |
| 41 | developing countries/ | 5228 |
| 42 | Developing Countries.sh,id. | 5522 |
| 43 | (Africa or Asia or Caribbean or West Indies or South America or Latin America or Central America).hw,id,ti,ab,cp. | 37760 |
| 44 | (Afghanistan or Albania or Algeria or American Samoa or Angola or Armenia or Armenian or Azerbaijan or Bangladesh or Benin or Byelarus or Byelorussian or | 198674 |

| | |
|---|---|
| | Belarus or Belorussian or Belorussia or Belize or Bhutan or Bolivia or Bosnia or Herzegovina or Hercegovina or Botswana or Brasil or Brazil or Bulgaria or Burkina Faso or Burkina Fasso or Upper Volta or Burundi or Urundi or Cambodia or Khmer Republic or Kampuchea or Cameroon or Cameroons or Cameron or Camerons or Cameroun or Cape Verde or Cabo Verde or Central African Republic or Chad or China or Colombia or Comoros or Comoro Islands or Comores or Mayotte or Congo or Zaire or Costa Rica or Cote d'Ivoire or Ivory Coast or Cuba or Djibouti or French Somaliland or Dominica or Dominican Republic or East Timor or East Timur or Timor Leste or Ecuador or Egypt or United Arab Republic or El Salvador or Equatorial Guinea or Eritrea or Ethiopia or Fiji or Gabon or Gabonese Republic or Gambia or Gaza or Georgia or Georgia Republic or Georgian Republic or Ghana or Gold Coast or Grenada or Guatemala or Guinea or Bissau or Guiana or Guyana or Haiti or Honduras or India or Maldives or Indonesia or Iran or Iraq or Jamaica or Jordan or Kazakhstan or Kazakh or Kenya or Kiribati or Korea or Democratic People's Republic of Korea or Kosovo or Kyrgyzstan or Kirghizia or Kyrgyz Republic or Kirghiz or Kirgizstan or Lao PDR or Laos or Lebanon or Lesotho or Basutoland or Liberia or Libya or Macedonia or Madagascar or Malagasy Republic or Malaysia or Malaya or Malay or Sabah or Sarawak or Malawi or Nyasaland or Mali or Marshall Islands or Mauritania or Mauritius or Agalega Islands or Mexico or Micronesia or Middle East or Moldova or Moldovia or Moldovian or Mongolia or Montenegro or Morocco or Ifni or Mozambique or Myanmar or Myanma or Burma or Namibia or Nauru or Nepal or Nicaragua or Niger or Nigeria or Pakistan or Papua New Guinea or Palestine or Paraguay or Peru or Philippines or Philipines or Phillipines or Phillippines or Romania or Rumania or Roumania or Russia or Russian or Rwanda or Ruanda or Saint Lucia or St Lucia or Saint Vincent or St Vincent or Grenadines or Samoa or Samoan Islands or Navigator Island or Navigator Islands or Sao Tome or Senegal or Serbia or Montenegro or Sierra Leone or Sri Lanka or Ceylon or Solomon Islands or Somalia or South Africa or Sudan or South Sudan or Suriname or Surinam or Swaziland or Eswatini or Syria or Syrian Arab Republic or Tajikistan or Tadzhikistan or Tadjikistan or Tadzhik or Tanzania or Thailand or Togo or Togolese Republic or Tonga or Tunisia or Turkey or Turkmenistan or Turkmen or Tuvalu or Uganda or Ukraine or USSR or Soviet Union or Union of Soviet Socialist Republics or Uzbekistan or Uzbek or Vanuatu or New Hebrides or Venezuela or Vietnam or Viet Nam or West Bank or Yemen or Yugoslavia or Zambia or Zimbabwe or Rhodesia).hw,id,ti,ab,cp. |

| | | |
|---|---|---|
| 45 | ((developing or less* developed or under developed or underdeveloped or middle income or low* income or underserved or under served or deprived or poor*) adj2 (countr* or nation? or population? or world)).tw,id. | 18646 |
| 46 | ((developing or less* developed or under developed or underdeveloped or middle income or low* income) adj2 (economy or economies)).tw,id. | 438 |
| 47 | (low* adj2 (gdp or gnp or gross domestic or gross national)).tw,id. | 51 |
| 48 | (lmic or lmics or third world or lami countr*).tw,id. | 1716 |
| 49 | transitional countr*.tw,id. | 64 |
| 50 | 41 or 42 or 43 or 44 or 45 or 46 or 47 or 48 or 49 | 225672 |
| 51 | ((displace* or refugee?) adj3 (child* or famil* or men or wom* or individual* or adolescent* or people* or population* or person?* or girl* or boy* or youth)).tw,id. | 3750 |
| 52 | exp mothers/ | 39047 |
| 53 | exp human females/ | 139939 |
| 54 | "mother?".tw,id. | 119113 |
| 55 | ("pregnant wom*n" or wom*n).tw,id. | 284068 |
| 56 | parturients.tw,id. | 60 |
| 57 | "sexually active".tw,id. | 3742 |
| 58 | refugees/ or asylum seeking/ | 5577 |
| 59 | (refugee or refugees).tw,id. | 7869 |
| 60 | (women adj2 reproductive adj2 age).tw,id. | 712 |
| 61 | 51 or 52 or 53 or 54 or 55 or 56 or 57 or 58 or 59 or 60 | 416365 |
| 62 | ("health status indicator*" or indicator* or "health indicator" or evaluation or tool* or assessment or methodolog* or standards or "unmet need" or coverage).tw,id. | 809275 |
| 63 | (monitor* or surveillance or screening).tw,id. | 154228 |

| | | |
|---|---|---|
| 64 | ("rapid counting" or "aerial surveillance" or "flow monitoring" or "enumeration" or (reproductive health adj3 assessment toolkit)).tw,id. | 1137 |
| 65 | evaluation/ or needs assessment/ or program evaluation/ or measurement/ | 83222 |
| 66 | ((rapid or needs) adj2 (assessment or evaluation)).tw,id. | 5746 |
| 67 | data collection/ or methodology/ or surveys/ | 47169 |
| 68 | ("data collection tool*" or "data source*" or "data collection").tw,id. | 37577 |
| 69 | (scale* or survey* or questionnaire*).tw,id. | 763587 |
| 70 | questionnaires/ | 17761 |
| 71 | (index or indices).tw,id. | 169137 |
| 72 | "index (testing)"/ | 150 |
| 73 | ("minimum initial service package" or MISP).tw,id. | 3 |
| 74 | (measure* or metric* or method*).tw,id. | 1381127 |
| 75 | psychometrics/ | 58014 |
| 76 | "health access".tw,id. | 242 |
| 77 | (index or indices).tw,id. | 169137 |
| 78 | morbidity/ or comorbidity/ | 35227 |
| 79 | "death and dying"/ or mortality rate/ | 34358 |
| 80 | ("disease burden" or "quality adjusted life year" or "disability adjusted life year").tw,id. | 2053 |
| 81 | (outcome* adj3 (health or measur* or assess* or (score or scoring) or index or indices or scale* or monitor*)).tw,id. | 84257 |
| 82 | (morbidity or mortality or incidence or prevalence).tw,id. | 192686 |
| 83 | or/62-82 | 2266428 |

| | | |
|---|---|---|
| 84 | gynecology/ | 775 |
| 85 | (gynaecology or gynecology).tw,id. | 1561 |
| 86 | reproductive health/ | 2927 |
| 87 | ("reproductive health" or "sexual health" or "reproductive services").tw,id. | 8297 |
| 88 | sex education/ | 3480 |
| 89 | ("family planning education" or "family planning counselling" or "family planning instructor*" or "family planning training" or "sex* education" or "sex* instruction" or (reproductive health adj3 knowledge) or (HIV adj3 knowledge) or (STI* adj3 knowledge) or (sex* adj3 knowledge)).tw,id. | 12279 |
| 90 | ("minimum initial service package" or MISP).tw,id. | 3 |
| 91 | mothers/ or adolescent mothers/ or expectant mothers/ | 38299 |
| 92 | ("maternal health" or "maternal welfare" or "maternal child health" or "maternal child welfare" or "maternal health services").tw,id. | 1720 |
| 93 | exp obstetrics/ | 2539 |
| 94 | obstetric*.tw,id. | 5879 |
| 95 | ((labor or labour) adj5 (birth* or breech or childbirth or childbirths or complicat* or difficult or early or easy or induce* or induction or late or obstetric* or onset or pregnan* or present*)).tw,id. | 2082 |
| 96 | pregnancy/ or adolescent pregnancy/ | 23664 |
| 97 | (pregnanc* or "child bearing" or childbearing).tw,id. | 40440 |
| 98 | exp Perinatal Period/ or exp Prenatal Care/ | 4413 |
| 99 | ("prenatal" or "pre natal" or "antenatal" or "ante natal").tw,id. | 21772 |
| 100 | ("perinatal care" or "peri natal care" or "perinatal care" or "peri natal care").tw,id. | 296 |
| 101 | ("peripartum* period*" or "perinatal* period*" or "peri natal* period*").tw,id. | 1498 |
| 102 | exp birth/ | 12878 |
| 103 | (birth or childbirth* or parturition* or "safe delivery" or "safely delivered").tw,id. | 58745 |

| | | |
|---|---|---|
| 104 | (antepartum or "ante partum" or intrapartum or "intra partum").tw,id. | 697 |
| 105 | INTRAPARTUM PERIOD/ | 8 |
| 106 | (stillbirth or stillbirths or stillborn or stillborns or "still birth" or "still births" or "still born" or "still borns").tw,id. | 878 |
| 107 | ("emergency obstetric care" or "basic emergency obstetric care" or "comprehensive emergency obstetric care" or "basic emoc" or "comprehensive emoc").tw,id. | 51 |
| 108 | (emoc or emonc or cemoc or bemoc).tw,id. | 20 |
| 109 | midwifery/ | 1256 |
| 110 | ("birth attendant" or "birth attendants" or midwife or midwives or midwifery or "traditional birth attendant*" or "skilled birth attendant*").tw,id. | 3127 |
| 111 | postnatal period/ or postpartum depression/ | 8217 |
| 112 | (postnatal or "post natal" or postpartum or "post partum" or puerperium or puerperal).tw,id. | 30647 |
| 113 | induced abortion/ or spontaneous abortion/ | 3258 |
| 114 | miscarriage*.tw,id. | 1176 |
| 115 | (abortion or abortions or aborted or aborting or abortus).tw,id. | 5950 |
| 116 | ((unwanted or unintended) adj2 pregnanc*).tw,id. | 1961 |
| 117 | (Abortion adj3 (safe or unsafe or post)).tw,id. | 284 |
| 118 | (Misoprostol or mifepristone or cytotec or mifeprex or mifegymiso or "abortion pill").tw,id. | 326 |
| 119 | family planning/ or birth control/ | 4620 |
| 120 | ("birth interval" or "birth intervals" or "birth spacing" or "birth spacings" or "child spacing" or "child spacings" or "family building" or "family planning" or "pregnancy interval" or "pregnancy intervals").tw,id. | 3244 |
| 121 | "safe motherhood".tw,id. | 105 |

| | | |
|---|---|---|
| 122 | rhythm method/ or tubal ligation/ or vasectomy/ or condoms/ or "sterilization (sex)"/ | 4244 |
| 123 | ("birth regulation" or "birth control*" or "coitus interruptus" or "fertility control" or contracept* or "conception control" or antifertility or anticonception or "fertility control" or "fertilization inhibition" or "inhibition of fertili?ation" or "fertili?ation inhibition" or "inhibition of fertili?ation" or "pregnancy prevent*").tw,id. | 9575 |
| 124 | exp contraceptive devices/ | 5477 |
| 125 | ("cervical cap" or "cervical caps" or "i.u.d." or "intra uterine device*" or "intracervical device" or "intrauterine device*" or "iucd" or "iud" or "iuds" or "progestin implant" or "vaginal diaphragm" or "vaginal diaphragms" or "vaginal ring" or "vaginal shield" or "vaginal rings" or "vaginal shields" or "vaginal sponge" or "vaginal sponges" or "coiled spring*" or condom or condoms or spermicide or "rhythm method" or "calendar method" or "pull out method" or "tubal ligation" or vasectomy or "depo provera").tw,id. | 10175 |
| 126 | (("birth control" or contracept*) adj3 (pill or patch or implant or shot or injection or withdrawal)).tw,id. | 339 |
| 127 | ("morning after pill" or "postcoital antifertility agent" or "postcoital pill" or "plan B" or "emergency contraception").tw,id. | 334 |
| 128 | sexual abstinence/ or virginity/ | 861 |
| 129 | (celibacy or "postpartum abstinence" or "sexual abstinence" or virginity).tw,id. | 1006 |
| 130 | hiv/ | 33583 |
| 131 | ("acquired immune deficiency syndrome virus" or "acquired immunodeficiency syndrome virus" or "aids associated lentivirus" or "aids associated retrovirus" or "aids associated virus" or "aids related virus" or "aids virus" or "aids viruses" or "hiv" or "human immuno deficiency virus" or "human immunodeficiency virus" or "human immunodeficiency viruses" or "aids related illness*").tw,id. | 50632 |
| 132 | ("hiv infection*" or "hiv seropositivit*" or "hiv coinfection*").tw,id. | 11122 |
| 133 | aids/ | 15160 |

| | | |
|---|---|---|
| 134 | ("Antiretroviral therapy" or "antiretroviral treatment" or antiretroviral* or HAART or "anti hiv agent*").tw,id. | 6763 |
| 135 | (pmtct or "prevention of HIV mother to child transmission" or "prevention of mother to child transmission" or "eliminate mother to child transmission" or "prevention of mother to child HIV transmission").tw,id. | 439 |
| 136 | exp Disease Transmission/ | 1849 |
| 137 | "post exposure prophylaxis".tw,id. | 129 |
| 138 | sexually transmitted diseases/ | 4268 |
| 139 | ("sexually transmitted disease*" or "sexually transmitted infection*" or "venereal disease*" or "venereal infection" or "venereal infections" or STIs or STDs).tw,id. | 8830 |
| 140 | gonorrhea/ or syphilis/ | 539 |
| 141 | ("chancroid" or "chancroids" or chlamidiosis or "chlamydia infection*" or "chlamydia trachomatis" or "gonorrhea" or "granuloma inguinale" or "granuloma venereum" or "Klebsiella granulomatis infection" or "Neisseria gonorrhoeae infection" or "syphilis" or "syphilitic disorder" or trichomoniasis or "trichomonas vaginitis").tw,id. | 2240 |
| 142 | herpes genitalis/ or herpes simplex/ or human papillomavirus/ | 2330 |
| 143 | hepatitis/ | 2580 |
| 144 | ("anal wart" or "anal warts" or "anogenital wart" or "anogenital warts" or "condyla acuminatum" or "condylatum acuminatum" or "condyloma accuminatum" or "condyloma acuminata" or "condylomata acuminata" or "genital herpes" or "genital wart" or "genital warts" or "herpes genitalis" or "herpes progenitalis" or "herpes simplex genitalis" or "herpes simplex virus genital infection" or "penile wart" or "penile warts" or "perianal wart" or "perianal warts" or "venereal wart" or "venereal wartsor human papillomavirus*" or HPV or "papillomavirus infection*" or "hepatitis B").tw,id. | 2994 |
| 145 | ("sarcoptic mange" or scabies).tw,id. | 35 |

| | | |
|---|---|---|
| 146 | ("crab lice" or "crab lices" or "crab louse" or "crab louses" or "Pediculus pubis" or "phthirus" or "Phtirus pubis" or "Pthirus pubis" or "pubic lice" or "pubic louse").tw,id. | 2 |
| 147 | partner abuse/ or intimate partner violence/ or domestic violence/ | 19285 |
| 148 | ("partner abuse" or "partner violence" or "wife abuse" or "spouse abuse" or "spousal abuse" or "domestic violence" or "domestic abuse" or "gender based violence" or "gender-based violence" or "GBV" or "IPV" or "sex* based violence").tw,id. | 19805 |
| 149 | sexual abuse/ or rape/ | 24542 |
| 150 | ((abuse* or assault* or violence) adj2 (woman or women)).tw,id. | 6917 |
| 151 | sex offenses/ | 9468 |
| 152 | human trafficking/ | 810 |
| 153 | circumcision/ | 769 |
| 154 | ("coerced intercourse" or "forced intercourse" or "forced prostitution" or "forced sex" or "human trafficking" or "human traffickings" or rape or "sex trafficking" or "sex traffickings" or "sex* abuse*" or "sex* assault*" or "sex* crime*" or "sex* offense" or "sex* offenses" or "sex* slave*" or "sexual aggression" or "sexual bullying" or "sexual coercion" or "sexual exploitation*" or "sexual harassment" or "sexual trauma" or "sexual violence" or "unwanted sex" or "unlawful sex" or "honor killings" or "transactional sex").tw,id. | 45313 |
| 155 | physical abuse/ | 5713 |
| 156 | ("physical* abuse*" or "physical* assault*" or "physical violence" or "psychological violence" or "emotional violence" or "economic violence" or "female circumcision" or "female genital mutilation" or "female genital cutting" or FGM).tw,id. | 12297 |
| 157 | (fistula or fistulas or "genital trauma" or "genital injury" or "vaginal trauma" or "vaginal injury").tw,id. | 533 |
| 158 | preeclampsia/ or obstetrical complications/ or gestational diabetes/ | 1493 |
| 159 | ("toxaemia" or " toxemia" or "hypertension edema proteinuria gestosis" or "pre eclampsia" or "pre eclamptic toxaemia" or "pre eclamptic toxemia" or | 612 |

| | | |
|---|---|---|
| | "preeclamptic toxaemia" or "preeclamptic toxemia" or "pregnancy toxaemia*" or "pregnancy toxemia*" or "toxemia of pregnanc*" or "toxemic pregnancy" or eclampsia or preeclampsia).tw,id. | |
| 160 | ("abnormal labor" or "abnormal labour" or "delayed labor" or "delayed labour" or "inertia uteri" or "labor obstruction" or "labour obstruction" or "obstructed labor" or "obstructed labour" or "uterus inertia" or dystocia or dystocias).tw,id. | 85 |
| 161 | (breech adj2 (present* or position*)).tw,id. | 74 |
| 162 | ("vagina* haemorrhage" or "vagina* hemorrhage" or "vaginal bleeding" or "postpartum hemorrhage" or "post partum hemorrhage" or "uterine hemorrhage").tw,id. | 180 |
| 163 | menstruation/ or menstrual cycle/ or menarche/ | 3704 |
| 164 | ((Menstruation or menarche or menstrual or menses) adj2 (hygiene or bleeding or cycle)).tw,id. | 3505 |
| 165 | human rights/ | 5946 |
| 166 | ((sexual or reproductive or "women's") adj3 rights).tw,id. | 1672 |
| 167 | ("Sexual initiation" or "early sexual debut").tw,id. | 675 |
| 168 | or/84-167 | 313289 |
| 169 | 40 and 50 and 61 and 83 and 168 | 1660 |
| 170 | limit 169 to yr="1990 -Current" | 1644 |

**<u>Database(s): SCOPUS</u>**

**Search Strategy:**

**Humanitarian concept:**

( TITLE-ABS-KEY ( "protected village" OR "refugee camp" ) ) OR ( TITLE-ABS-KEY ( ( camp ) W/3 ( refugee OR transit OR displace* OR temporary OR informal ) ) ) OR ( TITLE-ABS-KEY ( settlement W/3 ( temporary OR informal ) ) ) OR ( ( TITLE-ABS-KEY ( {disasters} OR {emergencies} OR {mass casualty incidents} OR {natural disasters} ) ) OR ( TITLE-ABS-KEY ( disasters OR emergencies OR "mass casualty incidents" OR "natural disasters" ) ) OR ( TITLE-ABS-KEY ( "disaster victims" ) ) OR ( TITLE-ABS-KEY ( ( disaster OR emergenc* ) W/2 victim* ) ) OR ( TITLE-ABS-KEY ( ( disaster* OR catastrophe* ) W/5 ( environment* OR human OR manmade OR "man made" OR nature OR natural OR weather ) ) ) OR ( TITLE-ABS-KEY ( {mass casualty} OR {mass casualties} OR {mass fatalities} OR {mass fatality} ) ) OR ( TITLE-ABS-KEY ( {mass casualty} OR {mass casualties} OR {mass fatalities} OR {mass fatality} ) ) OR ( TITLE-ABS-KEY ( ( crisis OR crises ) W/5 ( environ* OR human OR manmade OR {man made} OR nature OR natural OR weather OR setting* ) ) ) OR ( TITLE-ABS-KEY ( ( crisis OR crises OR conflict ) W/3 ( affected ) ) ) OR ( TITLE-ABS-KEY ( "warfare and armed conflicts" OR {armed conflict*} OR warfare OR {ethnic cleansing} OR genocide OR holocaust OR "war exposure" ) ) OR ( TITLE-ABS-KEY ( {afghan campaign} OR {gulf war} OR {iraq war} OR "war time" OR "wartime" OR "war torn" OR "war affected" OR {insurgency} OR {intra conflict} ) ) OR ( TITLE-ABS-KEY ( ( armed OR zone OR political OR civil OR setting* ) W/3 ( conflict OR conflicts OR attack OR attacks OR war OR wars OR {no fly} ) ) ) OR ( TITLE-ABS-KEY ( fragile W/2 ( state* OR countr* OR nation* OR situation* OR setting* ) ) ) OR ( TITLE-ABS-KEY ( "Post conflict" OR "postconflict" OR "post war" OR "post conflict setting" OR {peacebuilding} OR peacekeeping ) ) OR ( TITLE-ABS-KEY ( war W/2 related ) ) OR ( TITLE-ABS-KEY ( "militant group" OR "militant organization" OR "militant organisation" OR militia OR combatant OR rebel* ) ) OR ( TITLE-ABS-KEY ( {disaster medicine} OR "disaster outbreak" OR {epidemic} OR "disease outbreak" ) ) OR ( TITLE-ABS-KEY ( "Emergency Medical Services" ) ) OR ( TITLE-ABS-KEY ( ( emergency OR emergencies ) W/5 ( environment* OR human OR manmade OR "man made" OR nature OR natural OR weather OR complex ) ) ) OR ( TITLE-ABS-KEY ( famine OR starvation ) ) OR ( TITLE-ABS-KEY ( avalanche OR cyclone OR drought OR earthquake OR flood* OR hurricane OR landslide OR {land slide} OR {land slides} OR mudslide OR {mud slide} OR {mud slides} OR storm OR tornado* OR tsunami OR typhoon OR volcanic OR {volcanic eruption} OR {volcanic ash} OR {volcanic gases} OR rubble ) ) OR ( TITLE-ABS-KEY ( evacuee OR refugee OR squatter OR transients OR {asylum seeker} ) ) OR ( TITLE-ABS-KEY ( ( rescue OR relief OR aid ) W/3 ( plan OR activity OR activities OR agency OR agencies ) ) ) OR ( TITLE-ABS-KEY ( "aid plan" OR "aid work" OR "relief plan" OR "relief work" OR "rescue plan" OR "rescue work" ) ) OR ( TITLE-ABS-KEY ( ( staff OR worker ) W/3 ( relief OR aid OR rescue ) ) ) OR ( TITLE-ABS-KEY ( "humanitarian assistance" ) ) OR ( TITLE-ABS-KEY ( humanitarian W/3 ( aid OR response OR relief OR crisis OR crises OR emergency OR emergencies OR disaster OR disasters ) ) ) OR ( TITLE-ABS-KEY ( humanitarianism OR altruism ) ) OR ( TITLE-ABS-KEY ( {displaced children} OR {displaced child} OR {displaced families} OR {displaced family} OR {displaced individuals} OR {displaced internally} OR {displaced men} OR {displaced people} OR {displaced peoples} OR {displaced person} OR {displaced persons} OR {displaced population} OR {displaced populations} OR {displaced women} OR {displaced adolescent} OR {displaced adolescents} OR {forced displacement} OR {forced displacements} OR {forcibl* displace*} OR {internal displaced} OR {internal displacement}

OR {internally displaced} OR {population displaced} OR {population displacement} OR {forced migration} OR {migrant*} ) ) )

**Population concept:**

( TITLE-ABS-KEY ( displace* W/3 ( child* OR famil* OR men OR wom?n OR individual* OR adolescent* OR people* OR population* OR person* OR girl* OR boy* OR youth ) ) ) OR ( TITLE-ABS-KEY ( adolescen* OR teen* OR youth OR "young adult" OR girl* OR boy* OR "young wom?n" OR "young girl*" OR "young boy*" ) ) OR ( TITLE-ABS-KEY ( "Pregnant Wom?n" OR wom?n ) ) OR ( TITLE-ABS-KEY ( {mother to be} OR mother ) ) OR ( TITLE-ABS-KEY ( female ) ) OR ( TITLE-ABS-KEY ( parturient ) ) OR ( TITLE-ABS-KEY ( sexually AND active ) ) OR ( TITLE-ABS-KEY ( refugee ) ) OR ( TITLE-ABS-KEY ( women W/2 reproductive W/2 age ) )

**LMICs Concept**

( TITLE-ABS-KEY ( "Developing Countries" ) ) OR ( TITLE-ABS-KEY ( africa OR asia OR caribbean OR {West Indies} OR {South America} OR {Latin America} OR {Central America} ) ) OR ( TITLE-ABS-KEY ( fiji OR gabon OR {Gabonese Republic} OR gambia OR gaza OR georgia OR {Georgia Republic} OR {Georgian Republic} OR ghana OR {Gold Coast} OR grenada OR guatemala OR guinea OR bissau OR guiana OR guyana OR haiti OR honduras OR india OR maldives OR indonesia OR iran OR iraq OR jamaica OR jordan OR kazakhstan OR kazakh OR kenya OR kiribati OR korea OR {Democratic People's Republic of Korea} OR kosovo OR kyrgyzstan OR kirghizia OR {Kyrgyz Republic} OR kirghiz OR kirgizstan ) ) OR ( TITLE-ABS-KEY ( {Lao PDR} OR laos OR lebanon OR lesotho OR basutoland OR liberia OR libya OR macedonia OR madagascar OR {Malagasy Republic} OR malaysia OR malaya OR malay OR sabah OR sarawak OR malawi OR nyasaland OR mali OR {Marshall Islands} OR mauritania OR mauritius OR {Agalega Islands} OR mexico OR micronesia OR {Middle East} OR moldova OR moldovia OR moldovian OR mongolia OR montenegro OR morocco OR ifni OR mozambique OR myanmar OR myanma OR burma OR namibia OR nauru OR nepal OR nicaragua OR niger OR nigeria OR pakistan OR {Papua New Guinea} OR palestine OR paraguay OR peru OR philippines OR philipines OR phillipines OR phillippines OR romania OR rumania OR roumania OR russia OR russian OR rwanda OR ruanda OR {Saint Lucia} OR {St Lucia} ) ) OR ( TITLE-ABS-KEY ( {Saint Vincent} OR {St Vincent} OR grenadines OR samoa OR {Samoan Islands} OR {Navigator Island} OR {Navigator Islands} OR {Sao Tome} OR senegal OR serbia OR montenegro OR {sierra leone} OR {sri lanka} OR ceylon OR {solomon islands} OR somalia OR {south Africa} OR sudan OR {South Sudan} OR suriname OR surinam OR swaziland OR eswatini OR syria OR {Syrian Arab Republic} OR tajikistan OR tadzhikistan OR tadjikistan OR tadzhik OR tanzania OR thailand OR togo OR {Togolese Republic} OR tonga OR tunisia OR turkey OR turkmenistan OR turkmen OR tuvalu OR uganda OR ukraine OR ussr OR {Soviet Union} OR {Union of Soviet Socialist Republics} OR uzbekistan OR uzbek OR vanuatu OR {New Hebrides} OR venezuela OR vietnam OR {Viet Nam} OR {West Bank} OR yemen OR yugoslavia OR zambia OR zimbabwe OR rhodesia ) ) OR ( TITLE-ABS-KEY ( afghanistan OR albania OR algeria OR {American Samoa} OR angola OR armenia OR armenian OR azerbaijan OR bangladesh OR benin OR byelarus OR byelorussian OR belarus OR belorussian OR belorussia OR belize OR bhutan OR bolivia OR bosnia OR herzegovina OR hercegovina OR botswana OR brasil OR brazil OR bulgaria OR {Burkina Faso} OR {Burkina Fasso} OR {Upper Volta} OR burundi OR urundi OR cambodia OR {Khmer Republic} OR kampuchea OR cameroon OR cameroons OR cameron OR camerons OR cameroun OR {Cape Verde} OR {Cabo

Verde} OR {Central African Republic} OR chad OR china OR colombia OR comoros OR {Comoro Islands} OR comores OR mayotte OR congo OR zaire OR {Costa Rica} OR {Cote d'Ivoire} OR {Ivory Coast} OR cuba OR djibouti OR {French Somaliland} OR dominica OR {Dominican Republic} OR {East Timor} OR {East Timur} OR {Timor Leste} OR ecuador OR egypt OR {United Arab Republic} OR {El Salvador} OR {Equatorial Guinea} OR eritrea OR ethiopia ) ) OR ( TITLE-ABS-KEY ( ( developing OR "less* developed" OR "under developed" OR underdeveloped OR "middle income" OR "low* income" OR "underserved" OR "under served" OR deprived OR poor ) W/2 ( {countr*} OR {nation?} OR {population?} OR world ) ) ) OR ( TITLE-ABS-KEY ( ( developing OR "less* developed" OR "under developed" OR underdeveloped OR "middle income" OR "low* income" ) W/2 ( economy OR economies ) ) ) OR ( TITLE-ABS-KEY ( low* W/2 ( gdp OR gnp OR {gross domestic} OR {gross national} ) ) ) OR ( TITLE-ABS-KEY ( low W/3 middle W/3 ( {countr*} OR {nation*} ) ) ) OR ( TITLE-ABS-KEY ( lmic OR lmics OR {third world} OR {lami countr*} ) ) OR ( TITLE-ABS-KEY ( "transitional countr*" ) )

**Measurement concept:**

( TITLE-ABS-KEY ( "Health status indicators" ) ) OR ( TITLE-ABS-KEY ( indicator* OR "health indicator" OR evaluation OR tool* OR assessment* OR methodolog* OR standards OR "unmet need" OR coverage ) ) OR ( TITLE-ABS-KEY ( monitor* OR surveillance OR screening ) ) OR ( TITLE-ABS-KEY ( {rapid counting} OR {aerial surveillance} OR {flow monitoring} OR {enumeration} OR {reproductive health assessment toolkit} ) ) OR ( TITLE-ABS-KEY ( ( rapid OR needs ) W/2 ( assessment OR evaluation ) ) ) OR ( TITLE-ABS-KEY ( "data collection tool*" OR "data source*" OR "data collection" OR "needs assessment" ) ) OR ( TITLE-ABS-KEY ( "data collection tool*" OR "data source*" OR "data collection" OR "needs assessment" OR "questionnaire" OR "survey" OR "health survey" OR {contraceptive prevalence survey} OR "health care survey" ) ) OR ( TITLE-ABS-KEY ( scale* OR survey* ) ) OR ( TITLE-ABS-KEY ( index OR indices ) ) OR ( TITLE-ABS-KEY ( {minimum initial service package} OR misp ) ) OR ( TITLE-ABS-KEY ( measure* OR metric* OR method* ) ) OR ( TITLE-ABS-KEY ( ( "health care" ) W/3 ( access OR equity OR delivery OR quality ) ) ) OR ( TITLE-ABS-KEY ( "health access" ) ) OR ( TITLE-ABS-KEY ( "attitude to health" OR "health services research" OR "health status" ) ) OR ( TITLE-ABS-KEY ( morbidity OR {maternal mortality} OR {perinatal mortality} OR prevalence OR incidence ) ) OR ( TITLE-ABS-KEY ( mortality OR {maternal mortality} OR {perinatal mortality} ) ) OR ( TITLE-ABS-KEY ( outcome* W/3 ( health OR measur* OR assess* OR scor* OR index OR indices OR scale* OR monitor* ) ) ) OR ( TITLE-ABS-KEY ( "disease burden" OR {quality adjusted life year} OR {disability adjusted life year} ) )

**SRHR:**

( ( TITLE-ABS-KEY ( gynecology OR "reproductive health" OR "sexual health" OR "sexual and reproductive health" OR "reproductive services" ) ) OR ( TITLE-ABS-KEY ( "family planning education" OR "family planning counselling" OR "family planning instructor" OR "family planning training" OR "sex* education" OR "sex* instruction" OR "sexual health education" OR ( "reproductive health" W/2 knowledge ) OR ( hiv W/2 knowledge ) OR ( sti* W/2 knowledge ) OR ( sex* W/2 knowledge ) ) ) OR ( TITLE-ABS-KEY ( {minimum initial service package} OR misp ) ) OR ( TITLE-ABS-KEY ( "maternal health" OR {maternal welfare} OR "maternal child health" OR {maternal child welfare} OR "maternal health services" ) ) OR ( TITLE-ABS-KEY ( obstetric OR childbirth OR birth OR labor OR labour ) ) OR ( TITLE-ABS-KEY ( ( labor OR labour ) W/5 ( birth OR breech OR childbirth OR complicat* OR difficult* OR early OR easy OR induce* OR induction OR late OR obstetric OR onset OR pregnan* OR present* ) ) ) OR ( TITLE-ABS-KEY ( pregnanc* OR {child bearing} OR childbearing ) ) OR ( TITLE-ABS-KEY ( {prenatal care} OR prenatal OR "pre natal" OR antenatal OR "ante natal" OR {antenatal care} ) ) OR ( TITLE-ABS-KEY ( {peri natal care} OR {perinatal care} OR "perinatal" ) ) OR ( TITLE-ABS-KEY ( {peripartum period} OR {perinatal period} OR peripartum ) ) OR ( TITLE-ABS-KEY ( parturition OR {safe delivery} OR {safely delivered} ) ) OR ( TITLE-ABS-KEY ( antepartum OR "ante partum" OR intrapartum OR "intra partum" ) ) OR ( TITLE-ABS-KEY ( stillbirth OR stillborn OR {still birth} OR {still born} ) ) OR ( TITLE-ABS-KEY ( "emergency obstetric care" OR ( basic W/3 "emergency obstetric care" ) OR ( comprehensive W/3 "emergency obstetric care" ) OR ( basic W/3 emoc ) OR ( comprehensive W/3 emoc ) ) ) OR ( TITLE-ABS-KEY ( emoc OR emonc OR cemoc OR bemoc ) ) OR ( TITLE-ABS-KEY ( midwife OR midwives OR midwifery OR "birth attendant" OR {traditional birth attendant} OR {skilled birth attendant} ) ) OR ( TITLE-ABS-KEY ( postnatal OR {post natal} OR postpartum OR {post partum} OR puerperium OR puerperal OR {postnatal care} OR {postpartum period} ) ) OR ( TITLE-ABS-KEY ( miscarriage ) ) OR ( TITLE-ABS-KEY ( ( abortion ) W/3 ( spontaneous OR induced OR septic OR legal OR therapeutic OR criminal OR illegal OR incomplete OR selective OR medical ) ) ) OR ( TITLE-ABS-KEY ( abortion OR aborted OR aborting OR abortus ) ) OR ( TITLE-ABS-KEY ( ( unwanted OR unintended ) W/3 ( pregnanc* ) ) ) OR ( TITLE-ABS-KEY ( abortion W/3 ( safe OR unsafe OR post ) ) ) OR ( TITLE-ABS-KEY ( misoprostol OR mifepristone OR cytotec OR mifeprex OR mifegymiso OR "abortion pill" ) ) ) OR ( ( TITLE-ABS-KEY ( "birth interval" OR "birth spacing" OR "child spacing" OR "family planning" OR "pregnancy interval" OR "family planning services" ) ) OR ( TITLE-ABS-KEY ( {safe motherhood} ) ) OR ( TITLE-ABS-KEY ( contracepti* OR {coitus interruptus} OR "contraceptive barrier" OR {postcoital contraception} OR {emergency contraception} OR "natural family planning method" OR {contraceptive effectiveness} OR "long acting reversible contraception" OR "reproductive sterilization" OR "oral contracepti*" ) ) OR ( TITLE-ABS-KEY ( {birth regulation} OR "birth control*" OR "fertility control" OR {conception control} OR antifertility OR anticonception OR {fertili?ation inhibition} OR {inhibition of fertili?ation} OR "pregnanc* prevention" ) ) OR ( TITLE-ABS-KEY ( contraceptive AND devices ) ) OR ( TITLE-ABS-KEY ( {cervical cap} OR {i.u.d.} OR {intra uterine device*} OR {intracervical device} OR {intrauterine device*} OR {iucd} OR iud OR iuds OR {progestin implant} OR {vaginal diaphragm} OR {vaginal ring} OR {vaginal shield} OR {vaginal sponge} OR {coiled spring} OR condom OR spermicide OR {rhythm method} OR {calendar method} OR {pull out method} ) ) OR ( TITLE-ABS-KEY ( {tubal ligation} OR vasectomy OR {depo provera} ) ) OR ( TITLE-ABS-KEY ( ( "birth control" OR contracept* ) W/3 ( pill OR patch OR implant OR shot OR injection OR withdrawal ) ) ) OR ( TITLE-ABS-KEY ( {morning after pill} OR {postcoital antifertility agent} OR {postcoital pill} OR {plan B} ) ) OR ( TITLE-ABS-KEY ( celibacy OR {postpartum abstinence} OR "sexual abstinence" OR virginity ) ) OR ( TITLE-ABS-KEY ( {Human immunodeficiency virus} OR {HIV} OR {HIV-1} OR {HIV-2} ) ) or#149 OR ( TITLE-ABS-KEY ( "hiv infection*" OR "hiv seropositivit*" OR "hiv coinfection*" OR "human immunodeficiency virus infection" ) ) OR ( TITLE-ABS-KEY ( {aids} OR {acquired immune deficiency syndrome*} OR {acquired immuno deficiency syndrome*} OR {acquired immunodeficiency syndrome} OR "aids related opportunistic infection*" ) ) OR ( TITLE-ABS-KEY ( ( "vertical transmission" ) W/3 ( "infectious disease*" OR disease* ) ) ) OR ( TITLE-ABS-KEY ( "post exposure prophylaxis" OR ( hiv W/3 pep ) ) ) OR ( TITLE-ABS-KEY ( {Antiretroviral therapy} OR {antiretroviral treatment} OR antiretroviral* OR haart OR {anti hiv agent} OR {highly active antiretroviral therapy} ) ) OR ( TITLE-ABS-KEY ( pmtct OR {prevention of HIV mother to child transmission} OR {prevention of mother to child transmission} OR {eliminate mother to child transmission} OR "prevention of mother to child HIV transmission" ) ) OR ( TITLE-ABS-KEY ( "sexually transmitted disease" OR "sexually transmitted infection" OR "venereal disease" OR "venereal infection" ) ) OR ( TITLE-ABS-KEY ( "bacterial sexually transmitted disease" OR chancroid OR "chlamydia infection" OR "chlamydia trachomatis" OR gonorrhea OR syphilis OR "trichomonas vaginitis" OR "vaginal trichomoniasis" OR {granuloma inguinale} OR {granuloma venereum} OR {Klebsiella granulomatis infection} OR {Neisseria gonorrhoeae infection} OR {syphilitic disorder} OR trichomoniasis ) ) OR ( TITLE-ABS-KEY ( "viral sexually transmitted diseases" OR {herpes genitalis} OR {Condylomata Acuminata} OR "Hepatitis B" OR "papillomavirus infections" ) ) OR ( TITLE-ABS-KEY ( {anal wart} OR {anal warts} OR {anogenital wart} OR {anogenital warts} OR {condyla acuminatum} OR {condylatum acuminatum} OR {condyloma accuminatum} OR {condyloma acuminate} OR {condylomata acuminate} OR {genital herpes} OR {genital wart} OR {genital warts} OR {herpes progenitalis} OR {herpes simplex genitalis} OR {herpes simplex virus genital infection} OR {penile wart} OR {penile warts} OR {perianal wart} OR {perianal warts} OR {venereal wart} OR {venereal warts} OR {human papillomavirus} OR {papillomavirus infection} OR {hepatitis B} ) ) OR ( TITLE-ABS-KEY ( {sarcoptic mange} OR scabies OR "phthirus" ) ) OR ( TITLE-ABS-KEY ( {crab lice} OR {crab lices} OR {crab louse} OR {crab louses} OR {Pediculus pubis} OR {phthirus} OR {Phtirus pubis} OR {Pthirus pubis} OR {pubic lice} OR {pubic louse} ) ) ) OR ( ( TITLE-ABS-KEY ( {Intimate Partner Abuse} OR {Spouse abuse} OR {Domestic Violence} OR {intimate partner violence} ) ) OR ( TITLE-ABS-KEY ( {partner abuse} OR {partner violence} OR {wife abuse} OR {spouse abuse} OR {spousal abuse} OR "domestic violence" OR {domestic abuse} OR {gender based violence} OR {gender-based violence} OR "sex* based violence" ) ) OR ( TITLE-ABS-KEY ( ( abuse* OR assault* OR violence ) W/2 ( woman OR women ) ) ) OR ( TITLE-ABS-KEY ( "Sex offenses" OR "sexual assault" OR "sexual crime" OR "sexual abuse" OR {Human Trafficking} OR rape OR "female circumcision" OR "female genital mutilation" ) ) OR ( TITLE-ABS-KEY ( {coerced intercourse} OR {forced intercourse} OR {forced prostitution} OR {forced sex} OR {sex trafficking} OR {sex traffickings} OR "sex* slave" OR {sexual aggression} OR {sexual bullying} OR {sexual coercion} OR {sexual exploitation} OR {sexual harassment} OR {sexual trauma} OR {sexual violence} OR {unwanted sex} OR {unlawful sex} OR {honor killings} OR {transactional sex} ) ) OR ( TITLE-ABS-KEY ( {physical abuse} OR {physical assault} OR {physical violence} OR {psychological violence} OR {emotional violence} OR {economic violence} OR {female circumcision} OR {female genital mutilation} OR {female genital cutting} ) ) OR ( TITLE-ABS-KEY ( fistula OR {vaginal fistula} OR "urinary tract fistula" OR {rectovaginal fistula} OR "genital trauma" OR "genital injury" OR "vaginal trauma" OR "vaginal injury" ) ) OR ( TITLE-ABS-KEY ( "pregnancy complications" OR {gestational diabetes} OR "pregnancy induced hypertension" OR "obstetric labor complications" OR {pregnancy diabetes mellitus} OR {maternal hypertension} ) ) OR ( TITLE-ABS-KEY ( toxemia OR {hypertension edema proteinuria gestosis} OR {pre eclampsia} OR {pre eclamptic toxaemia} OR {pre eclamptic toxemia} OR {preeclamptic toxaemia} OR {preeclamptic toxemia} OR {pregnancy toxaemia} OR {pregnancy toxemia} OR {toxemia of pregnancy} OR {toxemic pregnancy} OR eclampsia OR preeclampsia OR pre-eclampsia ) ) OR ( TITLE-ABS-KEY ( "abnormal labor" OR "abnormal labour" OR "delayed labor" OR "delayed labour" OR {inertia uteri} OR "labor obstruction" OR "labour obstruction" OR "obstructed labor" OR "obstructed labour" OR {uterus inertia} OR dystocia OR "breech presentation" ) ) OR ( TITLE-ABS-KEY ( "postpartum hemorrhage" OR "uterine hemorrhage" OR "vaginal hemorrhage" OR "vaginal bleeding" ) ) OR ( TITLE-ABS-KEY ( ( menstruation OR menarche OR menstrual OR menses ) W/2 ( hygiene OR bleeding OR cycle ) ) ) OR ( TITLE-ABS-KEY ( menstruation OR {menstrual cycle} ) ) OR ( TITLE-ABS-KEY ( "Reproductive rights"

OR "women's rights" ) ) OR ( TITLE-ABS-KEY ( ( sexual OR reproductive OR women's ) W/3 ( rights ) ) ) )

- AND NOT INDEX (MEDLINE)
- AND ( LIMIT-TO ( PUBYEAR , 2019 ) OR LIMIT-TO ( PUBYEAR , 2018 ) OR LIMIT-TO ( PUBYEAR , 2017 ) OR LIMIT-TO ( PUBYEAR , 2016 ) OR LIMIT-TO ( PUBYEAR , 2015 ) OR LIMIT-TO ( PUBYEAR , 2014 ) OR LIMIT-TO ( PUBYEAR , 2013 ) OR LIMIT-TO ( PUBYEAR , 2012 ) OR LIMIT-TO ( PUBYEAR , 2011 ) OR LIMIT-TO ( PUBYEAR , 2010 ) OR LIMIT-TO ( PUBYEAR , 2009 ) OR LIMIT-TO ( PUBYEAR , 2008 ) OR LIMIT-TO ( PUBYEAR , 2007 ) OR LIMIT-TO ( PUBYEAR , 2006 ) OR LIMIT-TO ( PUBYEAR , 2005 ) OR LIMIT-TO ( PUBYEAR , 2004 ) OR LIMIT-TO ( PUBYEAR , 2003 ) OR LIMIT-TO ( PUBYEAR , 2002 ) OR LIMIT-TO ( PUBYEAR , 2001 ) OR LIMIT-TO ( PUBYEAR , 2000 ) OR LIMIT-TO ( PUBYEAR , 1999 ) OR LIMIT-TO ( PUBYEAR , 1998 ) OR LIMIT-TO ( PUBYEAR , 1997 ) OR LIMIT-TO ( PUBYEAR , 1996 ) OR LIMIT-TO ( PUBYEAR , 1995 ) OR LIMIT-TO ( PUBYEAR , 1994 ) OR LIMIT-TO ( PUBYEAR , 1993 ) OR LIMIT-TO ( PUBYEAR , 1992 ) OR LIMIT-TO ( PUBYEAR , 1991 ) OR LIMIT-TO ( PUBYEAR , 1990 ) )
