## Additional file 2 for "Tools for measuring sexual and reproductive health and rights (SRHR) indicators in humanitarian settings"

### Additional file 2: Inclusion and Exclusion Criteria

| | Inclusion Criteria | Exclusion Criteria |
|---|---|---|
| **Population of Interest** | Adolescent girls (10 – 19 years), women of reproductive age (WRA) (15-49 years), older women (49+, if the study population also contains WRA) | Males (unless the outcomes reported are on women), very young female children (<10 years), military personnel or groups other than beneficiaries (i.e. health workers, NGO workers) |
| **Intervention** | Sexual and reproductive health and rights (SRHR) interventions (general SRH, HIV, STIs, gender-based violence, family planning) | Interventions unrelated to SRHR |
| | Maternal health interventions (pregnancy, abortion, antenatal care, labor and delivery, postnatal care) | |
| **Outcomes of Interest** | Quantitative indicators (with reported outcomes) and their corresponding measurement tools, data sources, and data collection methodologies | • Quantitative indicators that do not report outcomes, or provide a methodology for how the data were collected<br>• Qualitative outcomes<br>• Outcomes based on mathematical modelling of descriptive statistics (i.e. regression coefficients, odds ratios, relative risk etc.)<br>• Health systems data with no indicator denominator |
| **Situation/ Setting** | Low- and middle-income countries (LMICs) that have experienced (within 5 years of data collection) one or more of the following humanitarian crises: conflict, epidemic (i.e., Ebola or Zika), or natural disaster (affecting a population greater than 1000) | High-income countries |
| | LMICs hosting displaced population(s) affected by one or more of the humanitarian crises of interest (text must specifically mention a refugee/displaced population living in a LMIC if the host country is not experiencing a conflict/natural disaster) | • LMICs that are not affected by one or more of the humanitarian crises of interest, nor mention hosting a refugee or displaced population from other LMICs<br>• LMICs that have only experienced HIV as an epidemic<br>• High-income countries hosting refugee/displaced populations |
| **Type of Evidence** | Reports, indexed literature, reviews, conference or other abstracts with details about the methodology | Blogs, web pages, dissertations, pamphlets/fact sheets with no methodology reported, financial reports/programming budgets |
| **Time** | Any study published from 2004 — present | Any study published before 2004 |
| **Language** | English | Other languages with no translation available |
