## Additional file 3 for "Tools for measuring sexual and reproductive health and rights (SRHR) indicators in humanitarian settings"

**Additional file 3: Rayyan Screening Tool**

Rayyan uses a support vector machine supervised learning model and features such as unigrams, bigrams and Mesh Terms to rank unscreened titles and abstracts based on their probability of inclusion (1) . After generating a predicted inclusion rating on a five-point scale using a minimal set of 50 pre-screened abstracts, Rayyan uses active learning to inform and update its ranking of abstracts for screening. We chose to use Rayyan as machine learning streamlines the screening process and reduces the workload by minimizing the number of publications that need to be screened.  Two reviewers screened the ranked abstracts, and conflicts were resolved by a third reviewer.

Rayyan does not currently have a stopping feature that allows one to decide when the literature review is complete or nearly complete. In order to determine a stopping point, we set up a training and validation dataset, a-posteriori, using an 80% and 20% split respectively. A new Rayyan review was trained on 80% (n=6648) of the screened abstracts at the time, and the classifier was used to predict the rank of the withheld 20% (n=1662). We identified that a stopping point equal to or greater than Rayyan's third point on their five-point ranking scale corresponded in the validation dataset to a sensitivity of 96.4%, a specificity of 68.4%, an accuracy of 72.1%, and a precision of 31.9%. A sensitivity of 95% and above was acceptable to the authors of this scoping review.

In order to ensure that we had a high sensitivity, we ran a second algorithm to predict abstract inclusion classification. This classifier was a neural network algorithm developed using the deep learning library fastai in python (2) . The classifier was trained on all the screened literature from Rayyan. The highest ranked abstracts with a probability of inclusion of more than 50% were reviewed by two screeners. Two hundred and sixty-six additional abstracts were reviewed using this algorithm and 50 were included.

Overall, we screened 9875 of the 23,608 titles and abstracts from the pulled indexed literature. At our estimated recall of 96.4%, our work saved over random sampling at 53.17%.

1. Ouzzani M, Hammady H, Fedorowicz Z, Elmagarmid A. Rayyan - a web and mobile app for systematic reviews. Systematic Reviews. 2016;5(1):210.
2. Howard J, Gugger S. Fastai: A Layered API for Deep Learning. Information. 2020;11(2):108.
