## Additional file 4 for "Tools for measuring sexual and reproductive health and rights (SRHR) indicators in humanitarian settings"

### Additional file 4: Data Collection Toolkits and Surveys

| Measurement Tool (Standard Name) | Organization/ Author | Toolkit Topic/Domain | Population Type | Humanitarian Specific (Y/N) | # of Studies |
|---|---|---|---|---|---|
| 32-item Gender-Based Violence Survey | Sousa | GBV | Women | N | 1 |
| A Guide to Monitoring and Evaluating Adolescent Health Programs | Sullivan | ANC, FP, HIV & STIs | WRA | N | 1 |
| Adherence Questionnaire | Adult AIDS Clinical Trials Group (AACTG) | HIV & STIs | HIV Positive Individuals | N | 1 |
| Adolescent Sexual & Reproductive Health Assessment Toolkit for Humanitarian Settings | Catani, Ivanova | FP, GBV, HIV & STIs, Menstruation & Gynecological Health | Adolescent girls, Children | Y | 2 |
| Adolescents' Sexual Behaviour Questionnaire | Benner | FP, Menstruation & Gynecological Health | Adolescent girls, Young women | N | 1 |
| Assessment Screen to Identify Survivors Toolkit for Gender-Based Violence (ASIST-GBV) | Catani, Vu | GBV | Adolescent girls, Children, Women | Y | 3 |
| CARE Questionnaire | CARE International, Catani | GBV | Adolescents, Adults, Children | Y | 2 |
| CDC Reproductive Health Assessment Toolkit for Conflict-Affected Women | Adam, Al- Maharma, Anwar, Casey, Catani, Falb, Hynes, Ivanova, McGinn, Okanlawon, Raheel, Reese Masterson, Seyife, Simsek, Sipsma, Tanabe, The Reproductive Health Response in Conflict Consortium, UNFPA, Usta, Wako, Women's refugee commission | Abortion, ANC, FP, GBV, HIV & STIs, Maternal Health, Menstruation & Gynecological Health, Obstetric Care (Delivery) | Adolescent girls, Adolescents, Children, Pregnant & postpartum women, WRA, Young women | Y | 26 |
| Child Soldiers Trauma Questionnaire (CSTQ)* | Klasen | GBV | Adolescent girls | Y | 1 |
| Child War Trauma Questionnaire (CWTQ) | Betancourt, Klasen | GBV | Adolescent girls, Adolescents, Girls | Y | 3 |

| | | | | | |
|---|---|---|---|---|---|
| Chinese CDC National Institute of Nutrition and Food Safety Nutritional Survey* | Dong | Maternal Health, Menstruation & Gynecological Health | WRA | N | 1 |
| Demographic and Health Survey (DHS) | Alemayehu, Bhandari, Blanc, Casey, Chukwumalu, Curry, Gebrecherkos, Gizelis, Green, Husseini, Hynes, Kelly , Meiksin, Metheny, Mukunya, Okanlawon, Pierce, Tatah, The Reproductive Health Response in Conflict Consortium, UNFPA, Vyas, Yaya | Abortion, ANC, FP, GBV, HIV & STIs, Maternal Health, Menstruation & Gynecological Health, Obstetric Care (Delivery) | Adolescent girls, Pregnant & postpartum women, Survivors of violence, Women, WRA, Young women | N | 22 |
| East-Timor Gender-Based Violence Survey | Reproductive Health Response in Conflict Consortium (RHRC) | GBV | WRA | N | 1 |
| Emergency Obstetric Care Needs Assessment Toolkit | Kim | Maternal Mortality, Obstetric Care (Delivery) | WRA | N | 1 |
| Ethiopian Health and Nutrition Research Institute Survey* | Getachew | ANC, Maternal Health | Pregnant & postpartum women | N | 1 |
| Focus Group Discussion Tool containing GBV Assessment Toolkit* | ALNAP-Danish Refugee Council, Catani | FP, GBV | Adolescents, Adults, Children | Y | 2 |
| Gender Needs Assessment (GNA) Survey* | UNIFEM | HIV & STIs, Maternal Health, Obstetric Care (Delivery) | Adolescent girls, Pregnant & postpartum women, Women | Y | 1 |
| Gender-Based Violence Tools Manual in Conflict-Affected Settings | Reese Masterson, Usta | Abortion, ANC, FP, GBV, Maternal Health, Menstruation & Gynecological Health, Obstetric Care (Delivery) | WRA | Y | 3 |
| Georgia Reproductive Health Survey | Doliashvili | Abortion, FP, HIV & STIs, Menstruation & Gynecological Health | WRA | N | 1 |

| | | | | | |
|---|---|---|---|---|---|
| Gulu Sexual Health Study* | Duff | FP, GBV, HIV & STIs | Adolescent girls, Women | N | 2 |
| HIV Behavioral Surveillance Surveys | Catani, Harrison, UNHCR | FP, GBV, HIV & STIs | Adolescent girls, Adolescents, Adults, Children, Women | Y | 3 |
| HIV Knowledge Questionnaire (HIV KQ-18)* | Logie, Ssebunya | FP, HIV & STIs | Adolescent girls, Adolescents, Women | N | 3 |
| Health Access and Utilization Survey Among Non-Camp Syrian Refugees | Torun | ANC, FP, HIV & STIs, Obstetric Care (Delivery) | Pregnant & postpartum women, WRA | Y | 1 |
| Home-Based Life Saving Skills Complication Audit Form* | Lori | Obstetric Care (Delivery) | Pregnant & postpartum women, WRA | N | 1 |
| Household Survey Manual: Diarrhoea and Acute Respiratory Infections | Catani, de Jong, Kaz | GBV | Adolescents, Adults, Children | N | 2 |
| Human Rights History Survey | Graduate Institute Geneva | GBV | Women | Y | 1 |
| Human Rights and Sexual Violence Survey* | Catani, Hynes, The Reproductive Health Response in Conflict Consortium | GBV, Menstruation & Gynecological Health | Adolescents, Children, WRA | Y | 3 |
| IASC Guidelines for Gender-Based Violence Interventions in Humanitarian Settings | Usta | GBV | WRA | Y | 1 |
| ISPCAN Child Abuse Screening Tools | Catani, Stark , Usta | GBV | Adolescent girls, Adolescents, Children | N | 3 |
| International Men and Gender Equality Survey (IMAGES) | Catani, Promundo | Abortion, ANC, FP, GBV, HIV & STIs | Adults, Women | N | 4 |
| Intimate Partner Violence Assessment Questionnaire | Kinyanda | GBV, Menstruation & Gynecological Health | Adolescents, Adults | N | 2 |
| Johns Hopkins and Red Cross/Red Crescent Public Health Guide for Emergencies | Djafri | ANC, FP, Maternal Health, Maternal | WRA | Y | 1 |

| | | | | | |
|---|---|---|---|---|---|
| | | Mortality, Obstetric Care (Delivery) | | | |
| Knowledge, Attitudes & Practice Surveys: Zika Virus Disease & Potential Complications | Brissett | Maternal health | Adults | Y | 1 |
| Knowledge, Practice, & Coverage Survey (KPC) Tool | Edmond | ANC, Maternal Health, Obstetric Care (Delivery) | Pregnant & postpartum women | N | 1 |
| LSHTM Violence and Health Among Women Asylum Seekers | Shuman | GBV | Women | Y | 1 |
| Living Conditions Household Cross Sectional Survey* | Khawaja | GBV | Adolescents, Adults | N | 2 |
| Malaria Indicator Survey | Eyobo | ANC, Maternal Health | WRA | N | 1 |
| Maternal Postpartum Quality of Life Questionnaire | Hammoudeh | FP, Maternal Health, Obstetric Care (Delivery) | Women | N | 1 |
| Minimum Initial Service Package for Reproductive Health | Qayum | ANC, Maternal Health, Menstruation & Gynecological Health, Obstetric Care (Delivery) | Pregnant & postpartum women | Y | 1 |
| Multi-Cluster Rapid Assessment Mechanism (McRAM): Community & Household Survey | UNIFEM | ANC | Adolescent girls, Women | N | 1 |
| Multiple Indicator Cluster Survey (MICS) | Benange, Curry, Decker, Pham | ANC, FP, Maternal Health, Obstetric Care (Delivery), | Pregnant & postpartum women, WRA | N | 5 |
| National Violence Against Women Survey | Catani, Sloand | GBV | Adolescent girls, Adolescents, Children | N | 2 |
| Neighbourhood Method* | Parcesepe | GBV | Women | Y | 1 |
| NorVold Domestic Abuse Questionnaire (NORAQ)* | Al-Shdayfat | GBV | Women | N | 1 |
| Palestinian Central Bureau of Statistics (PCBS) Domestic Violence Survey | Assaf | GBV | WRA | N | 1 |
| Palestinian Family Survey | Kitabayashi | ANC, Maternal Health, Obstetric Care (Delivery) | WRA | Y | 1 |

| | | | | | |
|---|---|---|---|---|---|
| Priorities for Local AIDS Control Efforts (PLACE) Toolkit | Catani, Okigbo | GBV | Adolescent girls, Adolescents, Children, Young women | N | 2 |
| Quick Investigation of Quality | Sullivan | ANC, FP, HIV & STIs | WRA | N | 1 |
| Reproductive Health Response in Conflict Consortium Toolkit | Adam, Usta, Westhoff | Abortion, ANC, FP, GBV, Maternal Health, Menstruation & Gynecological Health, Obstetric Care (Delivery) | Adults, WRA | Y | 4 |
| Safe Motherhood Questionnaire | Jhpiego Maternal and Neonatal Health Program | ANC, Obstetric Care (Delivery) | Adolescent girls, Pregnant and postpartum women, Young women | N | 1 |
| Saferworld Gender Analysis of Conflict Toolkit | Oxfam | GBV | WRA | Y | 1 |
| Sexual Experiences-Victimization Survey | Catania, Sloand | GBV | Adolescents, Adolescent girls, Children | N | 2 |
| Sphere Minimum Standards and Indicators for Humanitarian Response | Qayum | Maternal Health | Adults | Y | 1 |
| Standardized Cervical Cancer Screening Form of Uganda's Ministry of Health | Izudi | HIV & STIs | HIV positive individuals | Y | 1 |
| Survey of War-Affected Youth (SWAY)* | Annan | FP, GBV, HIV & STIs | Adolescent girls, Adolescents, Young adults, Young women | Y | 1 |
| Tanzania Violence Against Children Survey | Wirtz | GBV | Adolescent girls, Women | N | 1 |
| The Survey of Living Conditions Among Palestinian Refugees in Lebanon (LIPRIL)* | Khawaja | GBV | Adolescent girls, Women | N | 1 |
| UNICEF Female Genital Mutilation Study | Wirtz | GBV | Adolescent girls, Women | N | 1 |

| | | | | | |
|---|---|---|---|---|---|
| USAID BASICS Healthy Timing and Spacing of Pregnancy Toolkit | Morris | FP | WRA | N | 1 |
| Violence Against Children Survey (VACS) | Catani, Gilbert, Stark, Sumner | GBV | Adolescent girls, Adolescents, Children, Young adults, Young women | N | 4 |
| WHO Female Genital Mutilation Multi-Country Study | Wirtz | GBV | Adolescent girls, Women | N | 1 |
| WHO Multi-Country Study on Women's Health and Domestic Violence Against Women | Al-modallal, Bhardwaj, Catani, Delkhosh, Erickson, Falb, Gibbs, Gupta, Hossain, Hynes, IRC, Mootz, Murphy, Shuman, Silove, The reproductive health response in conflict consortium, Wirtz | FP, GBV, HIV & STIs, Menstruation & Gynecological Health, Obstetric Care (Delivery) | Adolescent girls, Adolescents, Children, Survivors of violence, Women, WRA, Young adults | N | 18 |
| WHO Reproductive Health Indicators Guideline | Djafri | ANC, FP, Maternal Health, Maternal Mortality, Obstetric Care (Delivery) | WRA | N | 1 |
| WHO Safe Motherhood Needs Assessment Tool | Hoogenboom | ANC, HIV & STIs, Obstetric Care (Delivery) | Pregnant & postpartum women | N | 1 |
| Women's Health Assessment Form | Lash | Menstruation & Gynecological Health | WRA | N | 1 |
| Total | | | | | |

ANC = Antenatal Care, FP = Family Planning, GBV = Gender-Based Violence, STI = Sexually Transmitted Infections, WRA = Women of Reproductive Age
