## Additional file 5 for "Tools for measuring sexual and reproductive health and rights (SRHR) indicators in humanitarian settings"

**Additional file 5: Toolkits and Surveys and Standard Indicators**

| Measurement Tool (Standard Name) | Standard Indicators Included | Count of Standard Indicators |
|---|---|---|
| 32-item Gender-Based Violence Survey | Intimate partner violence prevalence | 1 |
| A Guide to Monitoring and Evaluating Adolescent Health Programs | Antenatal care coverage | 1 |
| Adherence Questionnaire | None | 0 |
| Adolescent Sexual & Reproductive Health Assessment Toolkit for Humanitarian Settings | Knowledge of HIV-related preventative practices, Prevalence of female genital mutilation/cutting | 2 |
| Adolescents' Sexual Behaviour Questionnaire | Sexual and reproductive health (SRH) knowledge (ages 15-24) | 1 |
| Assessment Screen to Identify Survivors Toolkit for Gender-Based Violence (ASIST-GBV) | Non-partner sexual violence prevalence | 1 |
| CARE Questionnaire | None | 0 |
| CDC Reproductive Health Assessment Toolkit for Conflict-Affected Women | Antenatal care coverage, Antenatal care (4+ visits), Antenatal care: Had blood pressure measured, Contraceptive prevalence, HIV prevalence rate, Intermittent preventative treatment for malaria during pregnancy, Intimate partner violence prevalence, Iron supplementation during pregnancy, Knowledge of HIV-related preventive practices, Neonatal tetanus protection, Percentage of ANC clients screened for syphilis, Prevalence of female genital mutilation/cutting, Perinatal mortality rate, Prevalence of positive syphilis serology in pregnant women, Proportion of births attended by a skilled birth attendant, Proportion of births delivered in a health facility, Protection against HIV at last high risk contact, Sexually transmitted infections (STIs) incidence rate, Stillbirth rate, Timing of first ANC visit | 20 |
| Child Soldiers Trauma Questionnaire (CSTQ)* | None | 0 |
| Child War Trauma Questionnaire (CWTQ) | None | 0 |
| Chinese CDC National Institute of Nutrition and Food Safety Nutritional Survey* | Prevalence of anemia in women 15-49, by age & pregnancy status | 1 |
| Demographic and Health Survey (DHS) | Antenatal care coverage, Antenatal care (4+ visits), Antenatal care: Had blood pressure measured, C-section rate, Contraceptive prevalence, HIV prevalence rate, Intermittent preventative treatment for malaria during pregnancy, Intimate partner violence prevalence, Iron supplementation during pregnancy, Malaria test positivity rate, Neonatal tetanus protection, Percentage of ANC clients screened for syphilis, Perinatal mortality rate, Postnatal care for mothers, | 21 |

| | | |
|---|---|---|
| | Prevalence of anemia in women 15-49, by age & pregnancy status, Prevalence of positive syphilis serology in pregnant women, Proportion of births attended by skilled birth attendant, Proportion of births delivered in a health facility, Protection against HIV at last high risk contact, Sexually transmitted infections (STIs) incidence rate, Timing of first ANC visit | |
| East-Timor Gender-Based Violence Survey | Intimate partner violence prevalence | 6 |
| Emergency Obstetric Care Needs Assessment Toolkit | Availability of basic essential obstetric care, C-section rate | 2 |
| Ethiopian Health and Nutrition Research Institute Survey* | Antenatal care coverage, Antenatal care (4+ visits) | 2 |
| Focus Group Discussion Tool containing GBV Assessment Toolkit* | Contraceptive prevalence | 1 |
| Gender Needs Assessment (GNA) Survey* | Perinatal mortality rate, Sexually transmitted infections (STIs) incidence rate | 2 |
| Gender-Based Violence Tools Manual in Conflict-Affected Settings | Antenatal care coverage, Contraceptive prevalence, Perinatal mortality rate, Proportion of births delivered in a health facility | 4 |
| Georgia Reproductive Health Survey | Contraceptive prevalence, Sexually transmitted infections (STIs) incidence rate | 2 |
| Gulu Sexual Health Study* | Contraceptive prevalence, HIV prevalence rate | 2 |
| HIV Behavioral Surveillance Surveys | Contraceptive prevalence, Non-partner sexual violence prevalence, Protection against HIV at last high-risk contact, Sexually transmitted infections (STIs) incidence rate | 4 |
| HIV Knowledge Questionnaire (HIV KQ-18)* | Contraceptive prevalence, HIV prevalence rate | 2 |
| Health Access and Utilization Survey Among Non-Camp Syrian Refugees | Antenatal care coverage, C-section rate, Iron supplementation during pregnancy, Perinatal mortality rate, Proportion of births delivered in a health facility, Sexually transmitted infections (STIs) incidence rate | 6 |
| Home-Based Life Saving Skills Complication Audit Form* | None | 0 |
| Household Survey Manual: Diarrhoea and Acute Respiratory Infections | None | 0 |
| Human Rights and Sexual Violence Survey* | Intimate partner violence prevalence | 1 |
| Human Rights History Survey | Sexually transmitted infections (STIs) incidence rate | 1 |
| IASC Guidelines for Gender-Based Violence Interventions in Humanitarian Settings | None | 0 |
| ISPCAN Child Abuse Screening Tools | Non-partner sexual violence prevalence, Sexual violence against children | 2 |
| International Men and Gender Equality Survey (IMAGES) | Antenatal care coverage, Contraceptive prevalence, Intimate partner violence prevalence, Sexually transmitted infections (STIs) incidence rate | 4 |
| Intimate Partner Violence Assessment Questionnaire | None | 0 |

| | | |
|---|---|---|
| Johns Hopkins and Red Cross/Red Crescent Public Health Guide for Emergencies | Antenatal care coverage, Contraceptive prevalence, Perinatal mortality rate, Proportion of births attended by skilled birth attendant, Stillbirth rate | 5 |
| Knowledge, Attitudes & Practice Surveys: Zika Virus Disease & Potential Complications | None | 0 |
| Knowledge, Practice, & Coverage Survey (KPC) Tool | Antenatal care coverage, Availability of basic essential obstetric care, Proportion of births attended by skilled birth attendant, Proportion of births delivered in a health facility | 4 |
| LSHTM Violence and Health Among Women Asylum Seekers | Intimate partner violence prevalence | 1 |
| Living Conditions Household Cross Sectional Survey* | None | 0 |
| Malaria Indicator Survey | Intermittent preventative treatment for malaria during pregnancy, Use of insecticide treated nets (ITNs) | 2 |
| Maternal Postpartum Quality of Life Questionnaire | Contraceptive prevalence, Stillbirth rate | 2 |
| Minimum Initial Service Package for Reproductive Health | None | 0 |
| Multi-Cluster Rapid Assessment Mechanism (McRAM): Community & Household Survey | Antenatal care coverage | 1 |
| Multiple Indicator Cluster Survey (MICS) | Antenatal care coverage, Antenatal care (4+ visits), Contraceptive prevalence, Iron supplementation during pregnancy, Neonatal tetanus protection, Proportion of births attended by skilled birth attendant | 6 |
| National Violence Against Women Survey | None | 0 |
| Neighbourhood Method* | None | 0 |
| NorVold Domestic Abuse Questionnaire (NORAQ)* | None | 1 |
| Palestinian Central Bureau of Statistics (PCBS) Domestic Violence Survey | None | 0 |
| Palestinian Family Survey | Antenatal care coverage, Antenatal care (4+ visits), Postnatal care for mothers, Proportion of births delivered in a health facility | 4 |
| Priorities for Local AIDS Control Efforts (PLACE) Toolkit | None | 0 |
| Quick Investigation of Quality | Antenatal care coverage | 1 |
| Reproductive Health Response in Conflict Consortium Toolkit | Antenatal care coverage, Contraceptive prevalence, Proportion of births delivered in a health facility | 3 |
| Safe Motherhood Questionnaire | None | 0 |
| Saferworld Gender Analysis of Conflict Toolkit | None | 0 |
| Sexual Experiences-Victimization Survey | None | 0 |

| | | |
|---|---|---|
| Sphere Minimum Standards and Indicators for Humanitarian Response | None | 0 |
| Standardized Cervical Cancer Screening Form of Uganda's Ministry of Health | HIV prevalence rate, Sexually transmitted infections (STIs) incidence rate | 2 |
| Survey of War-Affected Youth (SWAY)* | Contraceptive prevalence | 1 |
| Tanzania Violence Against Children Survey | Sexual violence against children | 1 |
| The Survey of Living Conditions Among Palestinian Refugees in Lebanon (LIPRIL)* | Intimate partner violence prevalence | 1 |
| UNICEF Female Genital Mutilation Study | Sexual violence against children | 1 |
| USAID BASICS Healthy Timing and Spacing of Pregnancy Toolkit | None | 0 |
| Violence Against Children Survey (VACS) | Non-partner sexual violence prevalence, Sexual violence against children | 2 |
| WHO Female Genital Mutilation Multi-Country Study | Sexual violence against children | 1 |
| WHO Multi-Country Study on Women's Health and Domestic Violence Against Women | C-section rate, Contraceptive prevalence, HIV prevalence rate, Intimate partner violence prevalence, Non-partner sexual violence prevalence | 5 |
| WHO Reproductive Health Indicators Guideline | Antenatal care coverage, Contraceptive prevalence, Perinatal mortality rate, Proportion of births attended by skilled birth attendant, Stillbirth rate | 5 |
| WHO Safe Motherhood Needs Assessment Tool | C-section rate, Percentage of ANC clients screened for syphilis, Intermittent preventative treatment for malaria during pregnancy | 3 |
| Women's Health Assessment Form | None | 0 |
