## Additional file 6 for "Tools for measuring sexual and reproductive health and rights (SRHR) indicators in humanitarian settings"

**Additional File 6: Link to our Tableau Dashboard**

Interactive Tableau Dashboard: Measurement of Sexual and Reproductive Health and Rights, Gender Equality, and Women's Empowerment in Humanitarian Settings.

https://public.tableau.com/profile/humairanakhuda#!/vizhome/SRHRGEWEScopingReviewStory_Final_Nov16/SRHRGEWEStory
