## Additional file 7 for "Tools for measuring sexual and reproductive health and rights (SRHR) indicators in humanitarian settings"

### Additional file 7: Study List

| Study ID | First Author | Title | Publication Year |
|---|---|---|---|
| 1. | Minza | Gender and Changes in Tsunami-Affected Villages in Nanggroe Aceh Darussalam province | 2005 |
| 2. | UN Women | Navigating through Shattered Paths: NGO Service Providers and Women Survivors of Gender-Based Violence | 2017 |
| 3. | United Nations Development Fund for Women | Pakistan Floods 2010: Rapid Gender Needs Assessment of Flood Affected Communities | 2010 |
| 4. | Toma | Rohingya Refugee Response Gender Analysis: Recognizing and responding to gender inequalities | 2018 |
| 5. | Slegh | Gender Relations, Sexual and Gender-Based Violence and the Effects of Conflict on Women and Men in North Kivu, Eastern Democratic Republic of the Congo: Results from the International Men and Gender Equality Survey (IMAGES) | 2014 |
| 6. | El Feki | Understanding Masculinities: Results from the International Men and Gender Equality Survey (IMAGES) - Middle East and North Africa | 2017 |
| 7. | Mansour | Understanding Masculinities: Results from the International Men and Gender Equality Survey (IMAGES) in Lebanon | 2017 |
| 8. | Bartels | Now the world is without me: An investigation of sexual violence in Eastern Democratic Republic of Congo | 2010 |
| 9. | Hirschfeld | Nowhere to turn: failure to protect, support and assure justice for Darfuri women | 2009 |
| 10. | Annan | The state of female youth in northern Uganda: findings from the survey of war-affected youth (SWAY) phase II | 2008 |

| | | | |
|---|---|---|---|
| 11. | McCoy | Baseline study: documenting knowledge, attitudes and behaviours of Somali refugees and the status of family planning services in UNHCR’s operation in Nairobi, Kenya | 2011 |
| 12. | Farzaneh | Baseline study: documenting knowledge, attitudes and behaviours of Somali refugees and the status of family planning services in UNHCR’s Ali Addeh site, Djibouti | 2011 |
| 13. | Amowitz | Human Rights Abuses and Concerns About Women’s Health and Human Rights in Southern Iraq | 2004 |
| 14. | Assaf | Domestic Violence Against Single, Never-Married Women in the Occupied Palestinian Territory | 2013 |
| 15. | Avdibegovic | Consequences of domestic violence on women's mental health in Bosnia and Herzegovina | 2006 |
| 16. | Bhardwaj | Interpersonal violence and suicidality among former child soldiers and war-exposed civilian children in Nepal | 2018 |
| 17. | Catani | War Trauma, Child Labor, and Family Violence: Life Adversities and PTSD in a Sample of School Children in Kabul | 2009 |
| 18. | Falb | Caregiver parenting and gender attitudes: Associations with violence against adolescent girls in South Kivu, Democratic Republic of Congo | 2017 |
| 19. | Green | Women's entrepreneurship and intimate partner violence: A cluster randomized trial of microenterprise assistance and partner participation in post-conflict Uganda | 2015 |
| 20. | Gupta | Gender norms and economic empowerment intervention to reduce intimate partner violence against women in rural Côte d’Ivoire: a randomized controlled pilot study | 2013 |
| 21. | Hossain | Working with men to prevent intimate partner violence in a conflict-affected setting: a pilot cluster randomized controlled trial in rural Côte d’Ivoire | 2014 |
| 22. | Kelly | From political to personal violence: Links between conflict and non-partner physical violence in post-conflict Liberia | 2019 |
| 23. | Khawaja | Prevalence of wife beating in Jordanian refugee camps: reports by men and women | 2005 |

| | | | |
|---|---|---|---|
| 24. | Khawaja | Attitudes of men and women towards wife beating: findings from Palestinian refugee camps in Jordan | 2007 |
| 25. | Kinyanda | Psychiatric disorders and psychosocial correlates of high hiv risk sexual behaviour in war-affected Eastern Uganda | 2012 |
| 26. | Loncar | Psychological consequences of rape on women in 1991-1995 War in Croatia and Bosnia and Herzegovina | 2006 |
| 27. | Merten | Sexuality education is associated with increased disclosure and treatment seeking after violence experiences of young women 15-24: Results from a multi-country-survey including Eastern DRC, Rwanda, and Burundi | 2015 |
| 28. | Mootz | Armed conflict, alcohol misuse, decision-making, and intimate partner violence among women in Northeastern Uganda: a population level study | 2018 |
| 29. | Okigbo | Risk factors for transactional sex among young females in post-conflict Liberia | 2014 |
| 30. | Ouma | Obstacles to family planning use among rural women of reproductive age in Atiak Health Centre IV, Amuru District, Northern Uganda | 2015 |
| 31. | Danish Refugee Council Protection Unit | A sexual and gender-based violence rapid assessment: Doro Refugee Camp, Upper Nile State, South Sudan | 2012 |
| 32. | Spencer | Gender Based Violence Against Women and Girls Displaced by the Syrian Conflict in South Lebanon and North Jordan: Scope of Violence and Health Correlates | 2015 |
| 33. | Khan | Midterm Evaluation: Epidemic Control and Reinforcement of Health Services (ECRHS) Project in Sierra Leone | 2017 |
| 34. | Wanyama | Diriswanaag Final Evaluation | 2013 |
| 35. | CARE Afghanistan Program Quality Unit | CARE International in Afghanistan Humanitarian Program (HP): Emergency shelter, NFI, Hygiene, SRHR and Livelihood Support for Disaster-Affected Populations in Afghanistan - 2018-2020, Baseline/KAP Survey Report | 2019 |

| | | | |
|---|---|---|---|
| 36. | Okello | Baseline evaluation of integrated emergency response program for South Sudanese refugees and affected host community members in Rhino and Imvepi settlements, Arua District | 2018 |
| 37. | International Institute of Independent Researchers (3IR) Pvt. Ltd. | Nepal Earthquake: Emergency Shelter and NFIs to Affected Households funded by Department of Foreign Affairs and Trade End-Line Assessment | 2016 |
| 38. | CARE International Switzerland | SDEARP Final Evaluation Report | 2012 |
| 39. | Ipsos | Our world. Views from the field. Summary Report: Afghanistan, Colombia, Democratic Republic of the Congo, Georgia, Haiti, Lebanon, Liberia and the Philippines | 2010 |
| 40. | Bhalla | The responsibility to prevent and respond to sexual and gender-based violence in disasters and crises | 2018 |
| 41. | Tanner | A safe place to shine: creating opportunities and raising voices of adolescent girls in humanitarian settings | 2017 |
| 42. | Murphy | No safe place: A lifetime of violence for conflict-affected women and girls in South Sudan | 2017 |
| 43. | Ssonko | Delivering HIV care in challenging operating environments: the MSF experience towards differentiated models of care for settings with multiple basic health care needs | 2017 |
| 44. | Reid | Providing HIV care in the aftermath of Kenya's post-election violence Medecins Sans Frontieres' lessons learned January - March 2008. | 2008 |
| 45. | Ferreyra | Provision and continuation of antiretroviral therapy during acute conflict: the experience of MSF in Central African Republic and Yemen | 2018 |
| 46. | Oxfam in Pakistan | Consolidated Gender Analysis for Disaster Response in Pakistan | 2017 |
| 47. | Wolfgang | From the Ground Up: Gender and conflict analysis in Yemen | 2016 |

| | | | |
|---|---|---|---|
| 48. | Dietrich | Gender and Conflict Analysis in ISIS Affected Communities of Iraq | 2017 |
| 49. | Oxfam | South Sudan Gender Analysis: A snapshot situation analysis of the differential impact of the humanitarian crisis on women, girls, men and boys in South Sudan | 2017 |
| 50. | Plan International | Adolescent Girls in Crisis: Experiences of risk and resilience across three humanitarian settings | 2018 |
| 51. | Stark | Building caregivers' emotional, parental and social support skills to prevent violence against adolescent girls: findings from a cluster randomised controlled trial in Democratic Republic of Congo | 2018 |
| 52. | Undie | Effectiveness of a community-based SGBV prevention model in emergency settings in Uganda: Testing the 'zero tolerance village alliance' intervention | 2016 |
| 53. | Family Health International | HIV and AIDS Behavioural Surveillance Survey (BSS): Refugee Camps and Hosting Communities in Kawambwa and Mporokoso, Zambia | 2006 |
| 54. | UNHCR | Refocusing Family Planning in Refugee Settings: Findings and Recommendations from a Multi-Country Baseline Study | 2011 |
| 55. | Kelly | Characterizing Sexual Violence in the Democratic Republic of the Congo. Profiles of Violence, Community Responses, and Implications for the Protection of Women | 2009 |
| 56. | Hudson | Picking up the pieces: Women's health needs assessment, Fond Parisien Region, Haiti | 2010 |
| 57. | Tomczyk | Women's reproductive health in Liberia: the Lofa County reproductive health survey | 2007 |
| 58. | Women's Wellness Center | Prevalence of gender-based violence: preliminary findings from a field assessment in nine villages in the Peja region, Kosovo | 2006 |
| 59. | Vinck | Association of exposure to violence and potential traumatic events with self-reported physical and mental health status in the Central African Republic | 2010 |
| 60. | Abbasi-Kangevari | Antenatal care utilisation among Syrian refugees in Tehran: A respondent driven sampling method | 2019 |

| | | | |
|---|---|---|---|
| 61. | Abu Hamad | Risk factors associated with preterm birth in the Gaza Strip: hospital-based case-control study | 2007 |
| 62. | Adam | The influence of maternal health education on the place of delivery in conflict settings of Darfur, Sudan | 2015 |
| 63. | Adam | Evidence from cluster surveys on the association between home-based counseling and use of family planning in conflict-affected Darfur | 2016 |
| 64. | Adam | Relationship between implementing interpersonal communication and mass education campaigns in emergency settings and use of reproductive healthcare services: evidence from Darfur, Sudan | 2015 |
| 65. | Agadjanian | Forced migration and hiv/aids risks in Angola | 2008 |
| 66. | Al-Khatib | Housing environment and women's health in a Palestinian refugee camp | 2005 |
| 67. | Al-Modallal | Effect of intimate partner violence on health of women of Palestinian origin | 2016 |
| 68. | Al-Modallal | Prevalence of intimate partner violence among women visiting health care centers in Palestine refugee camps in Jordan | 2015 |
| 69. | Al-Rukeimi | High rate of uterine rupture in a conflict setting of Hajjah, Yemen | 2017 |
| 70. | Al-Shdayfat | PHYSICAL ABUSE AMONG SYRIAN REFUGEE WOMEN IN JORDAN | 2017 |
| 71. | Dikmen | The attitudes of refugee women in Turkey towards family planning | 2018 |
| 72. | Alnuaimi | Pregnancy outcomes among Syrian refugee and Jordanian women: a comparative study | 2017 |
| 73. | Anastasi | Losing women along the path to safe motherhood: why is there such a gap between women's use of antenatal care and skilled birth attendance? A mixed methods study in northern Uganda | 2015 |
| 74. | Annan | Reducing PTSD symptoms through a gender norms and economic empowerment intervention to reduce intimate partner violence: A randomized controlled pilot study in Cote D'ivoire | 2017 |
| 75. | Anwar | Risk factors of posttraumatic stress disorder after an earthquake disaster | 2013 |

| | | | |
|---|---|---|---|
| 76. | Anwar | Reproductive health and access to healthcare facilities: risk factors for depression and anxiety in women with an earthquake experience | 2011 |
| 77. | Arnoso Martínez | Armed conflict, psychosocial impact and reparation in Colombia: Women's voice | 2017 |
| 78. | Bajracharya | Women of Nepal and post-earthquake humanitarian responses: An observation of three months | 2016 |
| 79. | Baloch | Screening of Reproductive Health Problems in Flood Affected Pregnant Women | 2012 |
| 80. | Balsara | Reproductive tract disorders among Afghan refugee women attending health clinics in Haripur, Pakistan | 2010 |
| 81. | Bardaweel | Impediments to use of oral contraceptives among refugee women in camps, Jordan | 2019 |
| 82. | Bartels | Patterns of sexual violence in Eastern Democratic Republic of Congo: Reports from survivors presenting to Panzi Hospital in 2006 | 2010 |
| 83. | Bell | Understanding the Effects of Mental Health on Reproductive Health Service Use: A Mixed Methods Approach | 2016 |
| 84. | Benage | An assessment of antenatal care among Syrian refugees in Lebanon | 2015 |
| 85. | Benner | Reproductive health and quality of life of young Burmese refugees in Thailand | 2010 |
| 86. | Betancourt | Sierra Leone's former child soldiers: a longitudinal study of risk, protective factors, and mental health | 2010 |
| 87. | Bhandari | Utilization of maternal health care services in post-conflict Nepal | 2015 |
| 88. | Blanc | The role of conflict in the rapid fertility decline in Eritrea and prospects for the future | 2004 |
| 89. | Boccia | High mortality associated with an outbreak of hepatitis E among displaced persons in Darfur, Sudan | 2006 |

| | | | |
|---|---|---|---|
| 90. | Borges | Women's reproductive health knowledge, attitudes and practices in relation to the Zika virus outbreak in northeast Brazil | 2018 |
| 91. | Bottcher | Maternal mortality in the Gaza strip: a look at causes and solutions | 2018 |
| 92. | Brissett | Zika Virus: Knowledge Assessment of Residents and Health-Care Providers in Roatan, Honduras, following an Outbreak | 2018 |
| 93. | Budhathoki | Menstrual hygiene management among women and adolescent girls in the aftermath of the earthquake in Nepal | 2018 |
| 94. | Campbell | Violence and abuse of internally displaced women survivors of the 2010 Haiti earthquake | 2016 |
| 95. | Carrara | Improved pregnancy outcome in refugees and migrants despite low literacy on the Thai-Burmese border: results of three cross-sectional surveys | 2011 |
| 96. | Casey | Availability of long-acting and permanent family-planning methods leads to increase in use in conflict-affected northern Uganda: evidence from cross-sectional baseline and endline cluster surveys | 2013 |
| 97. | Casey | Contraceptive availability leads to increase in use in conflict-affected Democratic Republic of the Congo: evidence from cross-sectional cluster surveys, facility assessments and service statistics | 2017 |
| 98. | Chen | Reproductive health for refugees by refugees in Guinea II: sexually transmitted infections | 2008 |
| 99. | Chukwumalu | Armed conflict and maternal health care utilization:Evidence from the Boko Haram insurgency in Nigeria | 2017 |
| 100. | Chukwumalu | Uptake of postabortion care services and acceptance of postabortion contraception in Puntland, Somalia | 2017 |
| 101. | Ciccio | Assessing the knowledge and behaviour towards HIV/AIDS among youth in Northern Uganda: A cross-sectional survey | 2009 |

| | | | |
|---|---|---|---|
| 102. | Correa | High burden of malaria and anemia among tribal pregnant women in a chronic conflict corridor in India | 2017 |
| 103. | Curry | Delivering high-quality family planning services in crisis-affected settings I: program implementation | 2015 |
| 104. | de Jong | Exposure to violence and PTSD symptoms among Somali women | 2011 |
| 105. | Dechen | Reproductive health naivety and perceived gender inequities among Tibetan refugee adolescent girls in India | 2011 |
| 106. | Decker | Factors associated with contraceptive use in Angola | 2011 |
| 107. | Delkhosh | Prevalence of intimate partner violence and reproductive health outcomes among Afghan refugee women in Iran | 2019 |
| 108. | Dong | Evaluating the micronutrient status of women of child-bearing age living in the rural disaster areas one year after Wenchuan Earthquake | 2014 |
| 109. | Dossa | Fistula and Other Adverse Reproductive Health Outcomes among Women Victims of Conflict-Related Sexual Violence: A population-based Cross-sectional study | 2014 |
| 110. | Dossa | Mental Health Disorders Among Women Victims of Conflict-Related Sexual Violence in the Democratic Republic of Congo | 2015 |
| 111. | Duff | High rates of Unintended Pregnancies among Young Women Sex Workers in Conflict-affected Northern Uganda: The Social Contexts of Brothels/Lodges and Substance Use | 2017 |
| 112. | Duroch | Description and consequences of sexual violence in Ituri province, Democratic Republic of Congo | 2011 |
| 113. | Edmond | Can community health worker home visiting improve care-seeking and maternal and newborn care practices in fragile states such as Afghanistan? A population-based intervention study | 2018 |
| 114. | Erenel | Clinical characteristics and pregnancy outcomes of Syrian refugees: a case-control study in a tertiary care hospital in Istanbul, Turkey | 2017 |

| | | | |
|---|---|---|---|
| 115. | Erickson | Structural determinants of dual contraceptive use among female sex workers in Gulu, northern Uganda | 2015 |
| 116. | Erickson | Incarceration and exposure to internally displaced persons camps associated with reproductive rights abuses among sex workers in northern Uganda | 2017 |
| 117. | Erickson | Interpersonal and structural contexts of intimate partner violence among female sex workers in conflict-affected northern Uganda | 2018 |
| 118. | Fabiani | A high prevalence of HIV-1 infection among pregnant women living in a rural district of north Uganda severely affected by civil strife | 2006 |
| 119. | Fabiani | HIV-1 prevalence and factors associated with infection in the conflict-affected region of North Uganda | 2007 |
| 120. | Falb | Violence against refugee women along the Thai-Burma border | 2013 |
| 121. | Falb | Symptoms associated with pregnancy complications along the Thai-Burma border: the role of conflict violence and intimate partner violence | 2014 |
| 122. | Foster | Community-based distribution of misoprostol for early abortion: evaluation of a program along the Thailand-Burma border | 2017 |
| 123. | Gamanga | The ebola outbreak: Effects on HIV reporting, testing and care in Bonthe district, Rural Sierra Leone | 2017 |
| 124. | Gebrecherkos | Unmet need for modern contraception and associated factors among reproductive age group women in Eritrean refugee camps, Tigray, north Ethiopia: a crosssectional study | 2018 |
| 125. | Getachew | Magnitude and factors associated with adherence to Iron-folic acid supplementation among pregnant women in Eritrean refugee camps, northern Ethiopia | 2018 |
| 126. | Gibbs | Factors associated with recent intimate partner violence experience amongst currently married women in Afghanistan and health impacts of IPV: a cross sectional study | 2018 |

| | | | |
|---|---|---|---|
| 127. | Gilbert | The experience of violence against children in domestic servitude in Haiti: Results from the Violence Against Children Survey, Haiti 2012 | 2018 |
| 128. | Gilder | Gestational diabetes mellitus prevalence in Maela refugee camp on the Thai-Myanmar border: a clinical report | 2014 |
| 129. | Gizelis | Maternal Health Care in the Time of Ebola: A Mixed-Method Exploration of the Impact of the Epidemic on Delivery Services in Monrovia | 2017 |
| 130. | Goldenberg | High burden of previously undiagnosed HIV infections and gaps in HIV care cascade for conflict-affected female sex workers in northern Uganda | 2019 |
| 131. | Hababeh | Contraceptive use by Palestine refugee mothers of young children attending UNRWA clinics: a cross-sectional follow-up study | 2018 |
| 132. | Hammoudeh | In search of health: Quality of life among postpartum Palestinian women | 2009 |
| 133. | Hammoury | Screening for domestic violence during pregnancy in an antenatal clinic in Lebanon | 2007 |
| 134. | Hammoury | Domestic violence against women during pregnancy: the case of Palestinian refugees attending an antenatal clinic in Lebanon | 2009 |
| 135. | Hannoun | Effect of war on the menstrual cycle | 2007 |
| 136. | Hapsari | Change in contraceptive methods following the Yogyakarta earthquake and its association with the prevalence of unplanned pregnancy | 2009 |
| 137. | Harrison | HIV behavioural surveillance among refugees and surrounding host communities in Uganda, 2006 | 2009 |
| 138. | Henwood | Ebola Virus Disease and Pregnancy: A Retrospective Cohort Study of Patients Managed at 5 Ebola Treatment Units in West Africa | 2017 |
| 139. | Ho | Using Program Data to Improve Access to Family Planning and Enhance the Method Mix in Conflict-Affected Areas of the Democratic Republic of the Congo | 2018 |

| | | | |
|---|---|---|---|
| 140. | Hoogenboom | Quality of intrapartum care by skilled birth attendants in a refugee clinic on the Thai-Myanmar border: a survey using WHO Safe Motherhood Needs Assessment | 2015 |
| 141. | Hossain | Men's and women's experiences of violence and traumatic events in rural Cote d'Ivoire before, during and after a period of armed conflict | 2014 |
| 142. | Howard | Reproductive health services for refugees by refugees in Guinea I: family planning | 2008 |
| 143. | Howard | Reproductive health for refugees by refugees in Guinea III: maternal health | 2011 |
| 144. | Husseini | HIV/AIDS-related knowledge and attitudes of Palestinian women in the Occupied Palestinian Territory | 2007 |
| 145. | Huster | Cesarean sections among Syrian refugees in Lebanon from december 2012/january 2013 to june 2013: probable causes and recommendations | 2014 |
| 146. | Hynes | A determination of the prevalence of gender-based violence among conflict-affected populations in East Timor | 2004 |
| 147. | Hynes | A study of refugee maternal mortality in 10 countries, 2008-2010 | 2012 |
| 148. | Ibrahim | Trauma and perceived social rejection among Yazidi women and girls who survived enslavement and genocide | 2018 |
| 149. | Ivanova | A cross-sectional mixed-methods study of sexual and reproductive health knowledge, experiences and access to services among refugee adolescent girls in the Nakivale refugee settlement, Uganda | 2019 |
| 150. | Johnson | Association of combatant status and sexual violence with health and mental health outcomes in postconflict liberia | 2008 |
| 151. | Johnson | Association of sexual violence and human rights violations with physical and mental health in territories of the Eastern Democratic Republic of the Congo | 2010 |
| 152. | Kabakian-Khasholian | Seeking maternal care at times of conflict: the case of Lebanon | 2013 |

| | | | |
|---|---|---|---|
| 153. | Khader | Anaemia among pregnant Palestinian women in the Occupied Palestinian Territory | 2009 |
| 154. | Khan | Maternal and newborn health situation of Rohingya migrants in cox's Bazar, Bangladesh | 2016 |
| 155. | Khawaja | Coerced sexual intercourse within marriage: a clinic-based study of pregnant Palestinian refugees in Lebanon | 2008 |
| 156. | Khawaja | Agreement between husband and wife reports of domestic violence: evidence from poor refugee communities in Lebanon | 2004 |
| 157. | Kidman | Intimate partner violence, modern contraceptive use and conflict in the Democratic Republic of the Congo | 2015 |
| 158. | Kim | HIV infection among internally displaced women and women residing in river populations along the Congo River, Democratic Republic of Congo | 2009 |
| 159. | Kim | Basic health, women's health, and mental health among internally displaced persons in Nyala Province, South Darfur, Sudan | 2007 |
| 160. | Kinyanda | War related sexual violence and it's medical and psychological consequences as seen in Kitgum, Northern Uganda: A cross-sectional study | 2010 |
| 161. | Kinyanda | Intimate partner violence as seen in post-conflict eastern Uganda: prevalence, risk factors and mental health consequences | 2016 |
| 162. | Kisindja | Family planning knowledge and use among women in camps for internally displaced people in the Democratic Republic of the Congo | 2017 |
| 163. | Kitabayashi | Association Between Maternal and Child Health Handbook and Quality of Antenatal Care Services in Palestine | 2017 |
| 164. | Kohli | A Congolese community-based health program for survivors of sexual violence | 2012 |
| 165. | Kohli | Risk for family rejection and associated mental health outcomes among conflict-affected adult women living in rural eastern Democratic Republic of the Congo | 2014 |

| | | | |
|---|---|---|---|
| 166. | Kottegoda | Reproductive health concerns in six conflict-affected areas of Sri Lanka | 2008 |
| 167. | Kruk | Availability of essential health services in post-conflict Liberia | 2010 |
| 168. | Landis | The school participation effect: investigating violence and formal education among girls in the Democratic Republic of the Congo | 2018 |
| 169. | Lafta | Needs of Internally Displaced Women and Children in Baghdad, Karbala, and Kirkuk, Iraq | 2016 |
| 170. | Larsen | Changes in HIV/AIDS/STI knowledge, attitudes and practices among commercial sex workers and military forces in Port Loko, Sierra Leone | 2004 |
| 171. | Lekskes | Appraisal of psychosocial interventions in Liberia | 2007 |
| 172. | Li | Influence of the Wenchuan earthquake on self-reported irregular menstrual cycles in surviving women | 2011 |
| 173. | Liu | A report on the reproductive health of women after the massive 2008 Wenchuan earthquake | 2010 |
| 174. | Logie | A psycho-educational HIV/STI prevention intervention for internally displaced women in Leogane, Haiti: results from a non-randomized cohort pilot study | 2014 |
| 175. | Loko Roka | One size fits all? Standardised provision of care for survivors of sexual violence in conflict and post-conflict areas in the Democratic Republic of Congo | 2014 |
| 176. | Lokuge | Mental health services for children exposed to armed conflict: Medecins Sans Frontieres' experience in the Democratic Republic of Congo, Iraq and the occupied Palestinian territory | 2013 |
| 177. | Lori | Behavior Change Following Implementation of Home-Based Life-Saving Skills in Liberia, West Africa | 2012 |
| 178. | Lori | Patient Satisfaction With Maternity Waiting Homes in Liberia: A Case Study During the Ebola Outbreak | 2017 |
| 179. | Malamba | The Cango lyec project-healing the elephant: Risk factors for HIV infection among post conflict populations in Northern Uganda | 2014 |

| | | | |
|---|---|---|---|
| 180. | Malamba | The Cango Lyec Project - Healing the Elephant: HIV related vulnerabilities of post-conflict affected populations aged 13-49 years living in three Mid-Northern Uganda districts | 2016 |
| 181. | Malemo Kalisya | Sexual violence toward children and youth in war-torn eastern Democratic Republic of Congo | 2011 |
| 182. | Mansson | Trends of HIV-1 and HIV-2 prevalence among pregnant women in Guinea-Bissau, West Africa: possible effect of the civil war 1998-1999 | 2007 |
| 183. | Martinez-Perez | Prevalence of Plasmodium falciparum infection among pregnant women at first antenatal visit in post-Ebola Monrovia, Liberia | 2018 |
| 184. | Mayhew | Determinants of skilled birth attendant utilization in Afghanistan: a cross-sectional study | 2008 |
| 185. | McGinn | Improving refugees' reproductive health through literacy in Guinea | 2006 |
| 186. | McGinn | Family planning in conflict: results of cross-sectional baseline surveys in three African countries | 2011 |
| 187. | McGready | Effect of early detection and treatment on malaria related maternal mortality on the north-western border of Thailand 1986-2010 | 2011 |
| 188. | McQuilkin | Health-Care Access during the Ebola Virus Epidemic in Liberia | 2017 |
| 189. | Meiksin | Domestic violence, marital control, and family planning, maternal, and birth outcomes in Timor-Leste | 2015 |
| 190. | Mels | Screening for traumatic exposure and posttraumatic stress symptoms in adolescents in the war-affected eastern Democratic Republic of Congo | 2009 |
| 191. | Mels | The psychological impact of forced displacement and related risk factors on Eastern Congolese adolescents affected by war | 2010 |
| 192. | Metheny | Help Seeking Behavior among Women Who Report Intimate Partner Violence in Afghanistan: an Analysis of the 2015 Afghanistan Demographic and Health Survey | 2019 |
| 193. | Mohammad | Postpartum depression symptoms among Syrian refugee women living in Jordan | 2010 |

| | | | |
|---|---|---|---|
| 194. | Morgos | Psychosocial effects of war experiences among displaced children in southern Darfur | 2007 |
| 195. | Moscardino | Mental health among former child soldiers and never-abducted children in Northern Uganda | 2012 |
| 196. | Muldoon | Policing the epidemic: High burden of workplace violence among female sex workers in conflict-affected northern Uganda | 2017 |
| 197. | Mullany | Access to essential maternal health interventions and human rights violations among vulnerable communities in eastern Burma | 2008 |
| 198. | Mullany | Impact of community-based maternal health workers on coverage of essential maternal health interventions among internally displaced communities in eastern Burma: the MOM project | 2010 |
| 199. | Murray | The impact of Cognitive Processing Therapy on stigma among survivors of sexual violence in eastern Democratic Republic of Congo: Results from a cluster randomized controlled trial | 2018 |
| 200. | Murty | Maternal health and maternal mortality in post war liberia: A survey analysis | 2013 |
| 201. | Nattabi | Family planning among people living with HIV in post-conflict Northern Uganda: A mixed methods study | 2011 |
| 202. | Nelson | Impact of sexual violence on children in the Eastern Democratic Republic of Congo | 2011 |
| 203. | O'Callaghan | A randomized controlled trial of trauma-focused cognitive behavioral therapy for sexually exploited, war-affected Congolese girls | 2013 |
| 204. | O'Laughlin | Feasibility and acceptability of home-based HIV testing among refugees: a pilot study in Nakivale refugee settlement in southwestern Uganda | 2018 |
| 205. | O'Laughlin | Predictors of HIV-infection during routine clinic-based HIV testing in Nakivale Refugee settlement in SW Uganda | 2016 |
| 206. | O'Laughlin | The cascade of HIV care among refugees and nationals in Nakivale Refugee Settlement in Uganda | 2017 |

| | | | |
|---|---|---|---|
| 207. | O'Laughlin | Clinic-based routine voluntary HIV testing in a refugee settlement in Uganda | 2014 |
| 208. | O'Laughlin | Predictors of HIV infection: a prospective HIV screening study in a Ugandan refugee settlement | 2016 |
| 209. | Obol | Knowledge and Misconceptions about Malaria among Pregnant Women in a Post-Conflict Internally Displaced Persons' Camps in Gulu District, Northern Uganda | 2011 |
| 210. | Ochola | HIV prevalence trend in the conflict to post-conflict transition period in Gulu District, Northern Uganda | 2013 |
| 211. | Odwe | Attitudes towards help-seeking for sexual and gender-based violence in humanitarian settings: the case of Rwamwanja refugee settlement scheme in Uganda | 2018 |
| 212. | Ojengbede | Sexual and gender-based violence in camps for internally displaced people and host communities in northeast Nigeria: a mixed methods study | 2019 |
| 213. | Okanlawon | Contraceptive use: knowledge, perceptions and attitudes of refugee youths in Oru Refugee Camp, Nigeria | 2010 |
| 214. | Orach | Perceptions, attitudes and use of family planning services in post conflict Gulu district, Northern Uganda | 2015 |
| 215. | Pack | Factors associated with unmet need for modern contraception in post-conflict Liberia | 2014 |
| 216. | Parker | Trends and birth outcomes in adolescent refugees and migrants on the Thailand-Myanmar border, 1986-2016: an observational study | 2018 |
| 217. | Parmar | Sexual violence among host and refugee population in Djohong District, Eastern Cameroon | 2012 |
| 218. | Patel | War and HIV: sex and gender differences in risk behaviour among young men and women in post-conflict Gulu District, Northern Uganda | 2014 |
| 219. | Patel | Comparison of HIV-related vulnerabilities between former child soldiers and children never abducted by the LRA in northern Uganda | 2013 |

| | | | |
|---|---|---|---|
| 220. | Pham | The use of a lot quality assurance sampling methodology to assess and manage primary health interventions in conflict-affected West Darfur, Sudan | 2016 |
| 221. | Pham | Validation of a screening tool to identify survivors of gender-based violence among displaced women in Colombia and Ethiopia | 2014 |
| 222. | Pham | Returning home: forced conscription, reintegration, and mental health status of former abductees of the Lord's Resistance Army in northern Uganda | 2009 |
| 223. | Plewes | Low seroprevalence of HIV and syphilis in pregnant women in refugee camps on the Thai-Burma border | 2008 |
| 224. | Potts | Measuring human rights violations in a conflict-affected country: results from a nationwide cluster survey in Central African Republic | 2011 |
| 225. | Pun | Exposure to domestic violence influences pregnant women's preparedness for childbirth in Nepal: A cross-sectional study | 2018 |
| 226. | Purdin | Reducing maternal mortality among Afghan refugees in Pakistan | 2009 |
| 227. | Qayum | Minimum Initial Service Package (MISP) access to displaced people of Pakistan based on Sphere Standards and Indicators | 2013 |
| 228. | Qayum | Frequency and physical factors associated with gender-based violence in the internally displaced people of Pakistan | 2012 |
| 229. | Raheel | Knowledge, attitudes and practices of contraception among Afghan refugee women in Pakistan: a cross-sectional study | 2012 |
| 230. | Rees | Intermittent explosive disorder amongst women in conflict affected Timor-Leste: Associations with human rights trauma, ongoing violence, poverty, and injustice | 2013 |
| 231. | Reese Masterson | Assessment of reproductive health and violence against women among displaced Syrians in Lebanon | 2014 |

| | | | |
|---|---|---|---|
| 232. | Rowley | Differences in HIV-related behaviors at Lugufu refugee camp and surrounding host villages, Tanzania | 2008 |
| 233. | Rutta | Prevention of mother-to-child transmission of HIV in a refugee camp setting in Tanzania | 2008 |
| 234. | Saile | Prevalence and predictors of partner violence against women in the aftermath of war: a survey among couples in northern Uganda | 2013 |
| 235. | Salami | High level of adherence to HAART among refugees and internally displaced persons on HAART in western equatorial region of Southern Sudan | 2010 |
| 236. | Sami | State of newborn care in South Sudan's displacement camps: a descriptive study of facility-based deliveries | 2017 |
| 237. | Scott | A mixed-methods assessment of sexual and gender-based violence in eastern Democratic Republic of Congo to inform national and international strategy implementation | 2013 |
| 238. | Seifeldin | Knowledge and utilization of family planning methods: A study from postconflict South Sudan | 2012 |
| 239. | Seyife | Utilization of modern contraceptives and predictors among women in Shimelba refugee camp, Northern Ethiopia | 2019 |
| 240. | Shuman | Perceptions and Experiences of Intimate Partner Violence in Abidjan, Cote d'Ivoire | 2016 |
| 241. | Sileo | A syndemic of psychosocial and mental health problems in Liberia: Examining the link to transactional sex among young pregnant women | 2019 |
| 242. | Silove | Pathways to perinatal depressive symptoms after mass conflict in Timor-Leste: a modelling analysis using cross-sectional data | 2015 |
| 243. | Simsek | A community-based survey on Syrian refugee women's health and its predictors in Sanliurfa, Turkey | 2018 |
| 244. | Sinha | Family planning in displaced populations: an unmet need among Iraqis in Amman, Jordan | 2008 |

| | | | |
|---|---|---|---|
| 245. | Sipsma | Violence against Congolese refugee women in Rwanda and mental health: a cross-sectional study using latent class analysis | 2015 |
| 246. | Sivaganesh | Antenatal care utilization in a conflict-affected district of Northern Sri Lanka | 2009 |
| 247. | Skokic | Perinatal and maternal outcomes in Tuzla Canton during 1992-1995 war in Bosnia and Herzegovina | 2006 |
| 248. | Sloand | Experiences of violence and abuse among internally displaced adolescent girls following a natural disaster | 2017 |
| 249. | Spiegel | High-risk sex and displacement among refugees and surrounding populations in 10 countries: the need for integrating interventions | 2014 |
| 250. | Spittal | Cango Lyec (Healing the Elephant): Gender Differences in HIV Infection in Post-conflict Northern Uganda | 2018 |
| 251. | Ssebunya | Prevalence and correlates of HIV testing among adolescents 10-19 years in a post-conflict pastoralist community of Karamoja region, Uganda | 2018 |
| 252. | Stark | Prevalence and associated risk factors of violence against conflict-affected female adolescents: a multi-country, cross-sectional study | 2017 |
| 253. | Stark | Measuring violence against women amidst war and displacement in northern Uganda using the "neighbourhood method" | 2010 |
| 254. | Stark | Measuring the incidence and reporting of violence against women and girls in liberia using the 'neighborhood method' | 2013 |
| 255. | Sukchan | Inadequacy of nutrients intake among pregnant women in the deep south of Thailand | 2010 |
| 256. | Sullivan | Using evidence to improve reproductive health quality along the Thailand-Burma border | 2004 |
| 257. | Sumner | Sentinel events predicting later unwanted sex among girls: A national survey in Haiti, 2012 | 2015 |

| | | | |
|---|---|---|---|
| 258. | Tanaka | Knowledge, attitude, and practice (KAP) of HIV prevention and HIV infection risks among Congolese refugees in Tanzania | 2008 |
| 259. | Tappis | Maternal Health Care Utilization Among Syrian Refugees in Lebanon and Jordan | 2017 |
| 260. | Tinuola | Insecurity and sexual rights violations of the female minors in internally displaced camps in Nigeria | 2016 |
| 261. | Tittle | Antenatal care among Palestine refugees in Jordan: factors associated with UNRWA attendance | 2019 |
| 262. | Truppa | Utilization of primary health care services among Syrian refugee and Lebanese women targeted by the ICRC program in Lebanon: a cross-sectional study | 2019 |
| 263. | Tsai | Medical evidence of human rights violations against non-Arabic-speaking civilians in Darfur: a cross-sectional study | 2012 |
| 264. | Usta | Child sexual abuse in Lebanon during war and peace | 2010 |
| 265. | Usta | Women, war, and violence: surviving the experience | 2008 |
| 266. | Usta | Women and health in refugee settings: The case of displaced syrian women in Lebanon | 2015 |
| 267. | Verelst | Mental health of victims of sexual violence in eastern Congo: associations with daily stressors, stigma, and labeling | 2014 |
| 268. | Verelst | The mediating role of stigmatization in the mental health of adolescent victims of sexual violence in Eastern Congo | 2014 |
| 269. | Vu | Psychometric properties and reliability of the Assessment Screen to Identify Survivors Toolkit for Gender Based Violence (ASIST-GBV): results from humanitarian settings in Ethiopia and Colombia | 2016 |
| 270. | Vu | Feasibility and acceptability of a universal screening and referral protocol for gender-based violence with women seeking care in health clinics in Dadaab refugee camps in Kenya | 2017 |

| | | | |
|---|---|---|---|
| 271. | Vyas | Marital violence and sexually transmitted infections among women in post-revolution Egypt | 2017 |
| 272. | Waheed | Maternal risk factors among pregnant internally displaced person women in Mardan, Pakistan | 2013 |
| 273. | Wako | Conflict, Displacement, and IPV: Findings From Two Congolese Refugee Camps in Rwanda | 2015 |
| 274. | walldorf | Recovery of HIV service provision post-earthquake | 2012 |
| 275. | Westhoff | Reproductive health education and services needs of internally displaced persons and refugees following disaster | 2009 |
| 276. | Wirtz | Lifetime prevalence, correlates and health consequences of gender-based violence victimisation and perpetration among men and women in Somalia | 2018 |
| 277. | Woodward | Reproductive health for refugees by refugees in Guinea IV: Peer education and HIV knowledge, attitudes, and reported practices | 2011 |
| 278. | yaya | Maternal health care service utilization in post-war Liberia: analysis of nationally representative cross-sectional household surveys | 2019 |
| 279. | Zhu | Fertility in older women following removal of long-term intrauterine devices in the wake of a natural disaster | 2013 |
| 280. | Zolala | Evaluation of the usefulness of maternal mortality ratio for monitoring long-term effects of a disaster: case study on the Bam earthquake | 2011 |
| 281. | Betancourt | Sierra Leone's child soldiers: war exposures and mental health problems by gender | 2011 |
| 282. | Clark | Association between exposure to political violence and intimate-partner violence in the occupied Palestinian territory: a cross-sectional study | 2010 |
| 283. | Djafri | Effect of the September 2009 Sumatra earthquake on reproductive health services and MDG 5 in the city of Padang, Indonesia | 2015 |
| 284. | Draebel | Prevalence of malaria and use of malaria risk reduction measures among resettled pregnant women in South Sudan | 2013 |

| | | | |
|---|---|---|---|
| 285. | Duff | Social and structural factors increase inconsistent condom use by sex workers' one-time and regular clients in Northern Uganda | 2018 |
| 286. | Eyobo | Malaria indicator survey 2009, South Sudan: baseline results at household level | 2014 |
| 287. | Flamand | The proportion of asymptomatic infections and spectrum of disease among pregnant women infected by Zika virus: systematic monitoring in French Guiana, 2016 | 2017 |
| 288. | Grbic | Vulnerability to HIV of internally displaced persons in the Republic of Serbia | 2014 |
| 289. | Izudi | Precancerous Cervix in Human Immunodeficiency Virus Infected Women Thirty Years Old and above in Northern Uganda | 2016 |
| 290. | Kaiser | HIV, syphilis, herpes simplex virus 2, and behavioral surveillance among conflict-affected populations in Yei and Rumbek, Southern Sudan | 2006 |
| 291. | Kennedy | Preliminary Impacts of an HIV-Prevention Program Targeting Out-of-School Youth in Postconflict Liberia | 2018 |
| 292. | Kim | Availability and quality of emergency obstetric and neonatal care services in Afghanistan | 2011 |
| 293. | Kinaro | Unsafe abortion and abortion care in Khartoum, Sudan | 2009 |
| 294. | Kitara | HIV/AIDS Stigmatization, the Reason for Poor Access to HIV Counseling and Testing (HCT) Among the Youths in Gulu (Uganda) | 2012 |
| 295. | Mayada | Conflicts in Yemen exacerbate lost to follow-up rates of people living with HIV | 2018 |
| 296. | McGready | Arthropod borne disease: the leading cause of fever in pregnancy on the Thai-Burmese border | 2010 |
| 297. | McGready | Diagnostic and treatment difficulties of pyelonephritis in pregnancy in resource-limited settings | 2010 |
| 298. | Morof | A cross-sectional survey on gender-based violence and mental health among female urban refugees and asylum seekers in Kampala, Uganda | 2014 |

| | | | |
|---|---|---|---|
| 299. | Muhammad | Malaria prevention practices and delivery outcome: a cross sectional study of pregnant women attending a tertiary hospital in northeastern Nigeria | 2016 |
| 300. | Negi | Impact of a massive earthquake on adherence to antiretroviral therapy, mental health, and treatment failure among people living with HIV in Nepal | 2018 |
| 301. | Ozel | Obstetric Outcomes among Syrian Refugees: A Comparative Study at a Tertiary Care Maternity Hospital in Turkey | 2018 |
| 302. | Parcesepe | Measuring Physical Violence and Rape Against Somali Women Using the Neighborhood Method | 2016 |
| 303. | Patel | Lost in transition: HIV prevalence and correlates of infection among young people living in post-emergency phase transit camps in Gulu District, Northern Uganda | 2014 |
| 304. | Roberts | Factors associated with post-traumatic stress disorder and depression amongst internally displaced persons in northern Uganda | 2008 |
| 305. | Shah | Unregulated usage of labour-inducing medication in a region of Pakistan with poor drug regulatory control: characteristics and risk patterns | 2016 |
| 306. | Tappis | Domestic violence among Iraqi refugees in Syria | 2012 |
| 307. | Tatah | Impact of Refugees on Local Health Systems: A Difference-in-Differences Analysis in Cameroon | 2016 |
| 308. | Guetiya Wadoum | Mobile health clinic for the medical management of clinical sequelae experienced by survivors of the 2013-2016 Ebola virus disease outbreak in Sierra Leone, West Africa | 2017 |
| 309. | Kolbe | Mortality, crime and access to basic needs before and after the Haiti earthquake: a random survey of Port-au-Prince households | 2010 |
| 310. | Sanguanklin | Effects of the 2011 flood in Thailand on birth outcomes and perceived social support | 2014 |

| | | | |
|---|---|---|---|
| 311. | Scott | Respondent-driven sampling to assess mental health outcomes, stigma and acceptance among women raising children born from sexual violence-related pregnancies in eastern Democratic Republic of Congo | 2015 |
| 312. | Turkay | Comparison of the pregnancy results between adolescent Syrian refugees and local adolescent Turkish citizens who gave birth in our clinic | 2018 |
| 313. | van Egmond | Reproductive health in Afghanistan: results of a knowledge, attitudes and practices survey among Afghan women in Kabul | 2004 |
| 314. | Weilg | Detection of Zika virus infection among asymptomatic pregnant women in the North of Peru | 2018 |
| 315. | Clark | Prevalence and risk factors for intimate partner violence in the West Bank and Gaza strip | 2011 |
| 316. | Kornilova | Restart of the HIV epidemic among PWID in occupied crimea and in the east of Ukraine | 2018 |
| 317. | McGready | Impact of ALSO (R) Australasia on PPH-Related Maternal Mortality on the Thailand-Myanmar Border in a Population Based Cohort Study | 2016 |
| 318. | Schumacher | The Relationship of Two Types of Trauma Exposure to Current Physical and Psychological Symptom Distress in a Community Sample of Colombian Women: Why Interpersonal Violence Deserves More Attention | 2010 |
| 319. | Makhoul | Impact of Syrian refugees on neonatal care in Hopital Notre dame de la paix, Akkar, north Lebanon | 2015 |
| 320. | Albutt | Stigmatisation and rejection of survivors of sexual violence in eastern Democratic Republic of the Congo | 2017 |
| 321. | Bannink | Prevention of spina bifida: Folic acid intake during pregnancy in Gulu district, northern Uganda | 2015 |
| 322. | Bannink | High PMTCT program uptake and coverage of mothers, their partners, and babies in Northern Uganda: Achievements and lessons learned over 10 years of implementation (2002-2011) | 2013 |

| | | | |
|---|---|---|---|
| 323. | Doliashvili | Women's sexual and reproductive health in post-socialist Georgia: does internal displacement matter? | 2008 |
| 324. | Duff | Intersecting reproductive health and HIV risks: Correlates of unintended pregnancies among a cohort of young women sex workers working in bars, truck-stops and lodges in postconflict Northern Uganda | 2013 |
| 325. | Muldoon | Alarming rates of occupational violence and associated HIV risks among young female sex workers in post-conflict northern Uganda | 2012 |
| 326. | Odjidja | Control of infectious disease during pregnancy among pastoralists in South Sudan: A case for investment into mobile clinics | 2018 |
| 327. | Shannon | Hyper-endemic HIV seroprevalence among young female sex workers in post-conflict, northern Uganda: A call for social and structural HIV interventions | 2012 |
| 328. | Abdalla | The need for a comprehensive response to HIV/ AIDS in north-western Somalia: evidence from a seroprevalence survey | 2007 |
| 329. | Bamrah | The impact of post-election violence on HIV and other clinical services and on mental health-Kenya, 2008 | 2013 |
| 330. | Bass | Controlled trial of psychotherapy for Congolese survivors of sexual violence | 2013 |
| 331. | Bayo | Estimating the met need for emergency obstetric care (EmOC) services in three payams of Torit County, South Sudan: a facility-based, retrospective cross-sectional study | 2018 |
| 332. | Callands | Experiences and acceptance of intimate partner violence: associations with sexually transmitted infection symptoms and ability to negotiate sexual safety among young Liberian women | 2013 |
| 333. | Falb | Recent abuse from in-laws and associations with adverse experiences during the crisis among rural Ivorian women: extended families as part of the ecological model | 2013 |
| 334. | Klasen | Multiple trauma and mental health in former Ugandan child soldiers | 2010 |

| | | | |
|---|---|---|---|
| 335. | Lash | 1999 earthquake of Marmara, Turkey, women’s health, and nursing care: leadership in action | 2008 |
| 336. | Lau | Severe antenatal depressive symptoms before and after the 2008 Wenchuan earthquake in Chengdu, China | 2011 |
| 337. | Maalim | Supporting 'medicine at a distance' for delivery of hospital services in war-torn Somalia: how well are we doing? | 2014 |
| 338. | Nagai | Violence against refugees, non-refugees and host populations in southern Sudan and northern Uganda | 2008 |
| 339. | Nakimuli-Mpungu | Implementation and Scale-Up of Psycho-Trauma Centers in a Post-Conflict Area: A Case Study of a Private-Public Partnership in Northern Uganda | 2013 |
| 340. | Qayum | Assessment of health services on relevant primary health care principles in internally displaced people of pakistan based on sphere standards and indicators | 2011 |
| 341. | Renzaho | Mortality, malnutrition and the humanitarian response to the food crises in Lesotho | 2006 |
| 342. | Salisbury | Family planning knowledge, attitudes and practices in refugee and migrant pregnant and post-partum women on the Thailand-Myanmar border -- a mixed methods study | 2016 |
| 343. | Schwab | Predictive factors for preterm delivery under rural conditions in post-tsunami Banda Aceh | 2016 |
| 344. | Sousa | The Co-Occurrence and Unique Mental Health Effects of Political Violence and Intimate Partner Violence | 2018 |
| 345. | Hadi | Raising institutional delivery in war-torn communities: experience of BRAC in Afghanistan | 2007 |
| 346. | Tanabe | Family planning in refugee settings: Findings and actions from a multi-country study | 2017 |
| 347. | Steiner | Sexual violence in the protracted conflict of DRC programming for rape survivors in South Kivu | 2009 |
| 348. | Kelly | Experiences of female survivors of sexual violence in eastern Democratic Republic of the Congo: a mixed-methods study | 2011 |
| 349. | Kinyanda | Prevalence and correlates of psychological distress as seen in post-conflict Liberia | 2010 |

| | | | |
|---|---|---|---|
| 350. | Baelani | Facing medical care problems of victims of sexual violence in Goma/Eastern Democratic Republic of the Congo | 2011 |
| 351. | Klasen | Posttraumatic Resilience in Former Ugandan Child Soldiers. | 2010 |
| 352. | Qu | Posttraumatic stress disorder and depression among new mothers at 8 months later of the 2008 Sichuan earthquake in China | 2012 |
| 353. | Pham | The use of lot quality assurance sampling in the assessment of health and water/sanitation services in a complex humanitarian emergency | 2012 |
| 354. | Ameh | Challenges to the provision of emergency obstetric care in Iraq | 2011 |
| 355. | Al-Nuaimi | Community violence and mental health among Iraqi women, a population-based study | 2013 |
| 356. | Zambrano | High incidence of Zika virus infection detected in plasma and cervical cytology specimens from pregnant women in Guayaquil, Ecuador | 2017 |
| 357. | Wollum | Requests for medication abortion support in Brazil during and after the zika epidemic | 2018 |
| 358. | Pena | Zika virus epidemic in pregnant women, Dominican Republic, 2016-2017 | 2019 |
| 359. | Borges | Zika Virus Outbreak - Should assisted reproduction patients avoid pregnancy? | 2017 |
| 360. | Zambrano | Detection of zika virus and cytomegalovirus in cervical cytology samples of pregnant women from guayaquil, ecuador, using two real-time polymerase chain reaction (RT-PCR) molecular assays | 2017 |
| 361. | Joao | Pregnant women co-infected with HIV and Zika: Outcomes and birth defects in infants according to maternal symptomatology | 2018 |
| 362. | de Oliveira | Infection-related microcephaly after the 2015 and 2016 Zika virus outbreaks in Brazil: a surveillance-based analysis | 2017 |
| 363. | Azeredo | Clinical and Laboratory Profile of Zika and Dengue Infected Patients: Lessons Learned From the Co-circulation of Dengue, Zika and Chikungunya in Brazil | 2018 |

| | | | |
|---|---|---|---|
| 364. | Alemayehu | Prevalence, Severity, and Determinant Factors of Anemia among Pregnant Women in South Sudanese Refugees, Pugnido, Western Ethiopia | 2016 |
| 365. | Ganle | Risky sexual behaviour and contraceptive use in contexts of displacement: insights from a cross-sectional survey of female adolescent refugees in Ghana | 2019 |
| 366. | Goessmann | The contribution of mental health and gender attitudes to intimate partner violence in the context of war and displacement: Evidence from a multi-informant couple survey in Iraq | 2019 |
| 367. | Ray-Bennett | Understanding reproductive health challenges during a flood: Insights from Belkuchi Upazila, Bangladesh | 2019 |
| 368. | Sileo | Trauma Exposure and Intimate Partner Violence Among Young Pregnant Women in Liberia | 2019 |
| 369. | Bouchghoul | Humanitarian obstetric care for refugees of the Syrian war. The first 6 months of experience of Gynécologie Sans Frontières in Zaatari Refugee Camp (Jordan) | 2015 |
| 370. | Tuncer | Predictors of adverse maternal and perinatal outcomes in a refugee population from an active conflict country, Syria | 2019 |
| 371. | Erenoglu | The Effect of Health Education Given to Syrian Refugee Women in Their Own Language on Awareness of Breast and Cervical Cancer, in Turkey: a Randomized Controlled Trial | 2019 |
| 372. | Ssebunya | Factors associated with prior engagement in high-risk sexual behaviours among adolescents (10-19 years) in a pastoralist post-conflict community, Karamoja sub-region, North eastern Uganda | 2019 |
| 373. | Murphy | The effects of conflict and displacement on violence against adolescent girls in South Sudan: the case of adolescent girls in the Protection of Civilian sites in Juba | 2019 |
| 374. | de Jong, Kaz | Conflict in the Indian Kashmir Valley I: exposure to violence | 2008 |
| 375. | Falb | Depressive symptoms among women in Raqqa Governorate, Syria: associations with intimate partner violence, food insecurity, and perceived needs | 2019 |

| | | | |
|---|---|---|---|
| 376. | Mukunya | Inequity in utilization of health care facilities during childbirth: a community-based survey in post-conflict Northern Uganda | 2019 |
| 377. | Seff | Forced Sex and Early Marriage: Understanding the Linkages and Norms in a Humanitarian Setting | 2019 |
| 378. | Al-Maharma | Knowledge, attitudes and practices of syrian refugee mothers towards sexually transmitted infections | 2019 |
| 379. | Godwin | Reproductive health sequelae among women who survived Ebola virus disease in Liberia | 2019 |
| 380. | Casey | Meeting the demand of women affected by ongoing crisis: Increasing contraceptive prevalence in North and South Kivu, Democratic Republic of the Congo | 2019 |
| 381. | Torun | Health and health care access for Syrian refugees living in Istanbul | 2018 |
| 382. | Pierce | Reproductive health care utilization among refugees in Jordan: Provisional support and domestic violence | 2019 |
| 383. | Morris | When political solutions for acute conflict in Yemen seem distant, demand for reproductive health services is immediate: a programme model for resilient family planning and post-abortion care services | 2019 |
| 384. | Bartels | Surviving sexual violence in eastern Democratic Republic of Congo | 2010 |
| 385. | Kalter | Prospective community based cluster census and case-control study of stillbirths and neonatal deaths in the West Bank and Gaza Strip | 2008 |
