## Additional file 8 for "Tools for measuring sexual and reproductive health and rights (SRHR) indicators in humanitarian settings"

### Additional file 8: SRHR Domains, Indicator Types, and Indicator Subtypes

| | Counted once per study | Counted once per country |
|---|---|---|
| **Abortion** | | |
| **Abortion procedures & post abortion care** | **6** | **6** |
| Received post-abortion care | 5 | 5 |
| Timing of abortion treatment | 1 | 1 |
| Type of abortion procedure | 4 | 4 |
| **Experience of abortion** | **24** | **18** |
| Experience of self-induced abortion | 1 | 1 |
| Experienced an abortion | 21 | 17 |
| Reason for legal abortion | 1 | 1 |
| Zika as a reason for abortion | 2 | 1 |
| **Location of abortion** | **4** | **4** |
| Facility abortion | 3 | 3 |
| Home abortion | 1 | 1 |
| Knowledge of where to receive an abortion | 1 | 1 |
| **Antenatal Care (ANC)** | | |
| **Access to ANC** | **35** | **20** |
| ANC Knowledge | 5 | 4 |
| Accessibility of ANC | 7 | 6 |
| Barriers to ANC | 9 | 8 |
| Discussion of ANC with provider | 1 | 1 |
| Location of ANC | 5 | 5 |
| Provision of ANC information | 4 | 4 |
| Received any ANC | 13 | 11 |
| Timing of first ANC visit | 6 | 5 |
| **ANC Procedures** | **27** | **19** |
| ANC testing | 5 | 5 |
| ART use during pregnancy | 4 | 4 |
| Antibiotic use during pregnancy | 1 | 1 |
| HIV/STI testing during pregnancy | 6 | 6 |
| IFA supplementation during pregnancy | 9 | 7 |
| Malaria treatment/prevention during pregnancy | 8 | 5 |
| Other treatment during ANC | 6 | 4 |
| Pregnancy risk at visit | 1 | 1 |
| Received counselling during ANC | 2 | 2 |
| Tetanus toxoid during pregnancy | 6 | 6 |
| **Number of ANC visits** | **37** | **24** |
| Attended 1-3 ANC visits | 11 | 14 |
| Attended 4+ visits | 15 | 19 |

| | | |
|---|---|---|
| Attended at least 1 ANC visit | 9 | 7 |
| Did not attend ANC | 15 | 17 |
| Other measures of ANC attendance | 12 | 8 |
| **Type of health provider at ANC visit** | **9** | **15** |
| Received ANC from a trained traditional birth attendant | 2 | 2 |
| Received ANC from skilled health provider | 7 | 14 |
| Received ANC from unskilled provider | 3 | 10 |
| Received a CHW visit during pregnancy | 1 | 1 |
| **Family Planning** | | |
| **Access to FP/Contraception** | **59** | **30** |
| Access to family planning services/contraception | 18 | 14 |
| Availability of family planning services/contraception | 2 | 2 |
| Desired method of contraception | 4 | 4 |
| Knowledge of where to get contraception | 2 | 2 |
| Location of FP services | 8 | 9 |
| Reasons for FP use | 3 | 3 |
| Reasons for no FP use | 16 | 14 |
| Unmet need for FP | 18 | 16 |
| Using some form of family planning | 29 | 21 |
| **Contraceptive effectiveness** | **6** | **5** |
| Contraceptive failure rate | 1 | 1 |
| Side effects of contraceptives | 4 | 4 |
| Unplanned pregnancy | 3 | 3 |
| **Ever used contraception** | **26** | **15** |
| Ever used a family planning method | 8 | 9 |
| Ever used modern contraceptives | 20 | 13 |
| **Family Planning/Contraception Knowledge** | **44** | **23** |
| Aware of family planning | 29 | 19 |
| Could identify family planning method | 3 | 2 |
| Knowledge of FP benefits | 6 | 6 |
| Knowledge of FP safety and effectiveness | 6 | 5 |
| Knowledge of where to get contraception | 8 | 8 |
| Received family planning information/instruction | 21 | 13 |
| Source of family planning knowledge | 9 | 6 |
| **Modern contraceptive use** | **56** | **34** |
| Condom use | 43 | 32 |
| Contraceptive prevalence rate | 3 | 7 |
| Hormonal contraceptive use | 4 | 4 |
| IUD use | 23 | 19 |
| Implant use | 15 | 11 |
| Injectable use | 29 | 22 |

| | | |
|---|---|---|
| Modern contraceptive use | 31 | 25 |
| Oral contraceptive use | 29 | 23 |
| Permanent contraception use | 14 | 12 |
| Use of emergency contraceptives | 6 | 6 |
| **Traditional contraceptive use** | **17** | **17** |
| Traditional contraceptive use | 8 | 13 |
| Use of rhythm method | 7 | 6 |
| Use of withdrawal method | 10 | 10 |
| Using abstinence | 5 | 5 |
| **Gender-Based Violence** | | |
| **Domestic violence** | **27** | **17** |
| Experienced domestic violence | 21 | 16 |
| Experienced economic domestic violence | 1 | 2 |
| Experienced emotional domestic violence | 3 | 3 |
| Experienced physical domestic violence | 5 | 6 |
| Experienced sexual domestic violence | 4 | 4 |
| Frequency of domestic violence | 2 | 2 |
| **Economic violence** | **2** | **3** |
| Experienced economic violence | 2 | 3 |
| **Emotional/Psychological violence** | **24** | **18** |
| Experienced emotional violence | 12 | 11 |
| Experienced psychological violence | 7 | 5 |
| Experienced threats of violence | 3 | 3 |
| Frequency of emotional violence | 1 | 1 |
| Type of emotional violence experienced | 4 | 10 |
| **Experienced any violence** | **41** | **18** |
| Experienced any violence | 13 | 11 |
| Experienced conflict victimization | 8 | 6 |
| Experienced gender-based violence | 4 | 5 |
| Experienced impact of violence | 3 | 3 |
| Experienced other form of violence | 9 | 5 |
| Experienced physical or sexual violence | 12 | 10 |
| Mean violence scores | 2 | 2 |
| **Experienced trauma** | **10** | **13** |
| Experience of torture | 3 | 9 |
| Experienced trafficking | 1 | 1 |
| Experienced trauma | 3 | 3 |
| Knowledge of human trafficking | 1 | 1 |
| Mental health symptoms of trauma | 2 | 2 |
| **Female circumcision and other traditional practices** | **3** | **3** |
| Experienced female circumcision | 2 | 2 |

| | | |
|---|---|---|
| Reporting harmful religious practices | 1 | 1 |
| **GBV knowledge and services** | **15** | **10** |
| Access to GBV services | 5 | 3 |
| Knowledge of services for violence survivors | 4 | 5 |
| Knowledge/Awareness of GBV | 2 | 2 |
| Post GBV treatment and procedures | 3 | 2 |
| Satisfaction with GBV information/services | 4 | 4 |
| Sought care for GBV | 6 | 5 |
| Timing of post GBV care | 4 | 1 |
| **Intimate partner violence (IPV)** | **61** | **30** |
| Experienced IPV | 38 | 25 |
| Experienced IPV resulting in injury | 6 | 6 |
| Experienced economic IPV | 4 | 4 |
| Experienced emotional/psychological IPV | 22 | 15 |
| Experienced lifetime IPV | 11 | 11 |
| Experienced physical IPV | 29 | 16 |
| Experienced sexual IPV | 31 | 20 |
| IPV during pregnancy | 2 | 2 |
| Type of physical IPV | 11 | 11 |
| **Morbidity due to sexual violence** | **36** | **19** |
| Anxiety due to sexual violence | 4 | 1 |
| Depression due to sexual violence | 6 | 2 |
| Experienced STI/fear of STI as a result of sexual violence | 8 | 7 |
| Experienced abdominal pain due to sexual violence | 6 | 4 |
| Experienced behaviour changes due to sexual violence | 7 | 3 |
| Experienced bleeding due to sexual violence | 4 | 3 |
| Experienced fistula/genital trauma due to sexual violence | 7 | 4 |
| Experienced lesions due to sexual violence | 1 | 1 |
| Experienced other morbidities due to sexual violence | 10 | 7 |
| Experienced pregnancy as a result of rape | 17 | 11 |
| PTSD due to sexual violence | 8 | 3 |
| Symptoms of child sexual violence | 1 | 1 |
| **Non-partner violence (NPV)** | **7** | **7** |
| Experienced other NPV | 3 | 4 |
| Experienced physical NPV | 4 | 5 |
| Experienced sexual NPV | 4 | 5 |
| Type of community violence experienced | 1 | 1 |
| **Physical violence** | **53** | **29** |
| Experienced physical violence | 32 | 19 |

| | | |
|---|---|---|
| Experienced physical violence as a child | 3 | 3 |
| Experienced physical violence during pregnancy | 2 | 2 |
| Experienced physical violence resulting in injury | 8 | 12 |
| Frequency of physical violence | 1 | 1 |
| Type of physical violence experienced | 19 | 16 |
| Type of physical violence perpetrated by armed forces | 4 | 3 |
| **Sexual violence** | **110** | **38** |
| Experienced sexual violence | 98 | 36 |
| Experienced sexual violence before the age of 18 | 6 | 6 |
| Experienced sexual violence during pregnancy | 2 | 2 |
| Frequency of sexual violence | 12 | 8 |
| Type of sexual violence experienced | 43 | 18 |
| **HIV & Sexually Transmitted Infections** | | |
| **Access to HIV Services** | **17** | **10** |
| Reasons for not receiving HIV care | 4 | 4 |
| Received HIV care | 13 | 8 |
| **Antiretroviral therapy (ART)** | **14** | **10** |
| ART adherence | 7 | 7 |
| ART use | 11 | 8 |
| **HIV knowledge** | **29** | **19** |
| Comprehensive HIV knowledge | 6 | 5 |
| General HIV knowledge/awareness | 21 | 16 |
| HIV disclosure | 3 | 1 |
| HIV misconceptions | 1 | 1 |
| Knowledge of HCT | 1 | 1 |
| Knowledge of HIV prevention/transmission | 14 | 8 |
| Source of HIV information | 5 | 5 |
| **HIV/STI infection** | **59** | **24** |
| HIV positive | 37 | 15 |
| HIV prevalence | 10 | 1 |
| HIV-related death | 1 | 2 |
| STI infection | 15 | 11 |
| STI symptoms | 14 | 7 |
| Stage of HIV symptoms | 3 | 3 |
| Syphilis infection | 9 | 6 |
| **HIV/STI testing** | **30** | **17** |
| CD4 testing | 4 | 4 |
| HIV testing | 23 | 15 |
| STI testing | 4 | 1 |
| **STI knowledge** | **10** | **9** |
| General STI knowledge | 9 | 9 |

| | | |
|---|---|---|
| Knowledge of STI symptoms | 3 | 2 |
| **STI treatment** | **9** | **9** |
| Reasons for not seeking STI treatment | 1 | 1 |
| Sought STI treatment | 9 | 9 |
| **Maternal Health** | | |
| **Access to Maternal Health Care** | **8** | **7** |
| Access to MH services | 7 | 6 |
| Conceived with assisted reproductive technology | 1 | 1 |
| **Illness/complications during pregnancy** | **52** | **23** |
| Anaemia during pregnancy | 14 | 12 |
| Ebola during pregnancy | 1 | 2 |
| Experienced bleeding during pregnancy | 10 | 9 |
| Experienced other pregnancy complication | 23 | 12 |
| Experienced pre-eclampsia | 1 | 1 |
| Gestational diabetes | 7 | 6 |
| Hepatitis E during pregnancy | 1 | 1 |
| HIV during pregnancy | 1 | 1 |
| Illness during pregnancy | 2 | 2 |
| Infection during pregnancy | 7 | 4 |
| Malaria during pregnancy | 9 | 6 |
| Malaria prevention during pregnancy | 5 | 4 |
| Malnutrition during pregnancy | 4 | 4 |
| Pyelonephritis during pregnancy | 2 | 1 |
| Symptoms of arthropod infection during pregnancy | 1 | 1 |
| Symptoms of Hepatitis E during pregnancy | 1 | 1 |
| Symptoms of Zika virus infection in pregnancy | 3 | 3 |
| Zika infection during pregnancy | 9 | 5 |
| **Postnatal care (PNC)** | **17** | **13** |
| Experienced post-partum complications | 1 | 1 |
| Postnatal care attendance | 10 | 8 |
| Postnatal care procedures | 6 | 6 |
| Post-partum depression | 3 | 3 |
| **Pregnancy knowledge** | **15** | **10** |
| General pregnancy knowledge | 3 | 2 |
| Knowledge of pregnancy danger signs | 5 | 5 |
| Malaria knowledge during pregnancy | 2 | 2 |
| Received maternal health education | 4 | 4 |
| Source of information about maternity waiting homes | 1 | 1 |
| Zika knowledge during pregnancy | 2 | 2 |
| **Maternal Mortality** | | |
| **Cause of maternal death** | **8** | **17** |

| | | |
|---|---|---|
| Contributing cause of death | 1 | 10 |
| Died of eclampsia | 2 | 2 |
| Died of indirect causes | 2 | 11 |
| Died of obstetric hemorrhage | 4 | 13 |
| Died of other direct obstetric causes | 7 | 16 |
| Died of sepsis | 3 | 12 |
| Reasons for maternal death | 1 | 10 |
| Unknown cause of death | 2 | 11 |
| **Location of maternal death** | **2** | **11** |
| Location of death | 1 | 10 |
| Timing of death | 2 | 11 |
| **Measure of maternal death** | **14** | **20** |
| MMR | 5 | 14 |
| Obstetric case fatality rate | 3 | 3 |
| Proportion of maternal deaths | 10 | 9 |
| **Menstruation & Gynecological Health** | | |
| **Access to SRH Care** | **19** | **15** |
| Accessed SRH services | 10 | 8 |
| Did not receive, seek, or practice SRH care | 3 | 3 |
| SRH knowledge | 4 | 4 |
| Type of SRH treatment/practice | 11 | 8 |
| **Menstrual Hygiene Management (MHM)** | **10** | **8** |
| Access to MHM | 7 | 6 |
| Knowledge of menstruation | 2 | 2 |
| Satisfaction with hygiene kit | 1 | 1 |
| Type of MHM used | 3 | 3 |
| **Menstrual irregularities** | **10** | **7** |
| Type of menstrual irregularity | 10 | 7 |
| **SRH symptoms** | **17** | **10** |
| Anemia | 3 | 3 |
| Gynecological complaints | 11 | 7 |
| Pelvic complaints | 5 | 5 |
| Reproductive tract infections | 3 | 3 |
| **Obstetric Care (Delivery)** | | |
| **Access to EmOC** | **5** | **4** |
| Accessibility of EmOC | 1 | 1 |
| Availability of EmOC | 1 | 1 |
| Inadequate EmOC | 2 | 2 |
| Met need for EmOC | 2 | 2 |
| **Assisted Delivery** | **24** | **16** |
| Delivery by skilled birth attendant | 18 | 14 |

| | | |
|---|---|---|
| Delivery by traditional birth attendant | 12 | 9 |
| Other assisted delivery | 7 | 6 |
| Planning for delivery | 6 | 4 |
| Reason for no delivery assistance | 1 | 1 |
| Source of medication | 1 | 1 |
| **Delivery Location** | **34** | **21** |
| Delivery outside of a health facility | 21 | 17 |
| Facility delivery | 28 | 17 |
| Home delivery | 4 | 3 |
| Reasons for delivery location | 1 | 1 |
| **Delivery knowledge** | **8** | **5** |
| Knowledge of safe delivery | 2 | 2 |
| Preparations made for delivery | 3 | 3 |
| Source of obstetric referral | 3 | 2 |
| **Delivery procedures** | **29** | **15** |
| C-section delivery | 28 | 15 |
| Delivery room | 2 | 2 |
| Induction of labour | 5 | 4 |
| Other treatment administered during delivery | 10 | 5 |
| Reason for c-section | 3 | 3 |
| Received injections | 1 | 1 |
| Removal/examination of the placenta | 2 | 2 |
| Treatment for sepsis | 1 | 1 |
| Treatment of post-partum hemorrhage | 2 | 2 |
| Vacuum delivery | 2 | 1 |
| **Miscarriage & Stillbirths** | **40** | **22** |
| Experienced miscarriage | 29 | 17 |
| Experienced stillbirth | 18 | 12 |
| Miscarriage or stillbirth | 2 | 2 |
| **Obstetric complication** | **28** | **14** |
| Experienced fetal complication | 4 | 4 |
| Experienced haemorrhage | 10 | 8 |
| Experienced obstetrical hysterectomy | 2 | 2 |
| Experienced other delivery complication | 21 | 12 |
| Experienced placenta previa | 1 | 1 |
| Experienced pre-eclampsia/eclampsia | 12 | 8 |
| Experienced premature rupture of membranes | 9 | 6 |
| Experienced preterm birth | 7 | 7 |
| Experienced prolonged/obstructed labour | 11 | 9 |
| Experienced sepsis | 4 | 4 |
